# Mono- and bi-allelic effects of coding variants on disease in 176,899 Finns

**DOI:** 10.1101/2021.11.06.21265920

**Authors:** H. O. Heyne, J. Karjalainen, K. J. Karczewski, S. M. Lemmelä, W. Zhou, FinnGen, A. S. Havulinna, M. Kurki, H. L. Rehm, A. Palotie, M. J. Daly

**Author notes:** corresponding authors, P.O. Box 20, FI-00014 University of Helsinki, FINLAND.

## Abstract

Identifying Mendelian diseases with recessive inheritance is challenging as the majority of cases are caused by compound heterozygous genotypes which require sequencing data in families to definitively identify. Bottleneck events, such as in the Finnish population, enrich specific homozygous variants to higher frequencies and thus facilitate identification of disease associations through easily recognized homozygous genotypes. Here, we study homozygous and heterozygous effects of 82,516 coding variants on 2,444 disease endpoints using nationwide electronic health record (EHR) data of 176,899 Finns. We find known and novel associations to homozygous genotypes across a broad spectrum of phenotypes such as retinal dystrophy, adult-onset cataract and female infertility (13/20 of which would have been missed by the traditional additive GWAS model). With these results, and supporting simulations, we demonstrate the added benefit of homozygous scans in GWAS. We further use these results to explore inheritance patterns of known Mendelian variants. We find many Mendelian variants whose inheritance cannot be adequately described with the traditional definition of dominant or recessive. In particular, we find disease risk in heterozygous carriers of variants known to cause disease with recessive inheritance, as well as for variants labeled benign in ClinVar. Our results demonstrate how biobank efforts, particularly in founder populations, can broaden our understanding of the impact of genetic variants.

## Main

### Introduction

Rare genetic variants with large effects on disease can have potential direct treatment implications^1,2^; studying their effects comprehensively requires however large sample sizes^3^. Identifying new bi-allelic effects on disease is particularly challenging, as the majority of causal variants with autosomal recessive inheritance are compound heterozygous in most individuals from non-isolated and non-consanguineous populations^4^, owing to selective pressure keeping individual variants at low frequency. However, this is not the case in populations that have undergone bottleneck events such as Ashkenazi Jews^5^, Icelanders^6^, French Canadians^7^, Anabaptist groups^8^, Puerto Ricans^9^ or Finns^10^. The Finnish population has been small and relatively isolated with lower genetic diversity than other European populations^11^. Multiple bottleneck events in the Finnish population history can today be detected as geographically distinct genetic clusters that partially correspond to historical events and local dialects^12,13^. Such geographically distinct genetic clusters are also characterized by higher rates of haplotype sharing^12,14^ and thus, higher rates of homozygosity increasing the chance occurrence of pathogenic variants in a homozygous state that lead to disease with recessive inheritance. This results in an enrichment of specific Mendelian genetic diseases such as congenital nephrotic syndrome (Finnish type)^15^ or Northern epilepsy syndrome^16^ in certain areas of Finland today. Collectively, these currently 36 “founder diseases” are referred to as the “Finnish disease heritage”^10,17^ and show mostly autosomal recessive inheritance. Populations that underwent recent bottlenecks are also characterized by an enrichment of mildly deleterious variants, including variants causing disease with recessive inheritance, as those stochastically rose in frequency following a bottleneck event but did not yet drop to the lower levels by purging as in long-term large populations^3^. In line with that, potentially deleterious variant classes (such as protein truncating point mutations) are specifically enriched at an intermediate frequency range (i.e. ca. 0.5%-5%) in Finland^18,19^. An enrichment of founder diseases with recessive inheritance has also been observed in other isolated populations such as Tay Sachs or Gaucher in Ashkenazi Jews^20^, Hermansky– Pudlak syndrome in Puerto Ricans^21^ or ARSACS in French Canadians^22^.

Isolated populations have been successfully used to map disease genes for decades^6,11,23^ as the higher allele frequencies of founder variants increases statistical power for detecting disease associations and long shared haplotypes across families facilitated linkage disequilibrium mapping at lower map resolution. While Mendelian variants of high penetrance can segregate and thus be identified in family studies in non-bottlenecked populations, family studies have not proven ideal for finding disease genes in more complex and common diseases^3^ given the massive polygenicity and incomplete penetrance which demand profoundly larger sample sizes to support discovery. An additional power advantage when doing case control studies in isolated populations comes from the reduced variant diversity, which decreases noise from neutral genetic variants and also helps the identification of causal disease variants e.g. Finns have only 1/3 ultra-rare truncating variants as Non-Finnish Europeans^19^. On the other hand, statistical finemapping to identify causal variants^24^ may be more difficult in populations with recent bottlenecks due to longer extent of linkage disequilibrium. Some relatively common European Mendelian disease variants in e.g. CFTR causing cystic fibrosis^25^ or BRCA1/BRCA2 causing breast cancer are at much lower frequencies in Finland. On the other hand, greater haplotype sharing in the Finnish population facilitates imputation of lower frequency variants in array data from deep whole genome sequencing data with a population-specific reference panel down to frequencies below 0.0005^26^. Genotypes can thus be generated with arrays thereby enabling the large scale of the FinnGen research project. Its participants are largely recruited via hospital biobanks and are enriched for individuals with diseases across the clinical spectrum. The phenotypes are derived from national healthcare registry data collected over more than 50 years.

In this manuscript, we analyze the effects of 82,516 coding variants on 2,444 disease endpoints using nationwide EHR data of 176,899 Finns (FinnGen data freeze 4, 08/2019). In addition to the standard additive GWAS model^27^, we systematically search for homozygous effects. This enables us to explore two related questions of interest to both Mendelian and quantitative genetics communities. Firstly, we investigate the potential benefit of searching for homozygous effects in GWAS. Secondly, we explore the broad phenotypic consequences of mono-versus bi-allelic states of rare coding variants previously documented with a specific mode of inheritance in Mendelian disease.

#### Variants causing disease with recessive inheritance are at higher MAF in Finnish than Non-Finnish Europeans

In light of a global enrichment of deleterious variants in Finland^18,19^ and the well-described “Finnish Disease Heritage”^10,17^, we hypothesized that a set of known disease-causing variants should exist at higher frequencies in Finland. We thus compared minor allele frequencies (MAF) of 2419 unique variants that were listed in the disease variant database ClinVar^28^ (version 2018) as pathogenic or likely pathogenic (=P/LP) in genes with only recessive inheritance annotated in the Online Mendelian Inheritance in Man, OMIM® (omim.org) between 10,824 Finnish (FIN) and 10,824 Non-Finnish Europeans (NFE) taken from the gnomAD population database^29^ (Figure 1). Concordant with a known lower rare variant diversity^10^, only 554 P/LP variants were present in FIN while 2169 were present in NFE at same sample sizes. As expected, however, observed P/LP variants were at higher MAF in FIN (median MAF 9.2×10^−5^) than NFE (median MAF = 4.7×10^−5^, p-value = 6×10^−7^, Wilcoxon rank test), with the difference particularly pronounced in 133 variants in Finnish disease heritage genes^17^ (see Supplementary Figure S1). In FIN, 12% (67/554) of P/LP variants were above MAF 0.001 (39% [217/554] above MAF 10^−4^) compared to 2.5% (54/2169) in NFE (26% [559/2169] above MAF 10^−4^).

**Figure 1.**
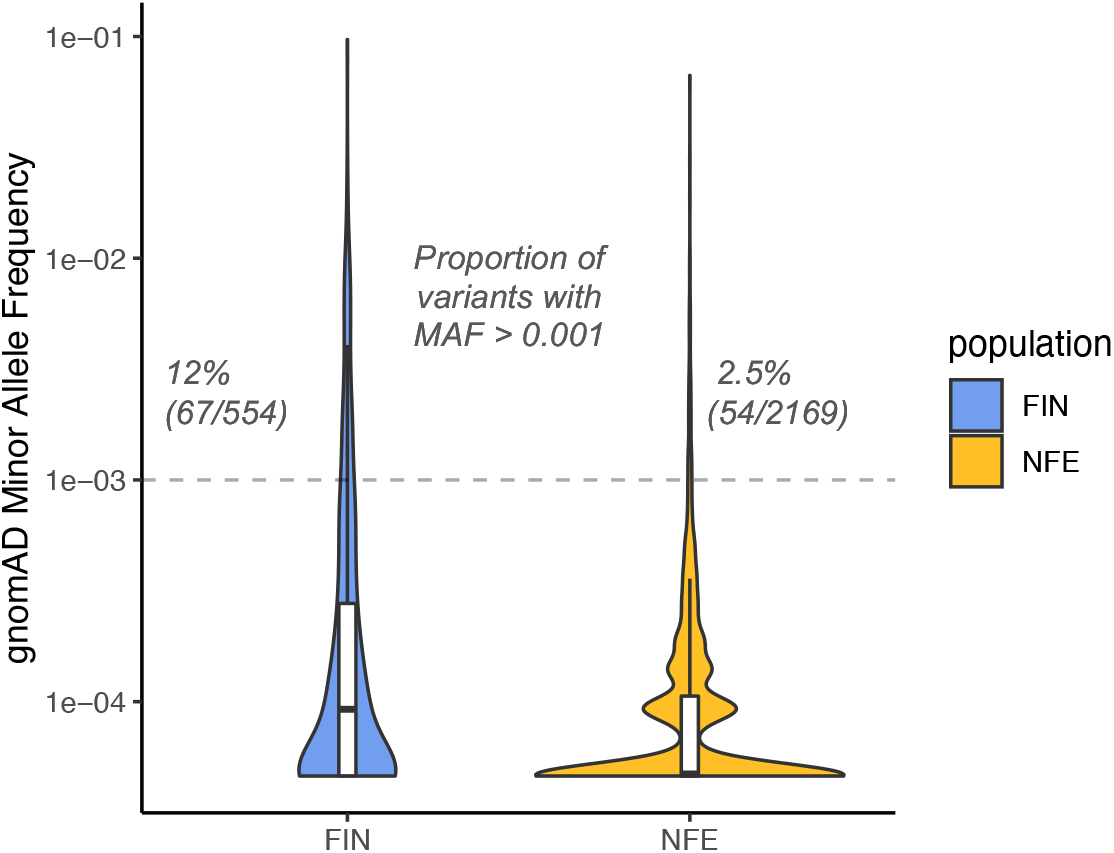
Variants known to cause disease with recessive inheritance are at higher MAF in Finnish than non-Finnish Europeans. We show MAF of 2419 unique P/LP variants (source: ClinVar^28^) in 10,824 individuals from FIN and 10,824 from NFE populations (source: gnomAD^29^). Violin plots are scaled to have the same area. Boxplots within violins show the 1st, 2nd and 3rd quartiles of the MAF distribution; whiskers maximally extend to 1.5 interquartile range.

#### Disease effects of variants known to (not) cause disease in 176,899 Finns

With such a large fraction of these alleles common enough to explore in genotype-based biobank studies, we examined 15,140 variants from ClinVar^28^ in 176,899 Finns from the FinnGen research project. These included 311 coding variants that were annotated as P/LP. We replicate known associations of the most frequent P/LP variants largely as expected (see Supplementary Note 1, Supplementary Figure S2), demonstrating such studies are powered to find disease associations across a wide range of phenotypes and inheritance models – noting as examples variants in *CHEK2, JAK2, OCA2* and *MC1R* that showed semidominant effects in different cancer phenotypes (Supplementary Figure S2 F,H,I,V,W,AC,AD,AE). In addition, we provide examples where these data can help verify or falsify previously described disease associations. We provide longitudinal disease onset data of 26 variants, labeled as P/LP in ClinVar by at least one submitter, with genome-wide significant associations in FinnGen (Supplementary Figure S2). We then investigated global effects of known ClinVar variants in a phenome-wide association analysis (pheWAS) of 2,444 disease phenotypes derived from health registry data using SAiGE^27^ cognizant of the fact that for many rare variants we may only be powered to identify their disease effects with moderate and not genome-wide levels of significance. To characterize the broader impact of these variants, we compared the ClinVar variants to randomly sampled intergenic variants in 15 MAF bins and in the same 3Mb windows using different p-value thresholds. As anticipated, we found significantly more phenotype associations than expected at all p-value thresholds for variants that were labeled as P/LP in ClinVar in genes described to cause disease with dominant inheritance (classification: OMIM). We also found a global association with disease phenotypes for variants listed as benign or likely benign (B/LB) in ClinVar regarded as “not implicated in monogenic disease”^30^ and often considered neutral^31^. 16 B/LB variants were even the most probable causal SNP of a GWAS locus following statistical finemapping^24^ (see Table 1). The ClinVar annotation labeling these variants as B/LB was above average quality (1.6; 7 of the 16 B/LB top causal variants had a one star and 9 of 16 a two star review status in ClinVar) compared to an average 1.2 stars for all B/LB variants in ClinVar (range: zero to three stars). As possible explanations for these phenotype associations, we found, that B/LB variants have high MAF, are more often protective and are more likely disease-associated than random coding variants as they are defined to be in genes that were already associated with disease (see Supplementary Note 2). An example is a protective B/LB probably causal (posterior inclusion probability [PIP] 0.99) variant we observed in the gene *DBH. DBH* is a gene associated with dopamine beta hydroxylase deficiency with recessive inheritance. This disease is characterized by severe hypotension^32^. We observed a missense variant in *DBH* that conveys protection from hypertension (p-value 5×10^−13^, beta -0.17, see Figure 3A), a plausible finding given *DBH*’s association with hypotension. The variant is also an example for a Finnish enriched variant (22 times higher AF than in non Finnish-Swedish-Estonian Europeans with an AF 0.05 in Finland).

**Table 1.**
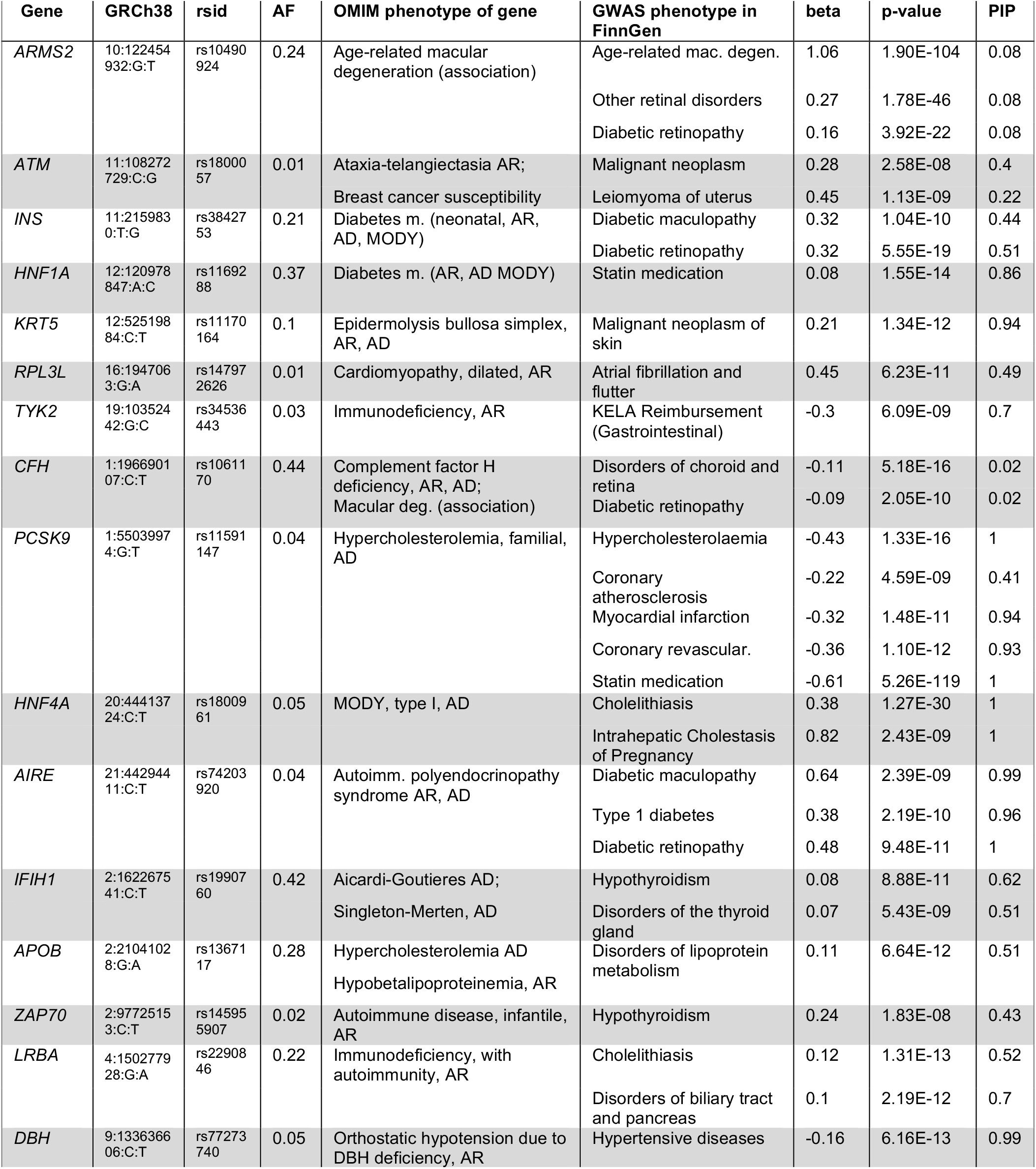
B/LB ClinVar variants that are the most likely causative (top PIP) variant in one GWAS locus. Genomic locations are given as rsids and GRCh38 coordinates (chromosome, position in bp, reference and alternate allele, separated by “:”). PIP, posterior inclusion probability (statistical finemapping^24^); AF, allele frequency in FinnGen; AD, autosomal dominant; AR autosomal recessive; KELA, Finnish Social Insurance Institution. All variants listed in Table 1 are missense variants.

**Figure 2.**
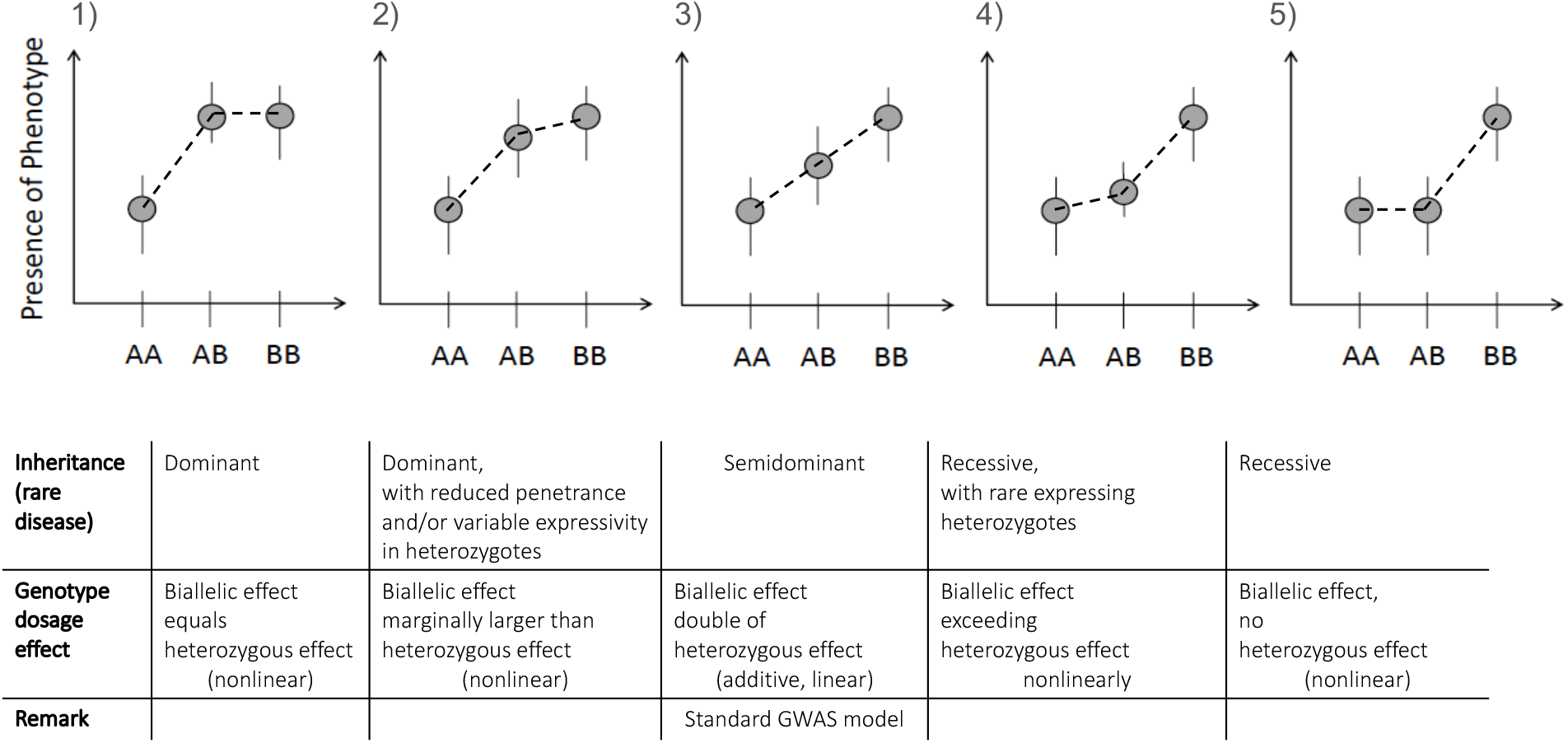
Schema of different effect sizes of heterozygous (mono-allelic) vs. bi-allelic variant states. A is the wildtype, B is the mutant allele. We distinguish five main scenarios that are associated with different inheritance modes used in rare disease genetics (table’s first row). In rare disease genetics, the phenotypes associated with the mono- and bi-allelic state in scenarios 2), 3) and 4) are usually viewed as distinct disease entities, with the mono-allelic phenotype regarded as dominantly inherited and the usually more severe bi-allelic phenotype regarded as recessively inherited. In the schema we don’t show overdominant/underdominant inheritance (rare outside the HLA region) and focus on autosomal inheritance.

**Figure 3.**
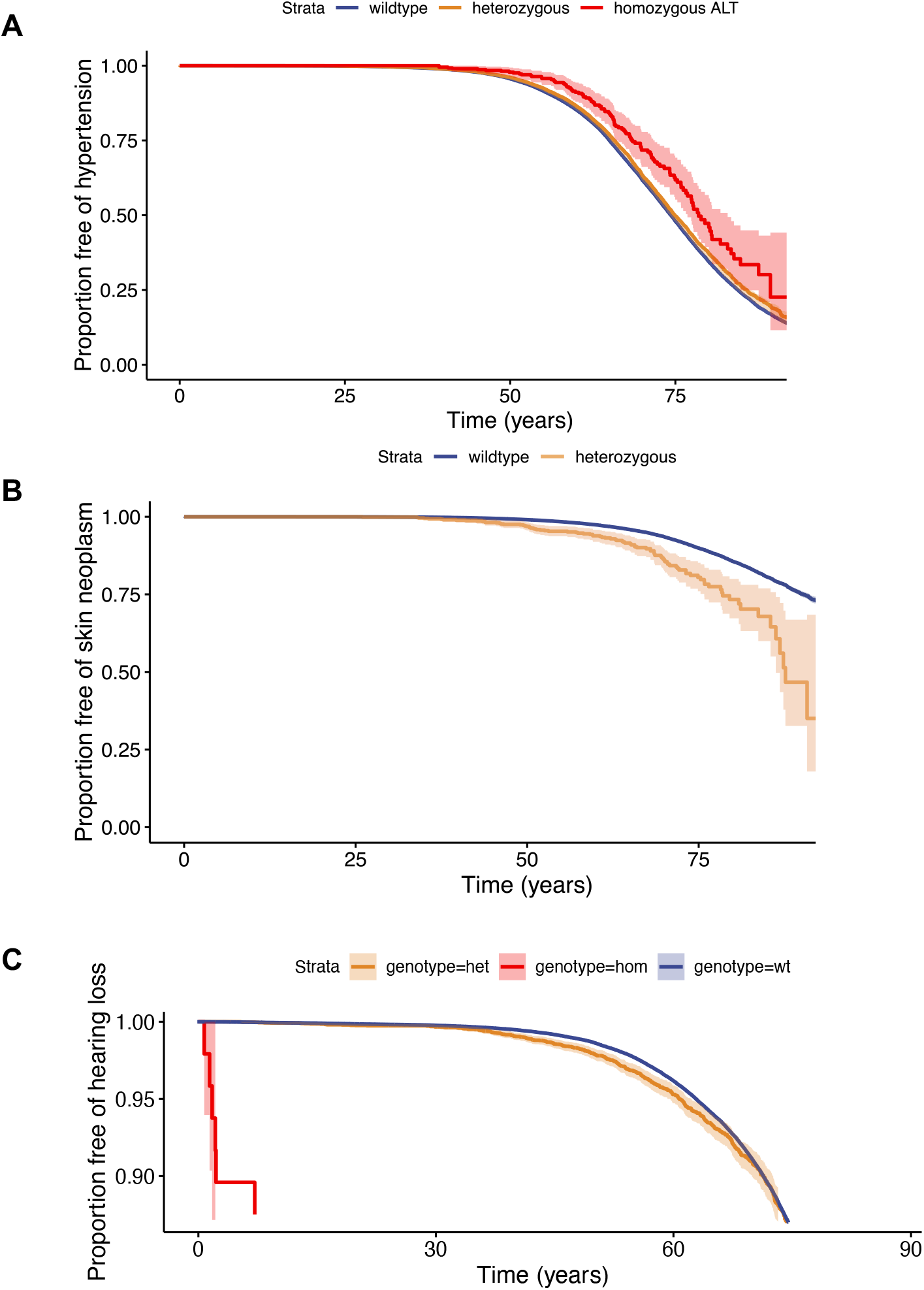
Age at first diagnosis of coding variant versus wildtypes (survival plots). **A)** B/LB missense variant in the gene *DBH* protects from hypertension. (*DBH* is associated with the recessively inherited disease dopamine beta-hydroxylase deficiency which is characterized by severe hypotension^32^), **B**) Known pathogenic variant in *XPA* is associated with skin cancer. In homozygous state that variant causes Xeroderma pigmentosum with childhood-onset skin cancer^33^. **C**) Known pathogenic variant (known recessive inheritance) in *GJB2* associated with hearing loss also in heterozygous state. The y-axis is cut at 0.9 for clarity.

#### Heterozygous effects of variants with known recessive inheritance

We found a global phenotype association signal for P/LP variants in disease genes with recessive inheritance (see Supplementary Figure S3). This could suggest some variants with recessive inheritance may have disease effects in a heterozygous state or simply that the additive model is detecting their known homozygous effects. To distinguish these possibilities, we set out to explore if heterozygous effects contributed to the global phenotype associations of P/LP variants in genes with recessive inheritance. First, we performed simulations of genotype and phenotype data (see Supplementary Note 3) and compared how well additive and recessive models captured simulated large homozygous effects with and without heterozygous effects. When no heterozygous effect was present (i.e., a traditional recessive model), an additive model did not capture rare Mendelian homozygous effects well in 1,000 simulations (median p-value 0.20, genome-wide significance in 1/1000) while the recessive model did (median p-value 1×10e-10, genome-wide significance in 912/1000 simulations). However, with increasing heterozygous effect sizes, the additive model’s significance increased (see Supplementary Figure S4). It is therefore likely that heterozygous effects contributed to our global phenotype association of variants with recessive inheritance. To confirm empirically, we repeated our global disease association after excluding 76 variants with homozygotes from our dataset of 311 variants. While reducing the number of variants expectedly reduced the signal, we still observed significantly (empirical p-value < 0.001) more disease associations with p-values < 10^−4^ than expectation. Overall, these results suggest multiple variants with known recessive inheritance have modest heterozygous effects. We further explored heterozygous effects in five P/LP variants in genes *SERPINA1, XPA, NPHS1, EYS* and *CLRN1* that were associated with diseases with p < 5×10^−8^. Variants in *SERPINA1, XPA* and *NPHS1* still had nominally significant effects after removing homozygotes from the GWAS. The GWAS’ phenotypes were similar to the known recessively inherited OMIM phenotypes (*SERPINA1* –Emphysema, *XPA* – skin cancer, NPHS1 – nephrotic syndrome). For variants in genes *SERPINA1* and *NPHS1* we found that heterozygotes also had significantly earlier disease onset than wildtypes (p-value 0.05, corrected for 5 tests, method: Wilcoxon-rank test). Expecting more heterozygous effects with increasing sample sizes, we found a heterozygous effect of a truncating variant in *GJB2* on hearing loss (Figure 3C, p=0.02, beta=0.11) with a larger FinnGen dataset (R7, n=309,154). We would like to highlight the gene *XPA* where homozygous loss-of-function variants are known to cause the disease Xeroderma pigmentosum, a condition with extreme vulnerability to UV radiation and childhood-onset skin cancer^33^. (No homozygotes were present in FinnGen.) We found that a known pathogenic pLoF^29^ (probable loss-of-function) variant in this gene was associated with adult-onset non-melanoma skin cancer in FinnGen in heterozygous state (p=8×10^−11^, beta=1.24 corresponding to odds ratio=3.5). For longitudinal data see Figure 3B. The same variant was previously associated with basal cell carcinoma in Japan with similar effect size^34^ (odds ratio=3.08, p-value = 0.0097). Thus, here we unequivocally confirm heterozygosity for *XPA* LoF is a significant risk factor for cancer and, given the high frequency of this variant in Finns, could be valuable personal risk information.

Widening our search to variants that were described as P/LP by at least one but not all submitters in ClinVar and variants in genes with recessive AND dominant inheritance in OMIM, we highlight a variant in *SCN5A* associated with severe arrhythmia disorders like sick sinus syndrome^35^ in homozygous (or compound heterozygous) state, which is confirmed in our dataset (Fisher’s Exact test, p-value: 9×10^−4^,OR: 48 (95%-CI 6-319)). The same *SCN5A* variant however protected from arrhythmia diseases in FinnGen (beta -0.48, p 2×10^−8^; PIP 0.996 after finemapping^24^ indicating it is the likely causal variant). This included atrial fibrillation (beta -0.62, p 7×10^−7^) and could be replicated in the UKBB^36^ (beta -0.39, p 0.04).

In summary, we find global as well as individual association signals of variants previously described to cause disease with recessive inheritance, semidominant inheritance in tumor suppressor genes and disease associations of variants labelled B/LB. Our data thus indicate the need for a nomenclature that better appreciates more complex inheritance of Mendelian diseases. We outline a suggestion in Figure 2.

#### 20 homozygous disease associations, of which 13 not found with additive models

To find potentially novel homozygous disease associations, we performed a pheWAS with additive and recessive models of 82,647 coding variants in FinnGen (2,634 pLoF, 76,884 missense, 3,129 others, INFO > 0.8) on 2,444 disease endpoints in 176,899 Finns using SAiGE^27^. After excluding the HLA region (chr6:25Mb-35Mb), we found 1780 additive associations with p-value < 5×10^−8^ for 443 coding variants in 303 genes. (For further information on these results see the FinnGen flagship paper, browse publicly released summary statistics at http://r4.finngen.fi/ or download at https://www.finngen.fi/en/access_results.) We then searched for homozygous effects across all coding variants comparing homozygous with heterozygous+wildtype alleles. We identified 124 associations - involving 39 unique variants - with a genome-wide significant p-value in the recessive GWAS model that was two orders of magnitude lower than the additive GWAS p-value (Figure 4A). Considering adjacent variants with r^2^>0.25 associated with the same (parent) trait as one locus, these associations corresponded to 31 unique loci (Figure 4B, Supplementary Table S2, Supplementary Figure S5). Genes *GJB2* and *EYS* each harbored two P/LP variants that were each significantly associated in the recessive model with hearing loss (variants in GJB2) or retinal dystrophy (variants in *EYS*). So we assume that here multiple disease-causing variants are causally associated at the respective loci. We confirmed this with comp-het effects for the P/LP variants in GJB2 (see Figure 5C) and additionally found comp-het effects in *CERKL* (see Supplementary Note 4). We next searched how many of the homozygous associations we identified were already known. 13 out of the 31 lead variants were in known OMIM genes (Figure 4B) of which all 13/13 genes were previously described with recessive disease inheritance in OMIM with the phenotype matching the known OMIM phenotype in 12/13 genes. 9/13 of variants in OMIM genes were previously reported disease-causing in ClinVar^28^ (6 [likely] P/LP, 3 conflicting). As expected, known Mendelian disease variants had large effect sizes with beta > 4 (corresponds to an odds ratio of 55). Of the 22 variants that were not previously reported disease-causing in ClinVar, 13/22 had Mendelian effect sizes with homozygous beta > 4 and were rare with MAF < 0.05. They included novel disease genes such as *CASP7* and *EBAG9* for which longitudinal disease onset data are shown in Figure 5 and discussed in Supplementary Note 5. In addition, we find that homozygotes of the *EBAG9* variant have fewer children and a later age at first child than wildtypes (Supplementary Figure S6). The association of *C10orf90* with hearing loss was published during the revision of this manuscript^37^. We thus provide a well-powered replication and additional information on disease onset (Figure 5). Our six most common variants with homozygous effects (MAF > 0.1) overlapped known associations in the GWAS catalog (Supplementary Table S3, variants in LD with r^2^>0.1^38^). For 4/6 of those variants, their phenotype in FinnGen matched the associated known GWAS hit. In addition, the variant in gene *TMEM214* that we found to be protective against pain (including limb, neck, head pain) in FinnGen in a homozygous state was in LD (r^2^ 0.78) with a GWAS hit for CRP level. It seems biologically plausible that CRP as a standard biomarker for systemic inflammation may be associated with unspecified pain.

**Figure 4.**
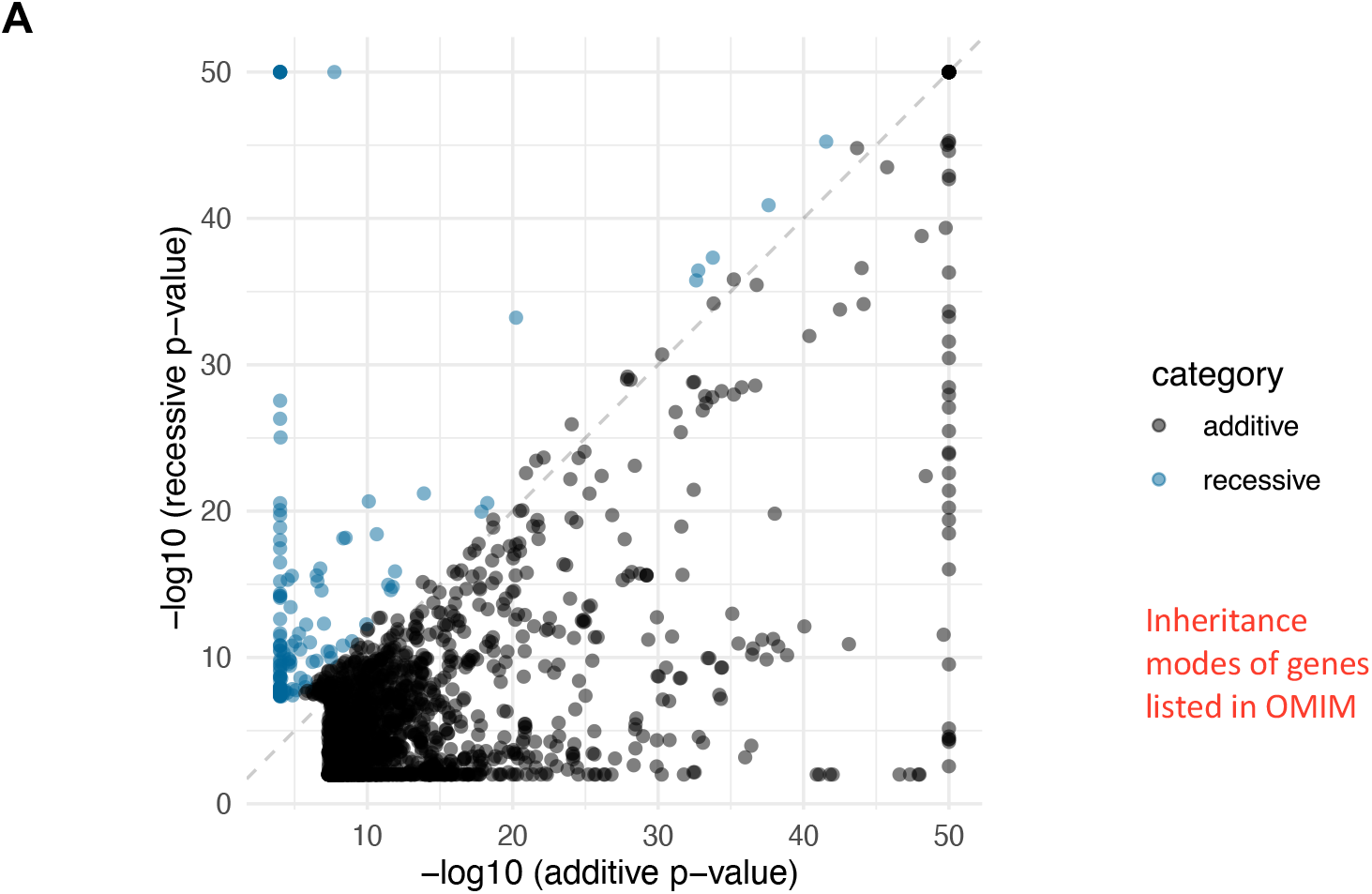
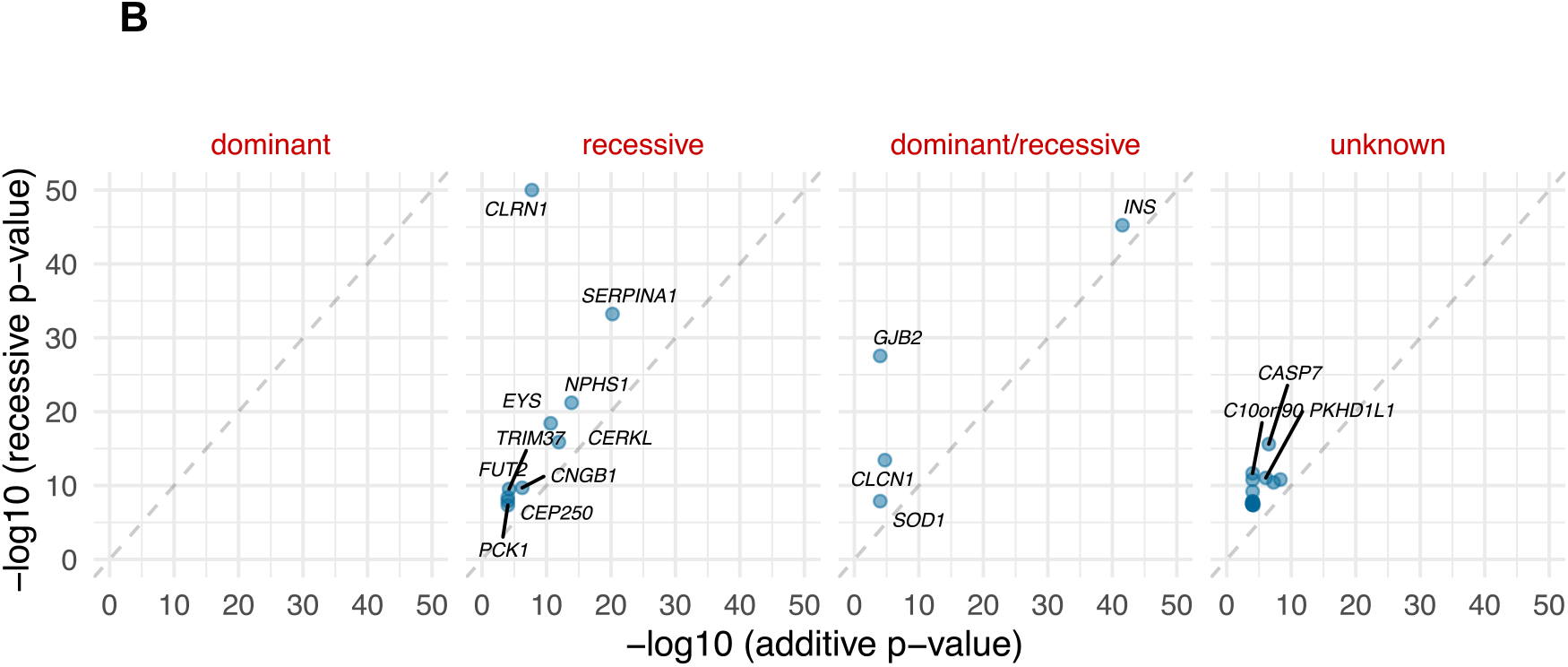
P-values of the additive vs. recessive GWAS model of all genome-wide significant variant-phenotype associations. Associations with recessive model’s p-value two orders of magnitude lower than additive model’s p-value are shaded in blue (=category “recessive”), all other associations are shaded in black. **A**) All associations. **B**) Recessive associations, independent loci, broken down by listed inheritance modes of the respective Mendelian genes in OMIM. For clarity, -log10 p-values are capped at 50.

**Figure 5.**
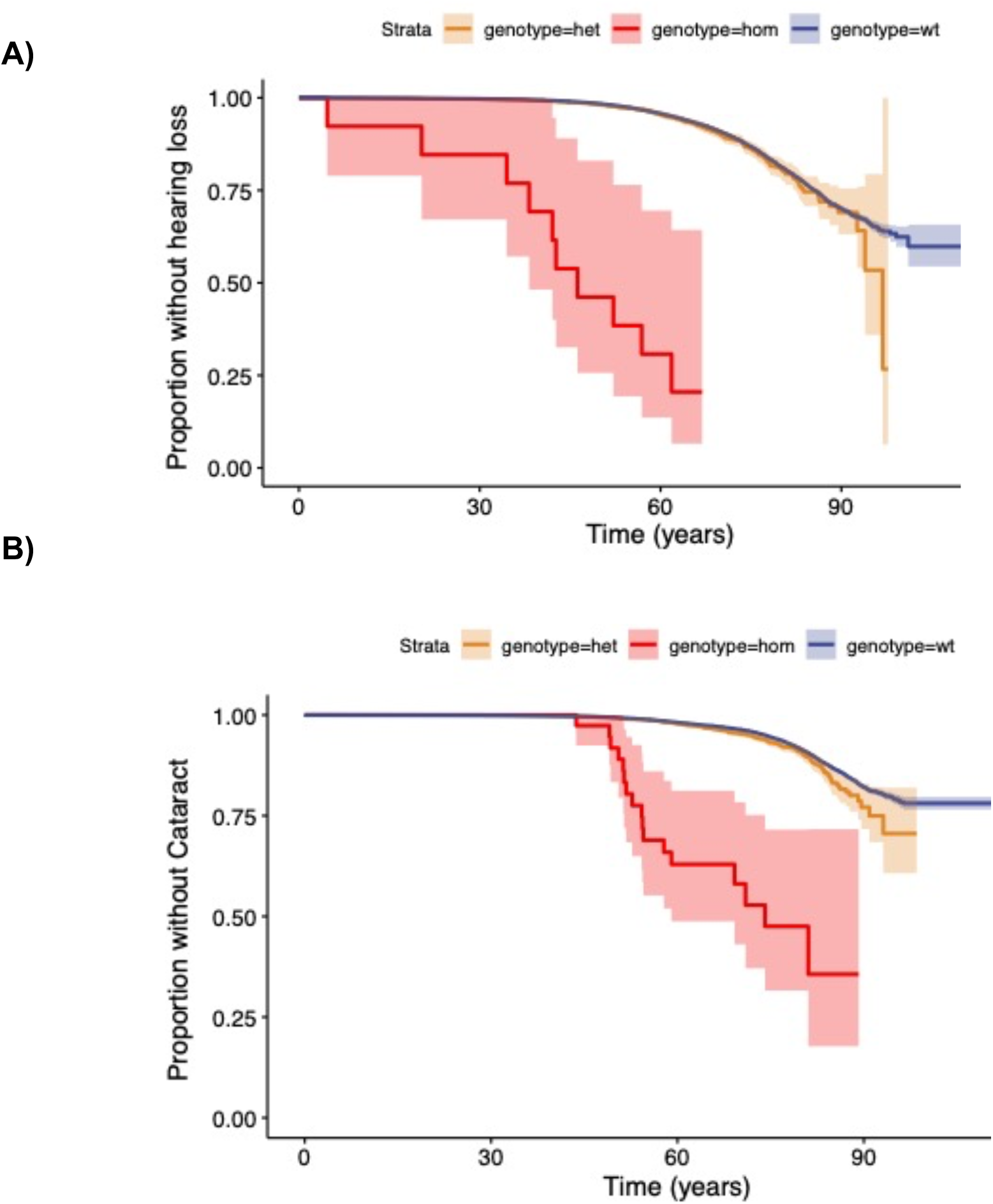
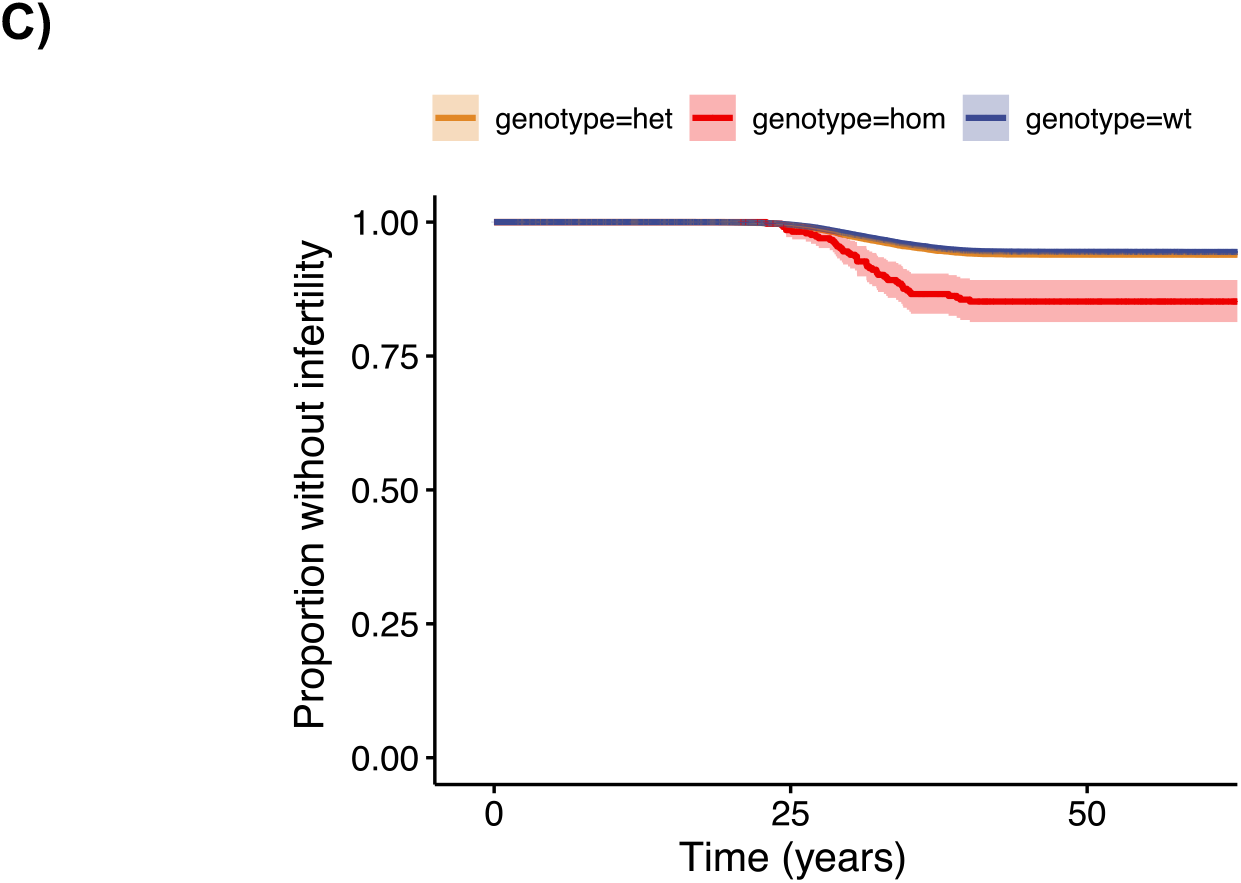
Age at first disease diagnosis of variant versus wildtypes (survival plots). Wt, wildtype; het, heterozygous; hom, homozygous. **A**) Missense variant in *C10orf90* associated with hearing loss, only recently described^37^, n=176,899. **B**) Missense variant in *CASP7* associated with cataract, not previously described, n=176,899. **C**) Intronic variant in *EBAG* associated with female infertility, not previously described, n=110,361 females.

We next sought to validate any homozygous associations in the UK biobank^36^ and in the FinnGen datafreeze R6. Owing to the Finnish population’s isolation^10^ (Figure 1, introduction), most of our variants with homozygous effects were Finnish-enriched with the exception of variants in genes *GJB2, SERPINA1* and *C15orf40* being >2 fold higher in Finnish than non-Finnish-Swedish-Estonian Europeans in gnomAD. In total, 13/31 variants had >= 5 homozygotes and partially matching phenotypes in UKBB suitable for validation. 8/13 of those had significant homozygous associations (p-value < 0.05) to related phenotypes in the UKBB (Supplementary Figure S7, Supplementary Table S4). For 3 variants with < 5 homozygotes in UKBB, we found homozygous associations (p-value < 0.05) in other coding variants in the UKBB. In addition, 8 variants had additive associations (p-value < 0.05) in the UKBB (Supplementary Table S5). 18/31 variants stayed genome-wide significant in FinnGen datafreeze R6 resulting in a total of 20 homozygous associations validated in FinnGen R6 and/or UKBB (Table 2). These included 4/20 novel associations. Homozygous associations that could not be validated are listed in Supplementary Table S2. The majority of homozygotes also had substantially earlier disease onset than wildtypes when affected with the same diseases, which we interpret as additional evidence for genetic variants’ effect on disease (Supplementary Figure S8).

**Table 2.**
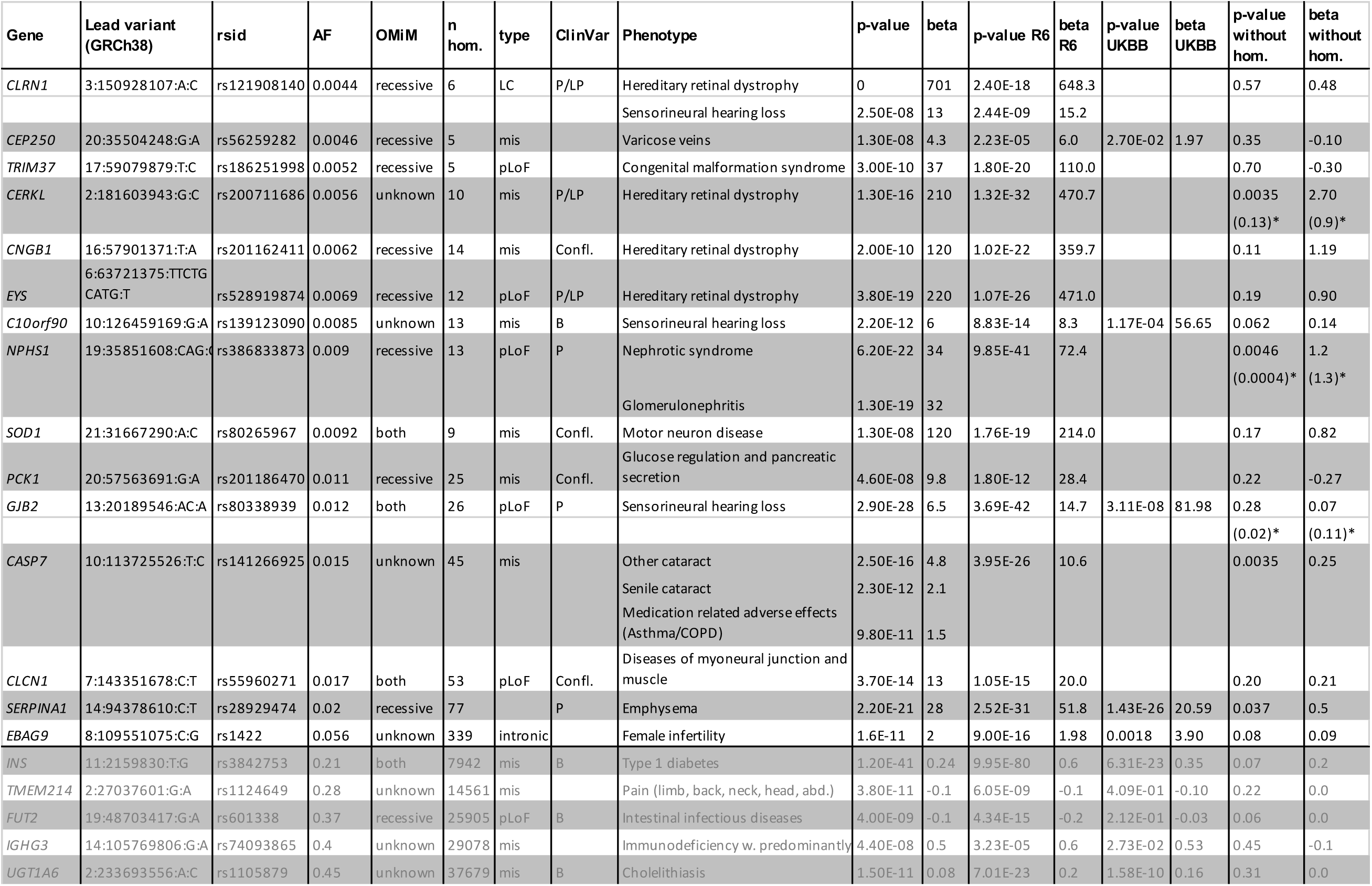
20 homozygous associations. mis, missense; pLoF, probable loss-of-function; AF, allele frequency in FinnGen; hom, homozygotes in FinnGen; R6, FinnGen data release 6 (n=234,553); UKBB, UK biobank (n=420,531). Adjacent variants with r^2^>0.25 associated with the same (parent) trait are considered as one locus and only the most significant variant shown per locus. Lead variants are given as rsids and GRCh38 coordinates (chromosome, position in bp, reference and alternate allele, separated by “:”). The p-value and beta in ()* refers to the repetition of the heterozygous test in R6 or R7 data (n=309,154, only *GJB2*) after exclusion of comp-het carriers. *EBAG9* is the only noncoding variant presented here, as we estimate it is a more likely causal variant than the originally identified pLoF in *PKHD1L1* (Supplementary Note 5). Variants at the bottom shaded in grey overlap known GWAS hits loci that are listed in Supplementary Table S3.

We next investigated if any of the variants more significant in the recessive than additive model also had subtle heterozygous effects on the same disease endpoints by excluding homozygotes from GWAS. Four variants had nominally significant heterozygous effects. These included variants in *SERPINA1* (phenotype: Emphysema, p-value: 0.037, beta: 0.5), *NPHS1* (phenotype: nephrotic syndrome, p-value: 0.0046, beta: 1.2), *CERKL* (phenotype: retinal dystrophy, p-value: 0. 0035, beta: 2.7) and *CASP7* (phenotype: adult-onset cataract, p-value: 0.0035, beta: 0.25). However, when excluding individuals with comp-het variants from the analysis, the association for *CERKL* dropped to p=0.14, beta 1.4 indicating the heterozygous effect was driven by comp-hets while the association in *NPHS1* stayed significant (p-value 0.0004, beta 1.3). We thus classify inheritance of variants in *NPHS1, CASP7* and *SERPINA1* as recessive, with rare expressing heterozygotes (schema: Figure 2).

13/20 of our validated homozygous associations were not genome-wide significant in the additive GWAS model (Supplementary Table 2). In simulations, an additive model found a homozygous effect (10x higher disease probability in homozygotes, MAF 0.01 in n=200,000) in 1/1000 simulations while the recessive model found it in 912/1000 simulations (Supplementary Note 3). This illustrates how using a recessive GWAS model may uncover associations that might otherwise be missed (Supplementary Figure S4).

## Discussion

A subset of disease-causing variants are enriched to unusually high frequencies in populations with a history of recent bottlenecks such as Finland. This applies particularly to homozygous variants causing disease with recessive inheritance. The FinnGen cohort, given its size and broad medical phenotypes over the lifespan, is therefore well-powered to study both the effects of Mendelian mutations with known recessive inheritance in heterozygotes and to discover novel alleles with an outsized effect in homozygotes.

We first studied effects of variants previously reported in human disease in ClinVar^28^ in > 170k participants of FinnGen. Here, we found that multiple variants previously described as B/LB had effects on disease phenotypes which is noteworthy as a reminder that variants labeled as benign, even if they do not cause monogenic disease, should not be presumed to be neutral^31^ or without phenotypic consequence (see Supplementary Note 2). We then found genome-wide significant effects of 26 known disease-causing ClinVar variants including semidominant effects in tumor suppressor genes *CHEK2, JAK2, OCA2* and *MC1R* that were in part previously described^39-42^. We also discovered that a heterozygous truncating variant in the tumor suppressor and DNA-repair gene *XPA* increased susceptibility to adult-onset skin cancer with an effect far stronger than usually found by GWAS. This association was already shown with nominal significance (p=0.01) in a previous study^34^. Thus, we provide the first definitive evidence of this link in heterozygotes. Homozygous loss-of-function variants in the gene *XPA* are known to cause the disease Xeroderma pigmentosum (XP), a condition with extreme vulnerability to UV radiation and childhood-onset skin cancer^33^. Given the phenotypic relatedness of skin cancer and XP and the semidominant effects of variants in other tumor suppressor genes, this highlights that our way of describing their respective inheritance with only dominant and recessive labels does not adequately capture disease biology. For distinct molecular effects, it makes however biologically sense to view their inheritance separately. *SERPINA1* is an example where the Pi-Z variant causes different disease entities with distinct molecular effects, which we can also observe in our data. The Pi-Z variant causes a molecular gain-of-function of Alpha1-Antitrypsin (A1AT) which is not properly secreted and therefore accumulates in the liver resulting in diseases such as Cholelithiasis or Fibrosis. Improper secretion of A1AT however results in a deficiency at its target tissue, the lung, a loss-of-function effect that would only manifest if A1AT level is very low as in Pi-Z homozygotes. The inheritance of Pi-Z is dominant for the liver phenotype, but recessive for the lung phenotype (with a small heterozygous effect). *LDLR* associated with familial hypercholesterolemia (FH) is another example for semidominant inheritance where bi-allelic pathogenic variants result in more severe disease while single heterozygous pathogenic variants lead to a milder form of FH. On OMIM, however, only recessive and dominant inheritance modes are given for FH associated with *LDLR*. Terms beyond recessive/dominant such as semidominant inheritance are rarely used in clinical genetics^43^ even though recommended in ClinGen’s gene curation SOP. While it has been estimated that semidominant effects are much more common than true dominant effects^44^, phenotypes of heterozygotes/homozygotes are frequently unknown. Semidominant inheritance is more frequently described in animal and plant genetics^45^, where mono- and bi-allelic effects can be more easily systematically studied. In addition to a more accurate description of biology, it could benefit clinical variant interpretation if evidence for pathogenicity was aggregated across bi-allelic and mono-allelic observations and not viewed as separate disease entities.

Adding to an increased complexity of Mendelian variants’ inheritance, we also detect a global heterozygous effect on disease in variants known to cause disease with recessive inheritance. This is in line with previous studies that found subtle disease and/or fitness effects of heterozygous variants in genes described to cause disease with recessive inheritance in mice^46^, drosophila^47^ but also in humans^48^. Individual examples include the already-mentioned variant in *XPA* known to cause XP in a homozygous state^33^, that increased risk of skin cancer while in a heterozygous state. We also find heterozygous effects of variants with recessive inheritance in *NPHS1* on kidney disease, which was previously debated^49^, in *SERPINA1* on emphysema^50^ and GJB2 on hearing loss, previously hypothesized^51^. As expected for recessive inheritance, the variants had an order of magnitude larger effect size and often earlier disease onset in the homozygous than in heterozygous state thereby exceeding a linear additive model. With increasing sample sizes, we identify more heterozygous effects indicating this could be a more widespread phenomenon (e.g. in *GJB2* we find the effect on hearing loss only with a larger dataset [n=309,154]). Of course, it is possible that some heterozygous effects come from low-frequency variants in comp-het state (as we observe for *CERKL*) that were not captured by our genotyping array. However, a different age of disease onset of heterozygotes than homozygotes suggests that scenario is unlikely. In addition, with population specific exome sequencing and imputation we could account for the presence of additional pathogenic variants at frequencies >0.2%. Another limitation of our approach is the lack of more in-depth phenotypes such as serological or diagnostic tests. We thus likely miss subtle physiological differences between heterozygotes and wildtypes. This should, however, not affect our main conclusions. Long before these large-scale data became available, small heterozygous effects were found in variants causing Mendelian disease with recessive inheritance^52^, in some cases giving an advantage against certain infectious diseases^53,54,55^. Similarly, we find that one variant in *SCN5A*, which was previously associated with severe arrhythmia disorders like sick sinus syndrome^35^ in a bi-allelic state, protected from mild arrhythmia diseases including atrial fibrillation in a heterozygous state in FinnGen. Previous experimental data found a mild loss-of-function effect of this variant^35,56^. This is in line with some ECG times being longer in a few SCN5A heterozygotes^35^ pointing to a potential slowing of electrical conduction in the heart outlining a possible protective mechanism against cardiac arrhythmia such as atrial fibrillation. Further studies are needed to elucidate the mechanism comprehensively.

We systematically investigated coding variants’ homozygous effects genome-wide with pheWAS at biobank-scale. We could validate 20 loci (4 of them novel) with large bi-allelic effects without, or with only nominally significant, heterozygous effects. Known associations (e.g. variants in *FUT2* protecting from intestinal infection^57^) served as positive controls. Novel associations include complex non-syndromic diseases with no/few previously described large-effect variants like adult-onset cataract (new disease gene: *CASP7*) and female infertility (new disease gene: *EBAG9*). Intuitively, these novel findings appear in phenotypes (deafness, cataracts, infertility) for which Mendelian subtypes would not obviously be clinically distinguished from other common presentations, though as in past examples earlier age of onset in these cases is notably seen. Bi-allelic associations of rare coding variants were found in other population biobanks^9,58-60^ though to our knowledge, not investigated at that broad phenotype scale.

In summary, our biobank-scale additive and homozygous pheWAS of coding variants demonstrate a benefit of homozygous scans in GWAS. We find known and novel bi-allelic associations across a broad spectrum of phenotypes such as retinal dystrophy, adult-onset cataract and female infertility that are missed by the standard additive GWAS model. As a related point, we find evidence of complex inheritance patterns of multiple Mendelian variants and highlight a more widespread presence of allelic series in clinically relevant genes than previously appreciated.

## Data Availability

All summary statistics described in this manuscript can be found in the Supplementary Material. Additional summary statistics of additive and recessive GWAS of FinnGen release 4 can be investigated in our web browser at http://r4.finngen.fi/ or downloaded at https://www.finngen.fi/en/access_results. A full list of FinnGen endpoints for release 4 is available at https://www.finngen.fi/en/researchers/clinical-endpoints.

https://www.finngen.fi/en/access_results

## Supplementary Material

### Supplementary Figures

Supplementary Figure S1. MAF of known disease-causing ClinVar variants in Finnish disease heritage genes in FIN and NFE gnomAD cohorts.

Supplementary Figure S2A-AG (separate pdf). Longitudinal survival curves showing disease onset of homozygous, heterozygous and wildtypes of likely P/LP or conflicting variants.

Supplementary Figure S3. Global disease associations of variant categories.

Supplementary Figure S4. Simulations of rare recessive and additive effects with varying sizes of heterozygous effects.

Supplementary Figure S5A-AE (separate pdf). Longitudinal survival curves showing disease onset of homozygous, heterozygous and wildtypes of variants with homozygous effects in FinnGen.

Supplementary Figure S6. EBAG is associated with female infertility.

Supplementary Figure S7. Replications of homozygous associations in the UK biobank.

Supplementary Figure S8. Age at first diagnosis of homozygous, heterozygous and wildtypes of variants with homozygous effects in FinnGen.

Supplementary Figure S9. Age at first disease diagnosis of variant carriers in GJB2.

### Supplementary Tables

Supplementary Table 1. Most significant disease associations of all 15140 coding variants annotated in ClinVar.

Supplementary Table 2. Summary statistics of coding variants with any homozygous association with p-value < 10^−7^ (all available disease endpoints, associations with p-value < 0.01).

Supplementary Table 3. Variants with homozygous disease associations in FinnGen that overlap known GWAS loci.

Supplementary Table 4. Variants with homozygous disease associations in FinnGen that could be replicated in the UK biobank.

Supplementary Table 5. Variants with disease homozygous associations in FinnGen that could be tested for additive association in the UK biobank.

### Supplementary Notes

Supplementary Note 1. Known disease-causing variants in FinnGen.

Supplementary Note 2. B/LB variants in FinnGen.

Supplementary Note 3. Simulations of homozygous and additive effects.

Supplementary Note 4. Compound heterozygous effects in *GJB2* and *CERKL*.

Supplementary Note 5. Novel homozygous disease associations in *CASP7, C10orf90* and *EBAG9*.

List of FinnGen contributors.

## Acknowledgements

We would like to thank all FinnGen participants for their contributions to research. Patients and control subjects in FinnGen provided informed consent for biobank research, based on the Finnish Biobank Act. Research cohorts collected prior the Finnish Biobank Act were collected based on study-specific consents and later transferred to the Finnish biobanks after approval by Fimea, the National Supervisory Authority for Welfare and Health. The FinnGen project, data release 4, was funded by Business Finland and eleven industry partners. Please find further information on funding and approvals in the Methods section and the full list of members of the FinnGen consortium in the Supplementary Information. We thank Leslie Biesecker, Alfred L. George and Jack Kosmicki for helpful comments.

## Author contributions

H.O.H wrote the manuscript and generated the figures. H.O.H and J.K. performed analyses. K.J.K., S.M.L., A.S.H, W.Z., M.K. and H.L.R. contributed data and tools. A.P. and M.J.D. supervised the study. H.O.H and M.J.D. conceived the study. All authors listed under FinnGen contributed to the generation of the primary data of the FinnGen data release 4. All authors reviewed the manuscript.

## Methods

### Data availability

All summary statistics described in this manuscript can be found in the Supplementary Material. Additional summary statistics of additive and recessive GWAS of FinnGen release 4 can be investigated in our web browser at http://r4.finngen.fi/ or downloaded at https://www.finngen.fi/en/access_results. A full list of FinnGen endpoints for release 4 is available at https://www.finngen.fi/en/researchers/clinical-endpoints. We are currently submitting variants with novel associations or variants that clarified conflicting or wrong classifications to ClinVar.

### Code availability

Please see https://finngen.gitbook.io/documentation/ for a detailed description of data production and analysis including code used to run analyses. Please see https://github.com/FINNGEN/ for further code repositories used to run analyses in FinnGen. R code to reproduce figures are available upon request.

### Ethics and Data Access Approvals

The Coordinating Ethics Committee of the Hospital District of Helsinki and Uusimaa (HUS) approved the FinnGen study protocol Nr HUS/990/2017. The FinnGen project is approved by Finnish Institute for Health and Welfare (THL), approval number THL/2031/6.02.00/2017, amendments THL/1101/5.05.00/2017, THL/341/6.02.00/2018, THL/2222/6.02.00/2018, THL/283/6.02.00/2019), Digital and population data service agency VRK43431/2017-3, VRK/6909/2018-3, the Social Insurance Institution (KELA) KELA 58/522/2017, KELA 131/522/2018, KELA 70/522/2019 and Statistics Finland TK-53-1041-17. The Biobank Access Decisions for FinnGen samples and data utilized in FinnGen Data Freeze 4 include: THL Biobank BB2017_55, BB2017_111, BB2018_19, BB_2018_34, BB_2018_67, BB2018_71, BB2019_7 Finnish Red Cross Blood Service Biobank 7.12.2017, Helsinki Biobank HUS/359/2017, Auria Biobank AB17-5154, Biobank Borealis of Northern Finland_2017_1013, Biobank of Eastern Finland 1186/2018, Finnish Clinical Biobank Tampere MH0004, Central Finland Biobank 1-2017, and Terveystalo Biobank STB 2018001. UK biobank data was accessed under protocol 31063.

### Funding and Partners

The FinnGen project is funded by two grants from Business Finland (HUS 4685/31/2016 and UH 4386/31/2016) and eleven industry partners (AbbVie Inc, AstraZeneca UK Ltd, Biogen MA Inc, Celgene Corporation, Celgene International II Sàrl, Genentech Inc, Merck Sharp & Dohme Corp, Pfizer Inc., GlaxoSmithKline, Sanofi, Maze Therapeutics Inc., Janssen Biotech Inc). Following biobanks are acknowledged for delivering biobank samples to FinnGen: Auria Biobank (www.auria.fi/biopankki), THL Biobank (www.thl.fi/biobank), Helsinki Biobank (www.helsinginbiopankki.fi), Biobank Borealis of Northern Finland (https://www.ppshp.fi/Tutkimus-ja-opetus/Biopankki/Pages/Biobank-Borealis-briefly-in-English.aspx), Finnish Clinical Biobank Tampere (www.tays.fi/en-US/Research_and_development/Finnish_Clinical_Biobank_Tampere), Biobank of Eastern Finland (www.ita-suomenbiopankki.fi/en), Central Finland Biobank (www.ksshp.fi/fi-FI/Potilaalle/Biopankki), Finnish Red Cross Blood Service Biobank (www.veripalvelu.fi/verenluovutus/biopankkitoiminta) and Terveystalo Biobank (www.terveystalo.com/fi/Yritystietoa/Terveystalo-Biopankki/Biopankki/). All Finnish Biobanks are members of BBMRI.fi infrastructure (www.bbmri.fi).

### Coding variants in FinnGen, release 4

In summary, we investigated 82,647 coding variants in the FinnGen project, release 4 (2,634 pLoF, 76,884 missense, 3,129 others) in 176,899 Finns. In 110,361 individuals sex was imputed as female. We filtered to variants with INFO > 0.8 at a median MAF 0.004 (minimum MAF 3×10^−5^). Circa half of the samples came from existing legacy collections while the other half was from participants newly recruited to the FinnGen project. Samples were genotyped on custom microarrays and rare variants imputed using a population specific reference panel^26^. Please see the flagship paper or https://finngen.gitbook.io/documentation/ for a detailed description of data production and analysis. To calculate variant enrichment in Finns after a bottleneck event, we utilize as a general European reference point exomes from European samples in gnomAD 2.1.1 excluding those from Finland, Sweden and Estonia (so-called non-Finnish-Swedish-Estonian Europeans). Due to large-scale migrations from Finland to Sweden in the 20th century, a substantial fraction of the genetic ancestry in Sweden is of recent Finnish origin, and the linguistically (and geographically) close population of Estonia is likely to share elements of the same ancestral founder effect.

### Validation in FinnGen, release 6

We validated homozygous associations of the lead variants in FinnGen data release 6 consisting of genotype and EHR data from 234,553 Finns.

### GWAS searching for additive, homozygous and heterozygous effects

We performed GWAS on 2,444 disease endpoints investigating the effects of 82,647 coding variants with an additive and recessive model using the method SAiGE^27^. Covariates in FinnGen were Age, Sex, genotyping batch and the first 10 Principal Components of genotypes. To identify heterozygous effects we performed GWAS with an additive model after excluding homozygotess. In the recessive GWAS model we analyzed effects of homozygous alleles on disease phenotypes in comparison to wildtype+heterozygous alleles. The docker container finngen/saige:0.39.1.fg with all necessary software used to run SAiGE in additive or recessive mode can be found at the docker container library hub.docker.com. We replicated our identified 31 genome-wide significant homozygous associations in 420,531 individuals with European ancestry in the UK biobank using the same recessive model in SAiGE. GWAS covariates in UKBB were Age, Sex, Age x Sex, Age^2^, Age^2^ x Sex and the first 10 Principal Components of genotypes. The genotype and phenotype files along with ancestry definitions, phenotype definitions and SAiGE null models were taken from the PAN UKBB project and are further described here https://pan.ukbb.broadinstitute.org.

### Annotating variant effects from ClinVar

We annotated variants from release 03/25/2020 of ClinVar^28^. For any variant included in the main tables, we rechecked current classifications in ClinVar and OMIM on 11/02/2021. We grouped variants into categories according to their “ClinVar_ReviewStatus”. Our main categories were P/LP (= likely pathogenic or pathogenic, 311 variants), conflicting evidence (=at least one submitter labeled a variant as P/LP but at least one other submitter labeled it different from P/LP, 298 variants) and B/LB (= likely benign or benign, 10,948 variants). Other categories into which we grouped variants which we are not explicitly discussing in the manuscript were ‘association’ (26 variants), ‘drug response’ (59 variants), ‘not provided’ (141 variants), ‘protective’ (14 variants), ‘risk factor’ (74 variants) and ‘VUS’ (= variant of unknown significance, 3269 variants), see Supplementary Table 1.

### Annotating inheritance mode from OMIM

We downloaded the Online Mendelian Inheritance in Man (OMIM) catalog of human genetic diseases (www.omim.org) version 06/2019. From OMIM, we annotated genes reported to have variants that cause disease with a recessive inheritance mode such as NPHS1. Genes showing recessive as well as dominant inheritance modes with the same or different phenotypes were labeled as “both OMIM genes” such as GJB2.

### Global phenotype associations of ClinVar variant categories

We compared global disease phenotype associations of different ClinVar variant categories (B/LB, P/LP or conflicting variants in genes with dominant or recessive inheritance) with phenotype associations of random intergenic variants. For a given variant category, we counted how many variants had at least one significant GWAS hit (2,444 phenotypes) below a given p-value threshold. We then compared those to the number of top GWAS loci below the p-value threshold of 1000 random samples of intergenic variants. We calculated with empirical p-values if any ClinVar variant categories had significantly more disease associations than random intergenic variants below respective p-value thresholds. Minor allele frequency (MAF) influences the power to identify significant associations. Therefore we adjusted for MAF by sampling intergenic variants in 15 equal sized bins that corresponded to the MAF of the variants under investigation. To account for linkage disequilibrium we sampled intergenic variants from the same 3Mb windows as variants in the respective gene set.

### Age at first diagnosis

We compared age at first diagnosis of homozygotes/heterozygotes compared to wildtypes, respectively using Wilcoxon rank tests. For few compound heterozygous variants (as indicated in the manuscript), we also performed survival analyses using age at first disease diagnosis as outcome using a Cox proportional hazard model with the same covariates that were also used in the GWAS (sex, age, genotyping batch, first 10 principal components).

