## Supplementary Material for "Mono- and bi-allelic effects of coding variants on disease in 176,899 Finns"

### Supplementary Figures

**Supplementary Figure S1. Disease-causing variants are at higher frequency in Finns.** Known disease-causing (ClinVar (likely) pathogenic) variants in genes with recessive inheritance are at higher minor allele frequency in 10,824 Finnish (FIN) than 10,824 non-Finnish Europeans (NFE) from gnomAD[1]. A) All 2419 variants. In B) we subset to 133 variants in Finnish disease heritage genes[2,3].

**A** **B**

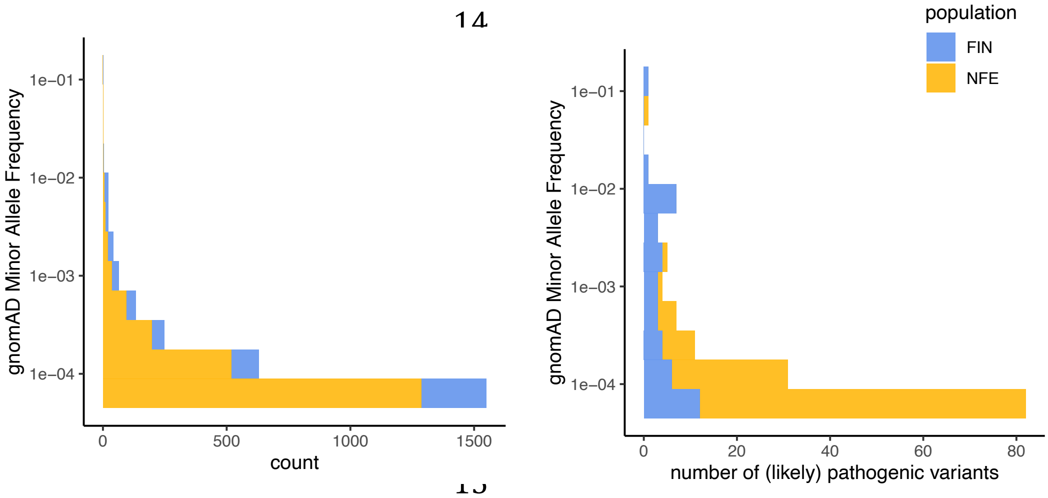

**Supplementary Figure S2.** Separate pdf. Longitudinal survival curves showing disease onset of homozygous, heterozygous and wildtypes of likely pathogenic or conflicting variants.

**Supplementary Figure S3. Global disease associations of variant categories.** Variants previously described as disease-causing (ClinVar (likely) pathogenic or conflicting variants[4]) but also ClinVar (likely) benign variants are globally associated with disease phenotypes compared with randomly sampled intergenic variants. Disease-causing variants in OMIM genes that were described with only dominant inheritance, as well as genes with only recessive inheritance were globally disease-associated.

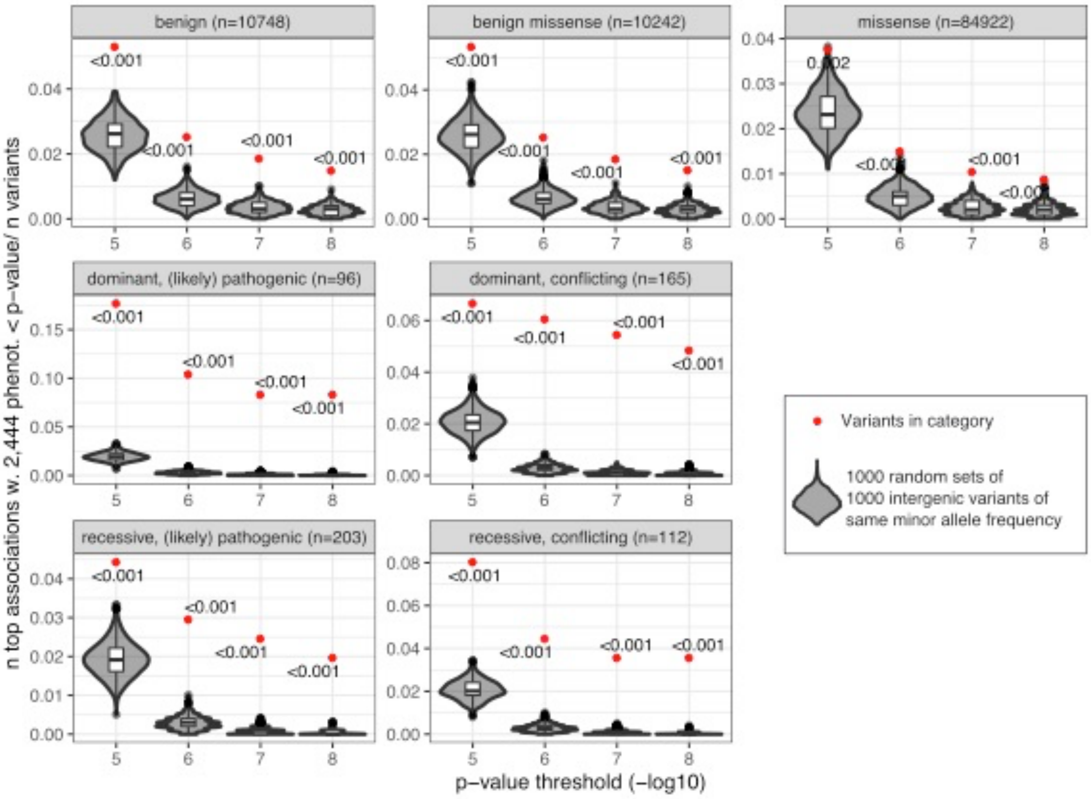


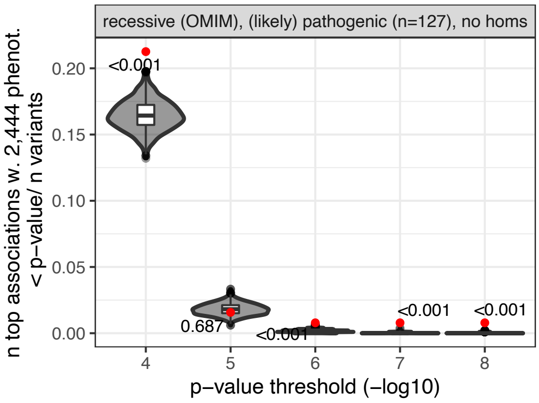

**Supplementary Figure S4. Simulations of rare recessive and additive effects with varying sizes of heterozygous effects.** Here, we generated genotype counts of wildtype, heterozygotes and mutant homozygotes in 200,000 individuals of a variant with an allele frequency of 0.01 following Hardy Weinberg Equilibrium and random controls and cases of a disease with a prevalence of 0.05. Simulations, where the additive model's p-value is < 5x10e-8 are colored red. In **A** we set the probability of homozygotes to develop the disease to 10x compared to wildtype. In this figure, we show the effect of different heterozygote effect sizes the significance (p-value) of an additive (y-axis) and a recessive (x-axis) logistic regression model. In **B** we set the homozygous disease effect to 2x and heterozygous disease effect to 1.5.

**A**

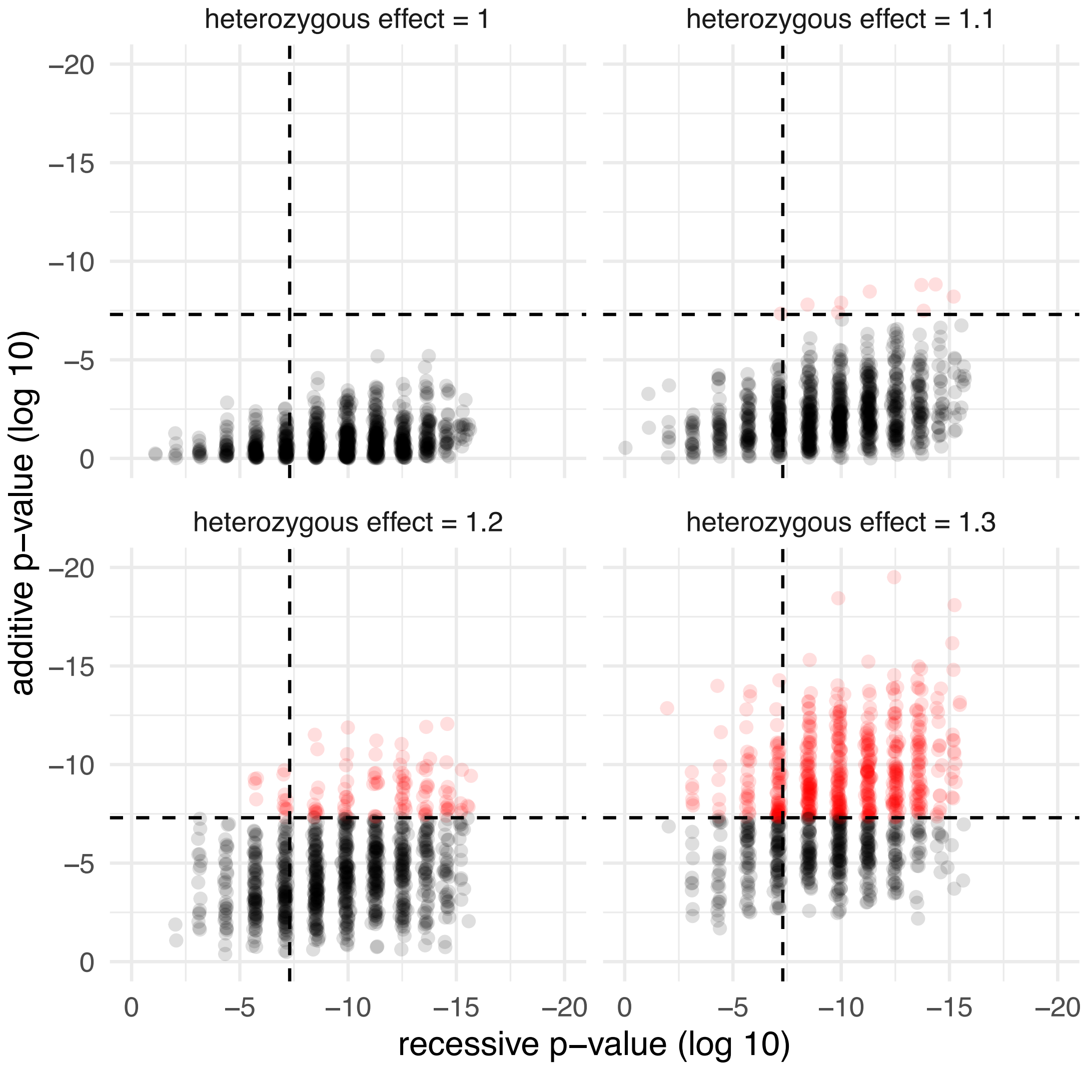

**B**

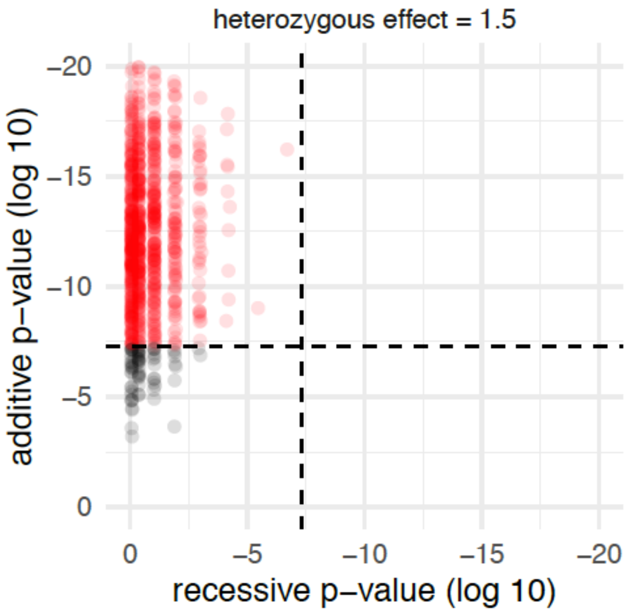
heterozygous effect = 1.5
additive p–value (log 10)
–20
–15
–10
–5
0
recessive p–value (log 10)
0
–5
–10
–15
–20

**Supplementary Figure S5.** Separate pdf. Longitudinal survival curves showing disease onset of homozygous, heterozygous and wildtypes of variants with recessive effects in FinnGen.

**Supplementary Figure S6. *EBAG* is associated with female infertility. A)** Comparing number of offspring in 147,061 women who are wildtype (n=131,141), heterozygous (n=15,455) or homozygous (n=465) for the *EBAG9* variant. Among 7,980 women with children and diagnosed with infertility 71 *EBAG9* homozygotes had **B)** fewer children and **C)** had their first child significantly later (see Supplementary Note 5).

**A**

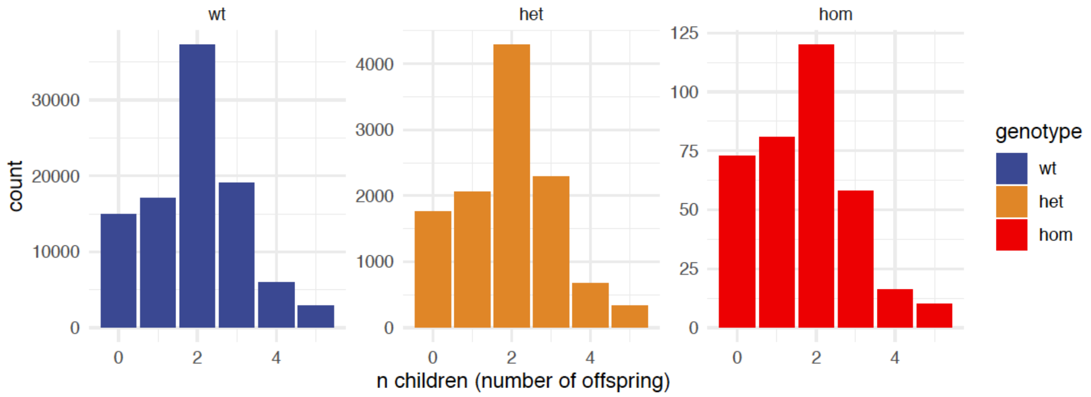


**B**

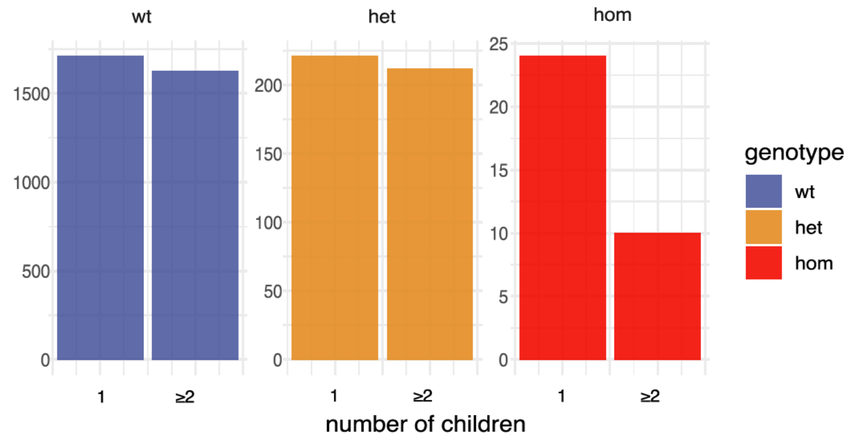


**C**

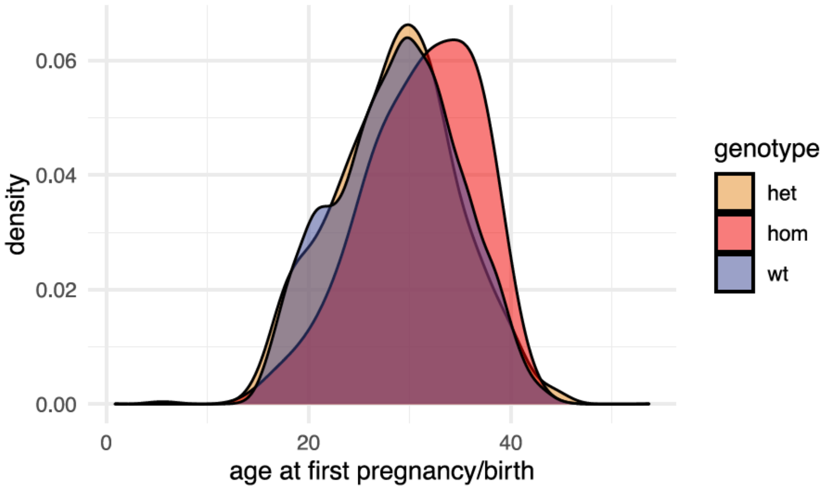

**Supplementary Figure S7. Replications with UKBB data.** We replicated the 31 recessive GWAS hits in FinnGen in the UKBB[5] with a recessive model in SAiGE[6]. Of the 31 variants, 13 had ≥ 5 homozygous carriers in the UKBB of which 8 had a significant recessive p-value in UKBB for the same/similar phenotype. Here, we show recessive betas in FinnGen versus UKBB (betas set to 5, when beta > 5). Plot size corresponds to MAF in gnomAD non-Finnish Europeans and plot color red means the recessive p-value in UKBB is < 0.05.

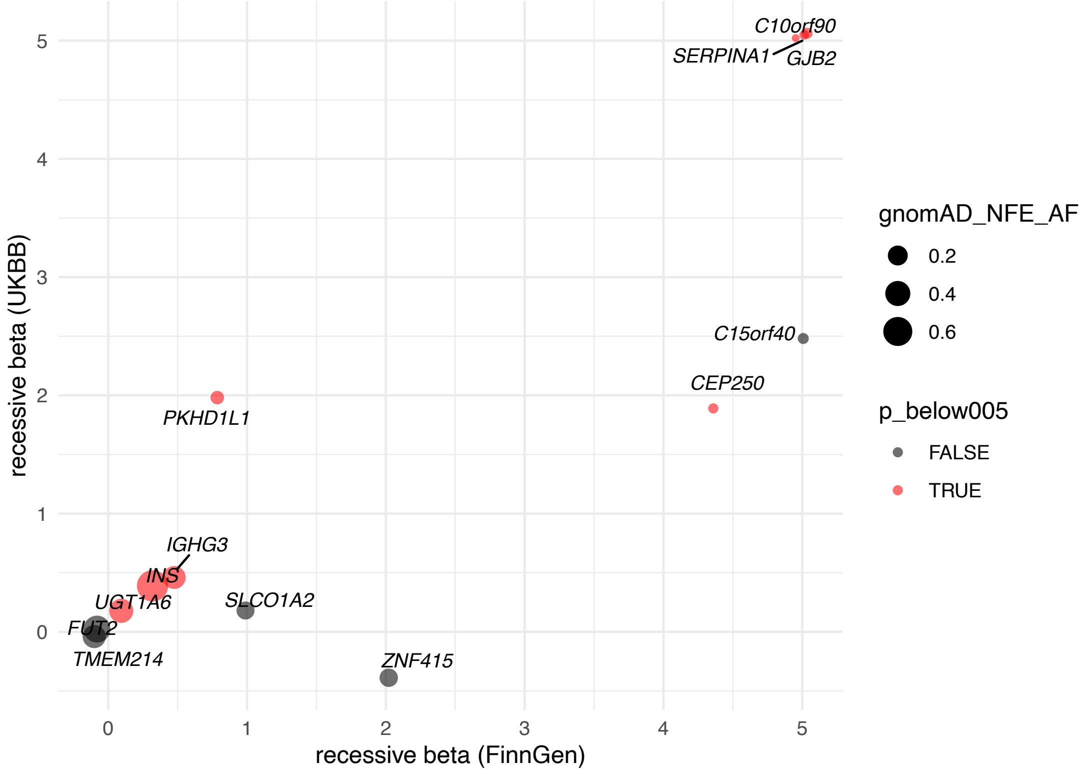

**Supplementary Figure S8. Age at disease onset.**

This forrest plot shows the median age at first diagnosis for each variant with significant homozygous effects in FinnGen. P-values indicate differences in disease onset between respective homozygous or heterozygous compared to wildtype carriers (Wilcoxon rank tests). Bars represent the first and third quartile of age at first diagnosis. Only variants with more than 5 affected homozygous carriers are shown. The y-axis lists gene-disease associations. Homozygotes had significantly earlier (or later for a known homozygous protective variant in *FUT2*[7]) disease onset for 7/31 variants than wildtypes (p-value < 0.0016 with Bonferroni correction for 31 tests, 14/31 tests with nominal p-value < 0.05, Wilcoxon rank test.)

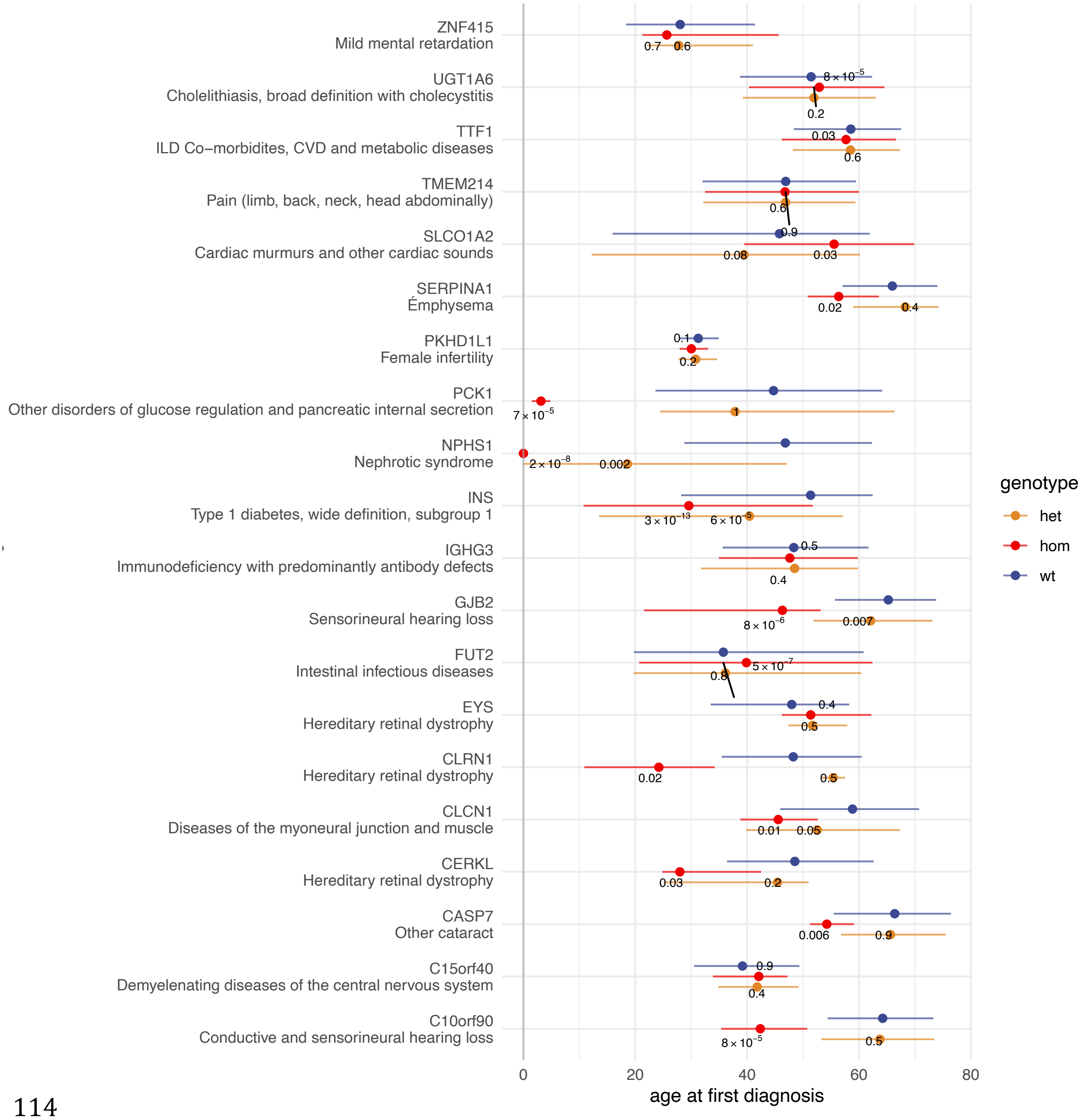

**Supplementary Figure S9. Age at first disease diagnosis of variant carriers in GJB2 (survival plot).** Wt, wildtype; het, heterozygous; hom, homozygous; comp-het, compound heterozygous; GT, genotype. Genotypes of a known pathogenic missense and pLoF variant in *GJB2* associated with hearing loss. Comp-het carry both the pathogenic missense and pLoF variant on different alleles.

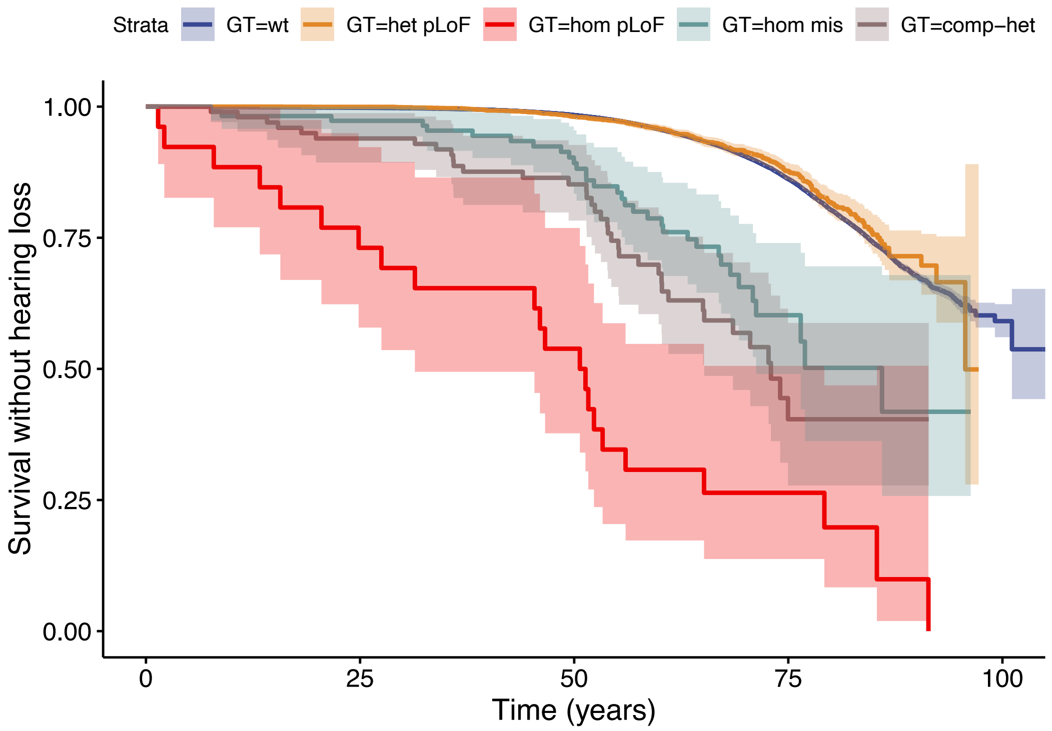

### Supplementary Tables

**Supplementary Table 3. Variants with homozygous associations in FinnGen that overlap known GWAS hits.** Adjacent variants with $r^2$>0.25 associated with the same trait or same parent trait are considered as one locus and only the most significant variant shown per locus. The first five columns show gene, rsid, associated phenotype, recessive p-value and beta in FinnGen. Columns six to eight give the known GWAS phenotypes, SNPs and p-values. LD ($r^2$>=0.1) of the known GWAS SNPs with respective SNPs in FinnGen is given in the last column. FinnGen, FG.

| Gene | rsid | Lead Phenotype FG | p-value FG | beta FG | known GWAS … phenotype in LD | ... SNP | … r2 | … p-value |
|---|---|---|---|---|---|---|---|---|
| *UGT1A6* | rs1105879 | Cholelithiasis | 4.10E-11 | 0.09 | Gallstone disease; Serum bilirubin levels in metabolic syndrome | rs2070959; rs2070959 | 0.8; 0.8 | $9x10^{-11}$; $2x10^{-14}$ |
| *IGHG3* | rs74093865 | Immunodeficiency w. predominantly antibody defects | 4.40E-08 | 0.5 | Composite immunoglobulin trait (IgG/IgM) | rs74093831 | 0.72 | $8x10^{-9}$ |
| *FUT2* | rs601338 | Intestinal infectious diseases | 4.00E-09 | -0.07 | Number of common colds;Vitamin B12 levels | rs601338; rs492602 | 1; 0.99 | $4x10^{-7}$; $5x10^{-17}$ |
| *TMEM214* | rs1124649 | Pain (limb, back, neck, head abdominally) | 3.80E-11 | -0.06 | CRP level; polymyalgia rheumatica | rs11681145 | 0.78;1 | $9x10^{-12}$; 0.004 |
| *INS* | rs3842753 | Type 1 diabetes | 1.20E-41 | 0.24 | Type 1 diabetes | rs689 | 0.99 | $5x10^{-196}$ |
| *SLCO1A2* | rs10841795 | Cardiac murmurs and other cardiac sounds | 1.40E-08 | 1 | Blood metabolite levels | rs12829704 | 0.6 | $9x10^{-21}$ |

**Supplementary Table 4. Variants with homozygous associations in FinnGen that could be tested for replication in the UK biobank[5].** The last three rows colored in brown/grey show homozygous associations of different coding variants in the UK biobank than the coding variant with a genomewide significant hit in FinnGen. UK biobank, UKB; FinnGen, FG; MAF, minor allele frequency.

| Gene | rsid | Phenotype FinnGen | Phenotype UKBB | MAF FG | MAF NFE gnomAD | p-value FG | beta FG | p-value UKB | beta UKB |
|---|---|---|---|---|---|---|---|---|---|
| *APPL2* | rs138491961 | noninflammatory disorders of uterus, except cervix | N85 noninflammatory disorders of uterus, except cervix | 0.0032 | 0.0015 | 3.69E-08 | 5.54 | NA | NA |
| *CEP250* | rs56259282 | Varicose veins | I83 Varicose veins of lower extremities | 0.0046 | 0.012 | 1.27E-08 | 4.33 | 0.027 | 1.97 |
| *C10orf90* | rs139123090 | Conductive and sensorineural hearing loss | H90 Conductive and sensorineural hearing loss | 0.0085 | 0.0045 | 2.25E-12 | 5.96 | 0.00012 | 56.7 |
| *PCK1* | rs201186470 | Glucose regulation and pancreatic secretion | E16 Other disorders of pancreatic internal secretion | 0.011 | 0.00045 | 4.65E-08 | 9.77 | NA | NA |
| *GJB2* | rs398123814 | Sensorineural hearing loss | H90 Conductive and sensorineural hearing loss | 0.012 | 0.0092 | 2.86E-28 | 6.49 | 3.11E-08 | 82.0 |
| *C15orf40* | rs115868366 | Demyelenating diseases of the central nervous system | Multiple sclerosis | 0.013 | 0.018 | 3.94E-08 | 11.3 | 0.058 | 2.53 |
| *CASP7* | rs141266925 | Other cataract | H26 Other cataract | 0.015 | 0.0035 | 2.47E-16 | 4.77 | NA | NA |
| *EBAG9* | rs1422 | Female infertility | N97 Female infertility | 0.056 | 0.041 | 1.60E-11 | 2.0 | 0.0018 | 3.90 |
| *SERPINA1* | rs28929474 | Emphysema | J43 Emphysema | 0.02 | 0.018 | 2.16E-21 | 28.0 | 1.43E-26 | 20.6 |
| *INS* | rs3842753 | Type 1 diabetes | Type 1 diabetes | 0.21 | 0.72 | 5.71E-46 | 0.28 | 6.31E-23 | 0.35 |
| *IGHG3* | rs74093865 | Immunodeficiency w. predominantly antibody defects | D80 Immunodeficiency w. predominantly antibody defects | 0.4 | 0.3 | 4.44E-08 | 0.50 | 0.027 | 0.53 |
| *UGT1A6* | rs1105879 | Cholelithiasis, broad definition with cholecystitis | Cholelithiasis and cholecystitis | 0.45 | 0.35 | 1.53E-11 | 0.08 | 1.58E-10 | 0.16 |
| *SLCO1A2* | rs10841795 | Cardiac murmurs and other cardiac sounds | R01 Cardiac murmurs and other cardiac sounds | 0.11 | 0.14 | 1.38E-08 | 1.03 | 0.47 | 0.14 |
| *TMEM214* | rs1124649 | Pain (limb, back, neck, head abdominally) | General pain for 3+ months | 0.28 | 0.31 | 3.79E-11 | -0.06 | 0.41 | -0.10 |
| *ZNF415* | rs10410030 | Mild mental retardation | F81 Specific developmental disorders of scholastic skills | 0.097 | 0.15 | 1.56E-08 | 2.10 | 0.42 | -0.37 |
| *FUT2* | rs601338 | Intestinal infectious diseases | Intestinal infection | 0.37 | 0.48 | 4.01E-09 | -0.07 | 0.21 | -0.03 |
| *CASP7* | rs2227309 | Other cataract | H26 Other cataract | 0.28 | 0.26 | NA | NA | 0.016 | 0.07 |
| *APPL2* | rs2272495 | noninflammatory disorders of uterus, except cervix | N85 noninflammatory disorders of uterus, except cervix | 0.23 | 0.20 | NA | NA | 0.0085 | 0.18 |
| *PCK1* | rs1804160 | Glucose regulation and pancreatic secretion | E16 Other disorders of pancreatic internal secretion | 0.043 | 0.023 | NA | NA | 0.038 | 1.67 |

**Supplementary Table 5. Variants with homozygous associations in FinnGen that could be tested for additive associations in the UK biobank (UKBB)[5].** The columns show gene, rsid, phenotype name in UKBB, additive p-value and beta in UKBB and the phenotype name in FinnGen.

| Gene | rsid | Phenotype UKBB | p-value | beta | Phenotype FinnGen |
|---|---|---|---|---|---|
| *ACOXL* | rs80265967 | Gestational diabetes | >0.05 | NA | Diabetes mellitus in pregnancy |
| *APPL2* | rs138491961 | noninflammatory disorders of uterus, except cervix | >0.05 | NA | N85 Other noninflammatory disorders of uterus, except cervix |
| *CASP7* | rs141266925 | Cataract | 4.50E-12 | 0.7 | Cataract |
| *C10orf90* | rs139123090 | Conductive and sensorineural hearing loss | >0.05 | NA | Conductive and sensorineural hearing loss |
| *C15orf40* | rs115868366 | Demyelenating diseases of the CNS | 1.40E-02 | 0.34 | multiple sclerosis |
| *CEP250* | rs56259282 | Varicose veins | >0.05 | NA | Varicose veins |
| *CLCN1* | rs55960271 | Diseases of the myoneural junction and muscle | >0.05 | NA | Myasthenia gravis and other myoneural disorders |
| *CNGB1* | rs201162411 | Hereditary retinal dystrophy | >0.05 | NA | Hereditary retinal dystrophy |
| *DMBT1* | rs189221852 | Nephropathy | 2.20E-04 | 5.3 | Non-proliferative glomerulonephritis |
| *EBAG9* | rs1422 | Female infertility | >0.05 | NA | Female infertility |
| *IGHG3* | rs74093865 | Immunodeficiency w. predominantly antibody defects | >0.05 | NA | Immunodeficiency with predominantly antibody defects |
| *INS* | rs3842753 | Type 1 diabetes | 9.10E-20 | 0.28 | Type 1 diabetes |
| *PCK1* | rs1804160 | Other disorders of pancreatic internal secretion | >0.05 | NA | Glucose regulation and pancreatic secretion |
| *PLTP* | rs56126980 | Unspecified lump in breast | >0.05 | NA | Lump or mass in breast |
| *SERPINA1* | rs28929474 | Emphysema | 2.30E-19 | 1.3 | Emphysema |
| *SLCO1A2* | rs10841795 | Cardiac murmurs and other cardiac sounds | >0.05 | NA | Cardiac murmurs and other cardiac sounds |
| *TMEM214* | rs1124649 | Pain (limb, back, neck, head abdominally) | 4.00E-03 | -0.13 | polymyalgia rheumatica |
| *UGT1A6* | rs200711686 | Cholelithiasis | 1.40E-06 | 0.064 | Cholelithiasis |
| *USP5* | rs200723728 | Endometriosis of ovary | >0.05 | NA | Endometriosis |
| *USP8* | rs147742292 | Postpartum haemorrhage | >0.05 | NA | Postpartum haemorrhage |
| *ZNF415* | rs10410030 | Mild mental retardation | 2.90E-02 | 0.31 | Mild cognitive impairment |

#### Supplementary Notes.

**1) Known disease causing variants in FinnGen.** We studied 15,140 variants that were annotated in ClinVar[4] in 176,899 Finns from the FinnGen research project. These included 311 coding variants that were annotated as (likely) pathogenic, 147 of them by multiple submitters or an expert panel (corresponds to ClinVar review status of 2-4 out of 4 possible stars) which we thus describe as being annotated as (likely) pathogenic with "high confidence". 298 further variants had "conflicting interpretations of pathogenicity" but were classified as (likely) pathogenic by at least one submitter. 3.4% of study participants carried at least one of ten high confidence (likely) pathogenic variant in one of nine genes causing disease with dominant inheritance according to OMIM. For 6 out of the 10 (likely) pathogenic variants we found expected genomewide significant disease associations (see also Supplementary Table S1) in the genes *FLG* (atopic dermatitis, 2690 and 1050 heterozygotes, respectively), *MYOC* (glaucoma, 1023 heterozygotes), *PALB2* (breast cancer, 512 heterozygotes), *JAK2* (Polycythaemia vera, 194 heterozygotes) and *MYBPC3* (Hypertrophic cardiomyopathy, 116 heterozygotes). We did not find genomewide significant associations for 4 of 10 high confidence (likely) pathogenic variants of which *CHEK2* was genomewide significant for breast cancer in the later FinnGen datafreeze R5. Variants in *COL5A2, G6PD* and *PKD1* were not significant in later releases. For *G6PD* this may be expected as the variant is rare (58 hemizygous males) and functionally predicted to lead to Glucose-6-phosphate dehydrogenase activity of 15 to 25 percent of wildtype (Cappellini 1994, Cappellini 1995, De Vita 1989, Frigerio 1994, Lenzerini 1969, Rattazzi 1969), thus characteristic of a moderate (Class III) deficiency where hemolysis is only triggered by stressors. For variants in *COL5A2* and *PKD1* we would expect at least nominally significant disease associations in later FinnGen releases, given they contain > 60,000 individuals with joint disorders and >1,000 individuals with cystic kidney disease. The fact that there are only two submitters for the variant in *COL5A2* and one submitter classifying the variant in *PKD1* as VUS in a later ClinVar release adds to the suspicion that they may in fact not be truly disease associated.

All of 0.41% of participants (n=726) carried one of 98 high confidence (likely) pathogenic variants in homozygous state in one of 86 disease genes with known recessive inheritance, 41.8% were heterozygous carriers. Most frequent homozygous mutants were in the genes *BTD* (partial biotinidase deficiency, 523 homozygotes), *GJB2* (nonsyndromic sensorineural hearing loss, 113 homozygotes) and *SERPINA1* (Alpha-1-antitrypsin deficiency, 77 homozygotes). We found expected recessive phenotype associations of *GJB2* and *SERPINA1* variants with genomewide significance. We were however surprised by the lack of genomewide signifiant disease associations for the known pathogenic variant [*BTD*:p.(Asp444His)] linked to biotinidase deficiency. *BTD*:p.(Asp444His) has been reported as pathogenic for biotinidase deficiency by 17 submitters in ClinVar and experimentally led to a ~25% reduction in biotinidase activity[8]. Homozygotes for *BTD*:p.(Asp444His) are thus expected to have ca. 50% of normal serum biotinidase activitiy. As partial biotinidase deficiency is however characterized by 10%-30% of normal serum biotinidase activity[9] the lack of disease associations above chance expectation for *BTD*:p.(Asp444His) homozygotes makes sense. We can thus empirically confirm the experimental hypothesis that the variant should only cause disease when compound heterozygous with a variant leading to near-zero biotinidase activity.

We can thus show that we are well-powered to identify known associations across a wide range of disease phenotypes in our dataset. In addition, it can serve as a resource to verify or falsify previously described disease associations. For verification purposes the statistical finemapping can be additionally helpful to identify associations' causal variants.

**2) (Likely) benign variants in FinnGen.** We studied effects of variants listed in ClinVar [4] as "(likely) benign". Those are regarded as "not the cause for a patient's disorder" ACMG [10]. However, we found more phenotype associations than expected in (likely) benign ClinVar variants as a group.

To test whether these might potentially just correlate with actual causal variants in linkage disequilibrium we assessed how often they were included in the 95%-cs for a given GWAS locus [11]. We found on the contrary, that 142 (likely) benign missense variants were slightly more often in the 95%-cs than 307 missense variants not annotated in ClinVar (Fisher's Exact test, p-value 0.05, OR 1.5, 95%-CI 1.0-2.4). 16 represented even the most likely causal variant for a GWAS locus. The ClinVar annotation of these variants as benign was of good quality as 7 of the 16 benign top causal variants had a one star and 9 of 16 a two star review status in ClinVar.

Expectedly, the disease phenotypes in FinnGen – though often related - are mostly different from the known OMIM phenotypes of the respective genes. Nevertheless, disease associations of benign variants may still be counterintuitive as they are often interpreted as generally disease-irrelevant [12,13]. How could we then explain global disease associations for them? Firstly, it is plausible that the frequent diagnostic sequencing and analysis of disease-associated genes with high clinical interest also results in submission of many benign variants to ClinVar for those genes. Hypothesizing that genes with many disease-causing variants would therefore also get many ClinVar submissions of benign variants, we found indeed a positive correlation of the number of (likely) pathogenic and (likely) benign variants per gene in ClinVar ($r^2$ 0.41, rho 0.07, respective p $<10^{-16}$). Genes with many disease-causing variants - particularly relevant to human disease - may be more likely involved in additional diseases than disease-irrelevant genes [14]. In line with that we found indeed that (likely) benign ClinVar missense variants had more phenotype associations than missense variants in different genes but that signal disappeared mostly when missense variants were matched to the same genes. In addition, (likely) benign variants are of significantly higher MAF (0.07±0.12, mean, sd) than other ClinVar variants (0.01±0.05) in FinnGen which is expected as MAF is a criterion for labeling a variant as benign ACMG[10]. So we are well-powered to find significant disease associations for benign variants, particularly in a bottleneck population like Finland that enriches disease variants to higher frequencies [3]. Of note, when investigating unique ClinVar variants in the 95%-cs, 19 out of 67 (likely) benign ClinVar variants had protective effects on disease, while only 1 out of 27 ClinVar risk variants (i.e. (likely) pathogenic, conflicting or risk factor variants) was protective (Fisher's Exact test, p-value 0.01 OR 10, 95%-CI 1.4-443). While their identified effects are more often protective than for known risk variants the majority of associations increase disease risk also in benign variants!

In conclusion, we think it is important to keep in mind that variants labeled as benign are not generally neutral and can still contribute to disease.

**3) Simulations of recessive and additive effects.** We performed simulations of genotype and phenotype data to assess different effect sizes of heterozygotes on the significance of the recessive versus additive model. We simulated genotype and phenotype data aiming to resemble our own data. Specifically, we generated genotype counts of wildtype, heterozygotes and mutant homozygotes in 200,000 individuals of a variant with an allele frequency of 0.01 following Hardy Weinberg Equilibrium (using R library HardyWeinberg[15]). We then generated random phenotype data of cases versus controls of a disease with a prevalence of 0.05 using the rbinom function in base R. We set the probability of homozygotes to develop the disease to 10x compared to wildtype. We generated different sets of cases versus controls with heterozygote effect sizes of 1 (=no effect), 1.1, 1.2 and 1.3. We then performed logistic regression using the glm function in R using an additive and a recessive model, respectively.

When setting to no heterozygous effect an additive model found the homozygous association with genomewide significance in only 1/1000 simulations while the recessive model found it in 912/1000 simulations. For increasing sizes of heterozygous effects the additive model's significance increased accordingly (see Supplementary Figure S4A). We found similar results for varying effect sizes and MAF. For well-powered associations (concerning variants with high MAF) the additive model also picked up associations only driven by homozygotes (data not shown). This is in agreement with observations in our real data e.g. the variant in *INS* (Table 2). When however simulating an expected 'additive hit' (Supplementary Figure S4B, heterozygous effect 1.5, homozygous effect 2) the recessive model is genomewide significant in 0/1000 simulations while the additive model is genomewide significant in 928/1000 times.

**4) Compound heterozygous effects in *GJB2* and *CERKL*.** The genes *GJB2* and *EYS* harbored more than one pathogenic variant with significant homozygous effects in our data so we sought to explore compound heterozygous (=comp-het) effects. LD between the two *GJB2* variants ($r^2$=0.002) was negligible but the two *EYS* variants were in strong LD ($r^2$=0.47) so we could not investigate comp-het effects for *EYS*. In *GJB2,* we found early onset sensorineural hearing loss in 100 *GJB2* comp-hets, i.e. individuals with one pathogenic missense and one pLoF *GJB2* variant (p-value $6x10^{-28}$, HR 6.1, coxph) that each also had effects in homozygous state. In agreement with the variants' different homozygous effect sizes, the effect of comp-het was in between the homozygous pLoF (HR 15, p-value $7x10^{-37}$) and homozygous missense variant (HR 3.9, p-value $8x10^{-15}$) (see Figure 3C). We also tested the effects of another (likely) pathogenic missense variant in *GJB2.* LD with the other *GJB2* variants was negligible ($r^2$~0.00 with both). That variant was at a much lower MAF of 0.004. Therefore, our dataset included only 3 individuals in homozygous state, 4 in compound heterozygous state with the pLoF *GJB2* variant and 5 in compound heterozygous state with the other missense *GJB2* variant. Combined, all 12 variants were associated with early onset sensorineural hearing loss with similar effect as the other pathogenic variants in *GJB2* (p-value $3x10^{-12}$, HR 11.8, coxph). As none of the *GJB2* variants had effects in heterozygous state, we can assume true comp-het effects of the three (likely) pathogenic variants. We also investigated comp-het effects in *CERKL* as there was an additional known pathogenic pLoF variant in our data chr2:181656814:C:A (hg38) at low frequency (MAF 0.0019) which was thus not well-powered to reach significance in homozygous state in the pheWAS. When excluding individuals with homozygous and comp-het *CERKL* variants we did not find a heterozygous effect of any of the pathogenic variants on the phenotype of retinal dystrophy anymore suggesting comp-het effects drove the initial nominally significant heterozygous effect.

**5) Novel recessive disease associations.** Here, we highlight two variants with large homozygous effects in genes with no previous disease associations, in *CASP7* associated with cataract, *C10orf90* associated with hearing loss and *EBAG9* associated with female infertility.

We would first like to present a missense variant in *CASP7* with a MAF of 0.015 in Finland (4x enriched in Finns) that we found associated with adult-onset cataract. The variant had homozygous Mendelian-like effects on the distinct phenotypes "other cataract" (p-value = $2x10^{-16}$, beta 4.8, most frequent ICD10 codes in 43 homozygous carriers: 13x secondary cataract and 7x presenile cataract) and "senile cataract" (p-value = $2x10^{-12}$, beta 2.1). While there were too few homozygous carriers in the UKBB we could replicate a genomewide significant effect on phenotype "all cataracts" in the additive model in UKBB (p-value = $5x10^{-12}$, beta 0.7) suggesting a potential heterozygous effect or homozygous effect of other variants in LD. There was also some evidence for an effect on "other cataracts" in heterozygous carriers in FinnGen (SAiGE GWAS p-value 0.003, beta 0.25). Age at first diagnosis was nominally significantly earlier in homozygous carriers (see Supplementary Figure 5). In the age group below 63 (mean retirement age in Finland) 50% of homozygous carriers (13/26) but only 3.4% (94/2750) of heterozygous and 3.1% (2710/87799) of wildtype carriers had already reported cataracts in our data. *CASP7* encodes Caspase-7, an executing enzyme in apoptosis (programmed cell death) [16]. In lens development, cell organelles are degraded to create transparent cells by a process very similar to apoptosis. Further, deactivating apoptosis genes in mice (e.g. *Casp3*[17] or *Apaf-1* and *p53*[18]) has been shown to lead to cataract and there has been evidence for apoptosis involved in human non-congenital cataract[19,20]. *CASP7* is therefore a biologically plausible and to the best of our knowledge first Mendelian gene for adult-onset cataract, the leading cause of vision loss worldwide with no other effective treatment than surgery and therefore particularly affecting low/middle income countries (Source: WHO, 2012).

The missense variant in *C10orf90* was associated with sensorineural hearing loss also with Mendelian effects (recessive p-value = $5x10^{-12}$, beta 6.0; additive p-value > $10^{-4}$), which we could replicate in the UKBB (recessive p-value $1x10^{-4}$, beta 56). The variant was 2.8x enriched in Finns with a MAF of 0.0083 in Finland. Homozygous carriers had significantly earlier onset of diseases: among 14k individuals diagnosed with hearing loss carriers of homozygous variants in *C10orf90* were first diagnosed at a mean age of 40±17 (mean±sd) compared to wildtype carriers (63±16, p-value $8*10^{-5}$, Wilcoxon-rank test). There is less known about the gene *C10orf90* which is also called fragile-site associated tumor suppressor (*FATS*)[21]. However, mutations in its most closely related paralog gene *ALMS1* cause the ciliopathy "Alstrom syndrome" with sensorineural hearing loss as one of its main symptoms[22]. As there is also some evidence for a role of *C10orf90* in cilia formation[23], it is plausible that *C10orf90* would also cause hearing via an effect on cilia. However, since there are no other symptoms in variant carriers beyond expectation apart from hearing loss, this is likely restricted to hearing loss and therefore distinct from *ALMS1*.

Next, we find a noncoding variant in *EBAG9* with a homozygous effect on female infertility. This is the only noncoding variant we investigated in our study but we believe it is the more likely causal candidate than the linked PTV variant in *PKHD1L1* we initially identified as it has a larger effect (beta 2.0 compared to 0.21), it is more likely causal after finemapping (PIP 0.2 while the PTV in *PKHD1L1* is not in the credible set) and *EBAG9* is a known estrogen responsive gene. 48 of 4,505 women (1.1%) with infertility in FinnGen are homozygous for the *EBAG9* variant. In FinnGen R6 data, we found that 465 female *EBAG9* homozygotes have fewer children than 131,141 wildtypes (p-value 0.0002, Wilcoxon rank test). Also among all 7,980 women diagnosed with infertility who also had children 71 *EBAG9* homozygotes were more severely affected as they had fewer children (p-value 0.005, Wilcoxon rank test) and had their first child significantly later (p-value 0.005, Wilcoxon rank test).
