## Supplementary Figure S5A-AE for "Mono- and bi-allelic effects of coding variants on disease in 176,899 Finns"

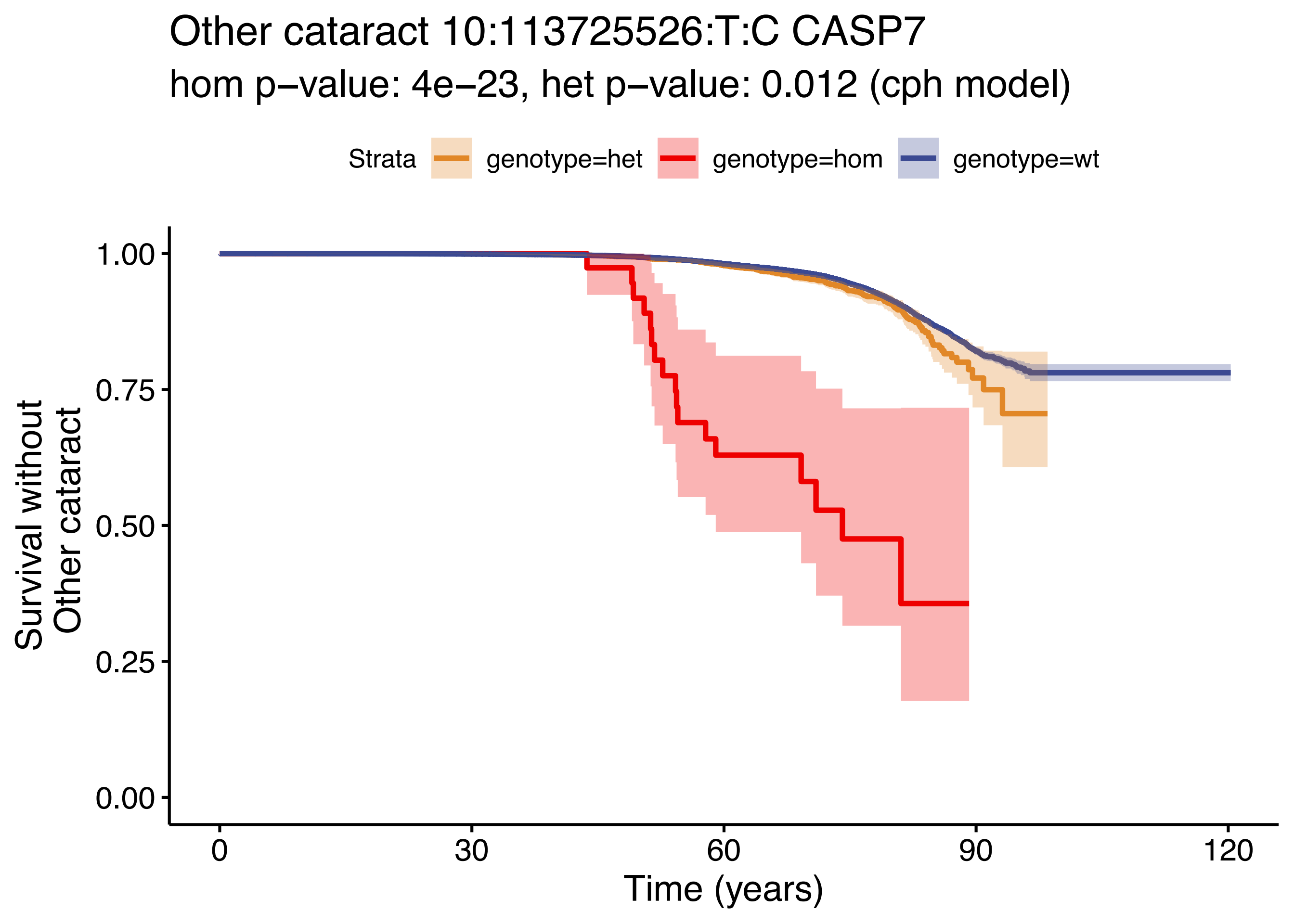

Other cataract 10:113725526:T:C CASP7
hom p−value: 4e−23, het p−value: 0.012 (cph model)
Strata
genotype=het
genotype=hom
genotype=wt
Survival without Other cataract
1.00
0.75
0.50
0.25
0.00
0
30
60
90
120
Time (years)

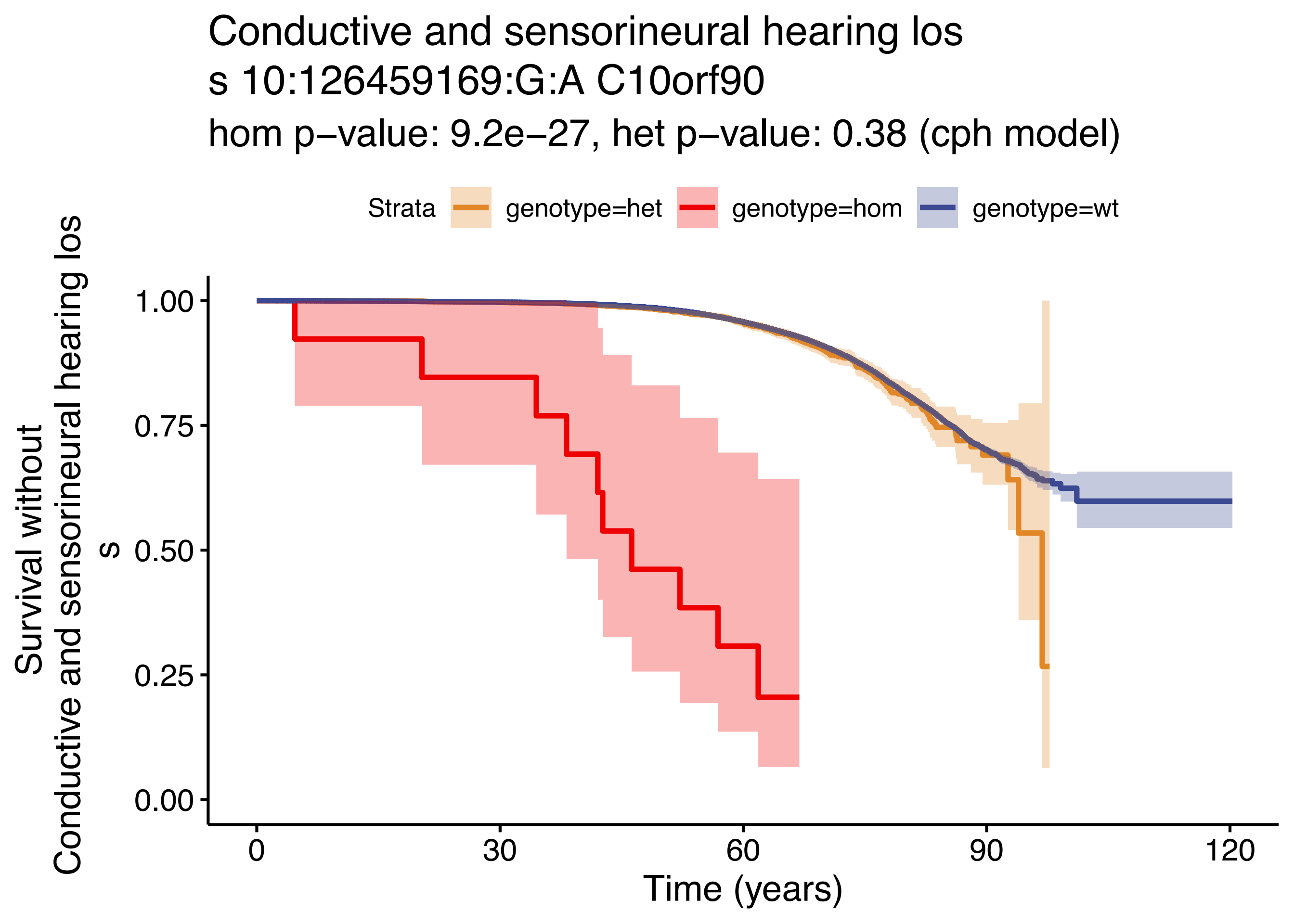

Conductive and sensorineural hearing los
s 10:126459169:G:A C10orf90
hom p−value: 9.2e−27, het p−value: 0.38 (cph model)
Strata
genotype=het
genotype=hom
genotype=wt
Survival without
Conductive and sensorineural hearing los
s
1.00
0.75
0.50
0.25
0.00
0
30
60
90
120
Time (years)

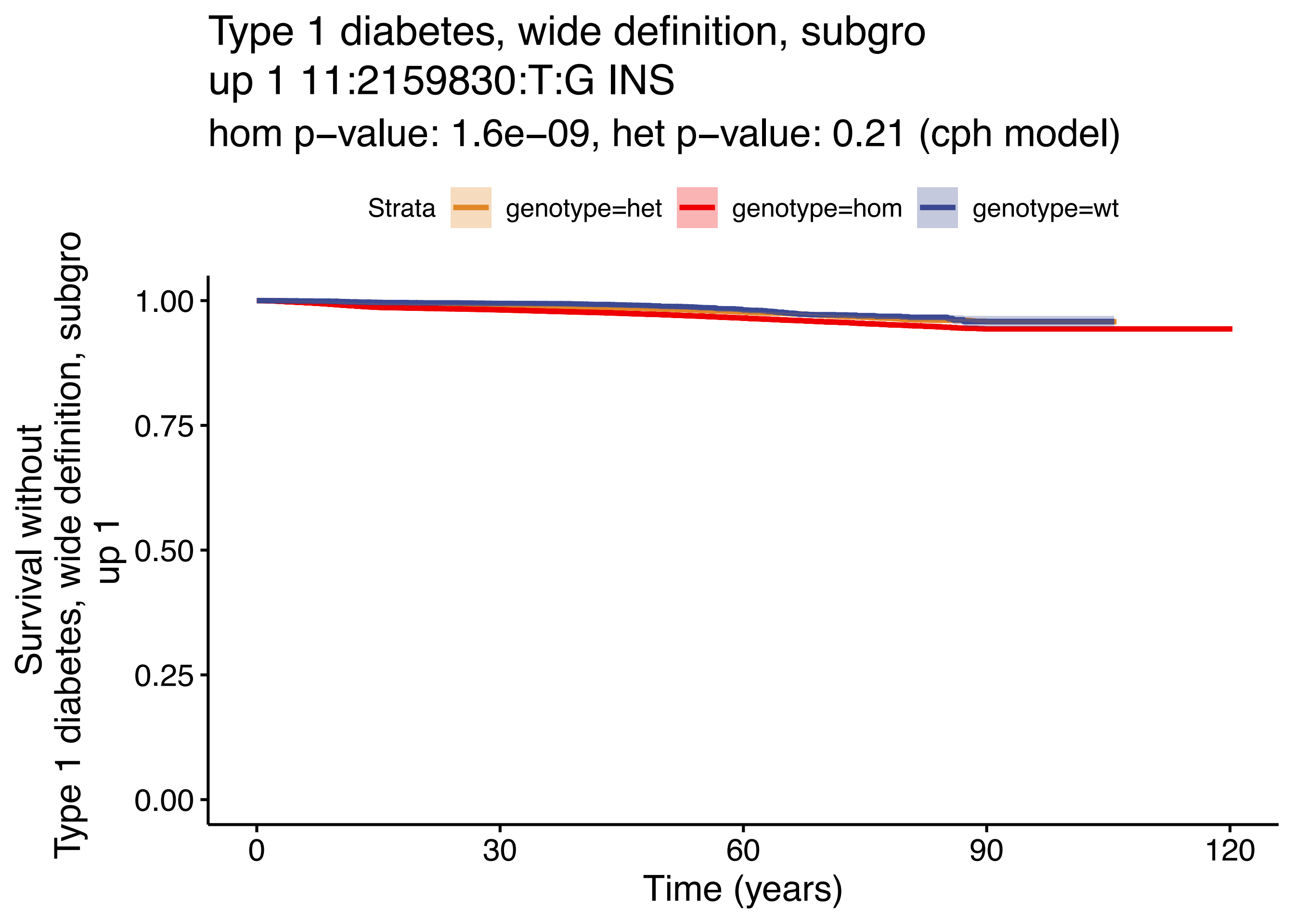
Type 1 diabetes, wide definition, subgro
up 1 11:2159830:T:G INS
hom p−value: 1.6e−09, het p−value: 0.21 (cph model)
Strata
genotype=het
genotype=hom
genotype=wt
Survival without
Type 1 diabetes, wide definition, subgro
up 1
1.00
0.75
0.50
0.25
0.00
0
30
60
90
120
Time (years)

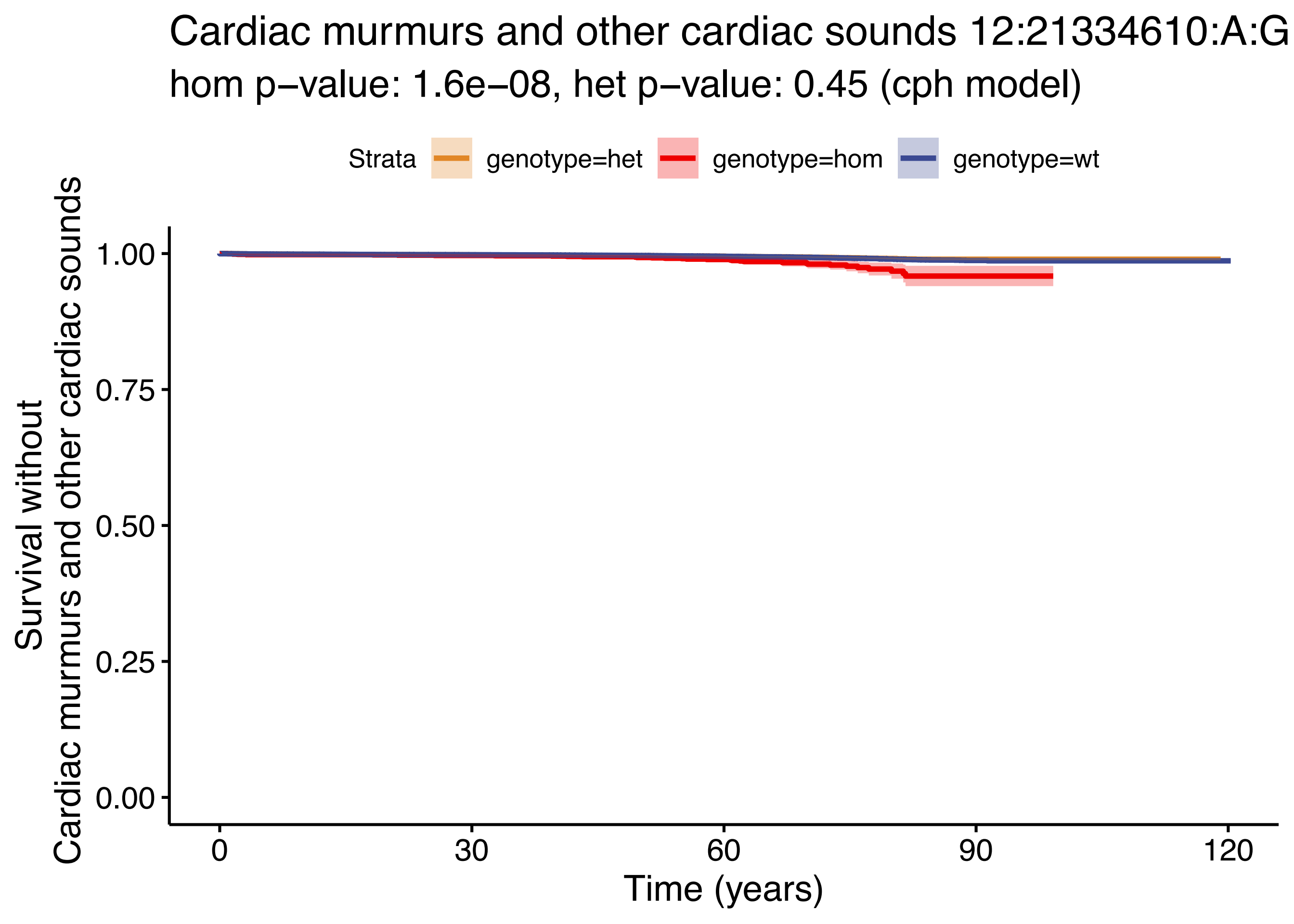

Cardiac murmurs and other cardiac sounds 12:21334610:A:G
hom p–value: 1.6e–08, het p–value: 0.45 (cph model)
Strata
genotype=het
genotype=hom
genotype=wt
Survival without
Cardiac murmurs and other cardiac sounds
1.00
0.75
0.50
0.25
0.00
0
30
60
90
120
Time (years)

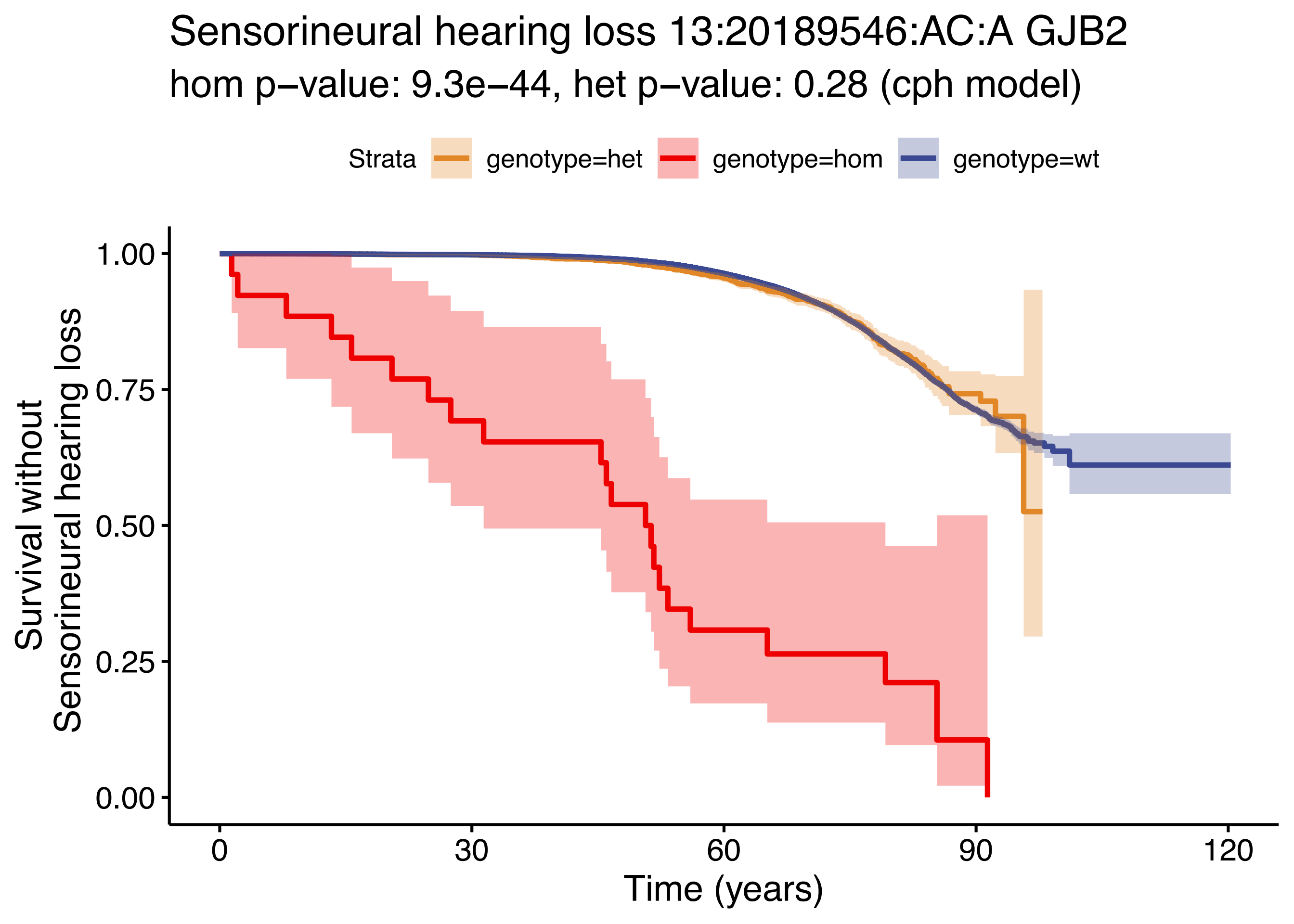

Sensorineural hearing loss 13:20189546:AC:A GJB2
hom p−value: 9.3e−44, het p−value: 0.28 (cph model)
Strata
genotype=het
genotype=hom
genotype=wt
Survival without
Sensorineural hearing loss
1.00
0.75
0.50
0.25
0.00
0
30
60
90
120
Time (years)

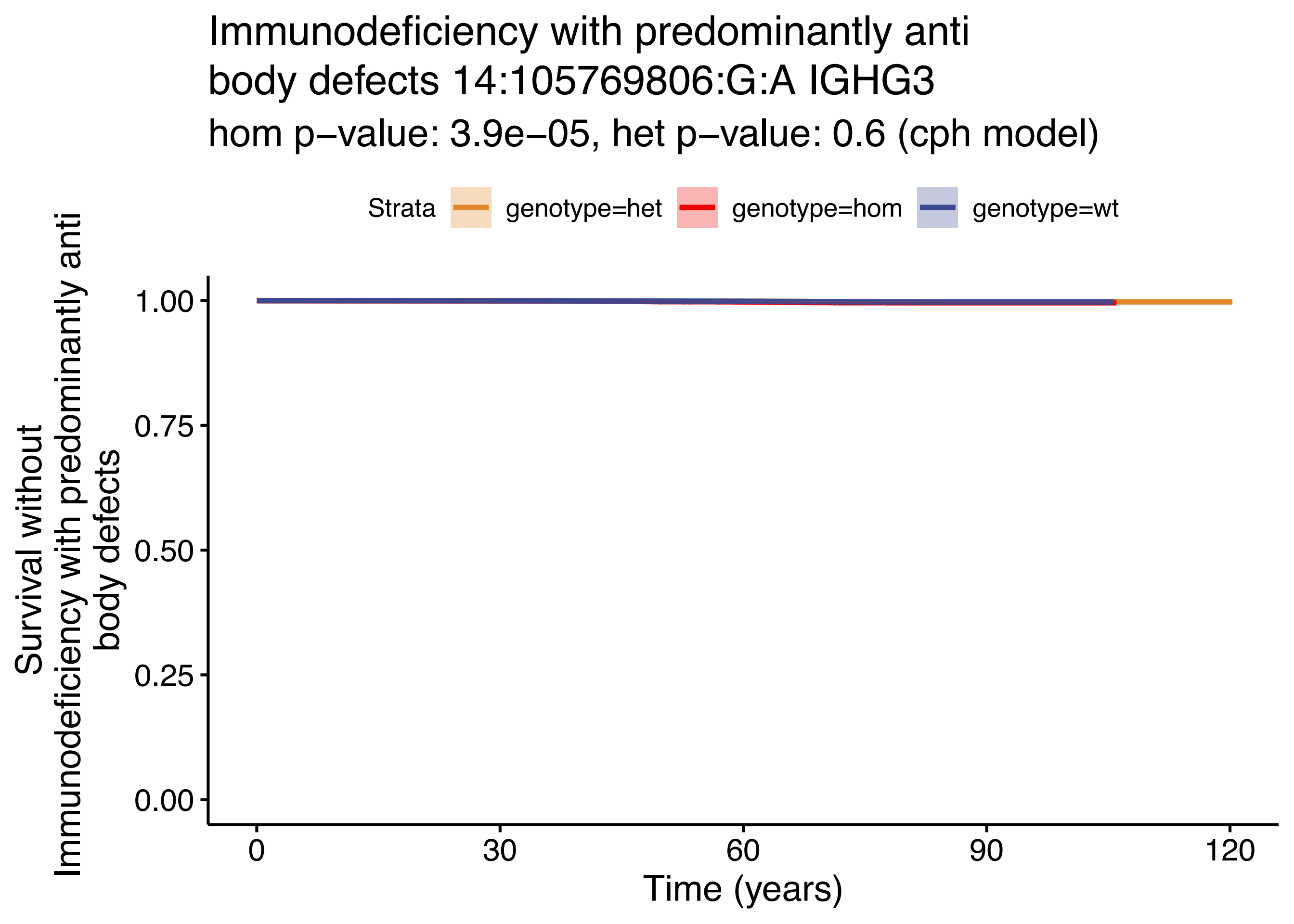
Immunodeficiency with predominantly anti
body defects 14:105769806:G:A IGHG3
hom p−value: 3.9e−05, het p−value: 0.6 (cph model)
Strata
genotype=het
genotype=hom
genotype=wt
Survival without
Immunodeficiency with predominantly anti
body defects
1.00
0.75
0.50
0.25
0.00
0
30
60
90
120
Time (years)

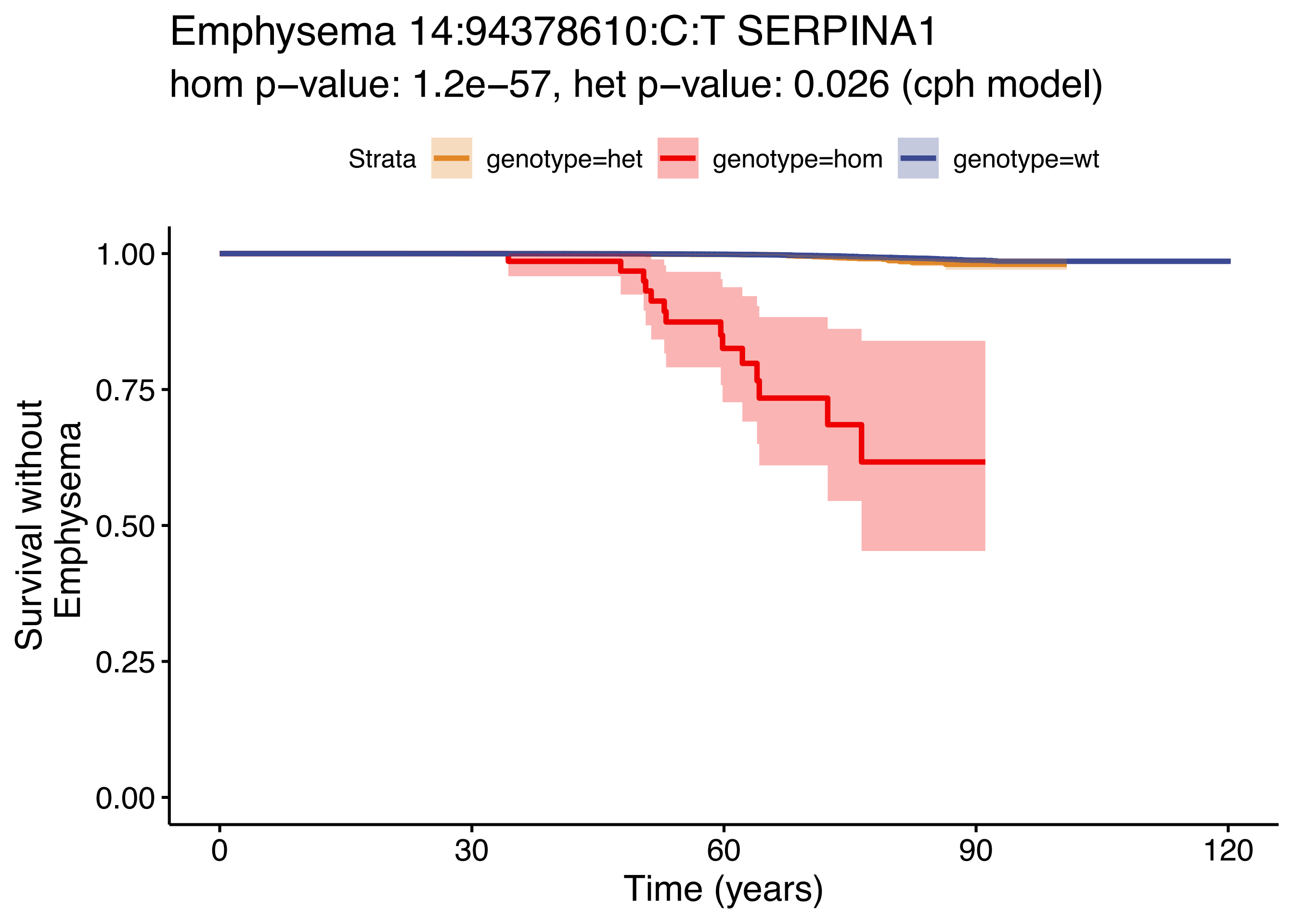

Emphysema 14:94378610:C:T SERPINA1
hom p−value: 1.2e−57, het p−value: 0.026 (cph model)
Strata
genotype=het
genotype=hom
genotype=wt
Survival without Emphysema
1.00
0.75
0.50
0.25
0.00
0
30
60
90
120
Time (years)

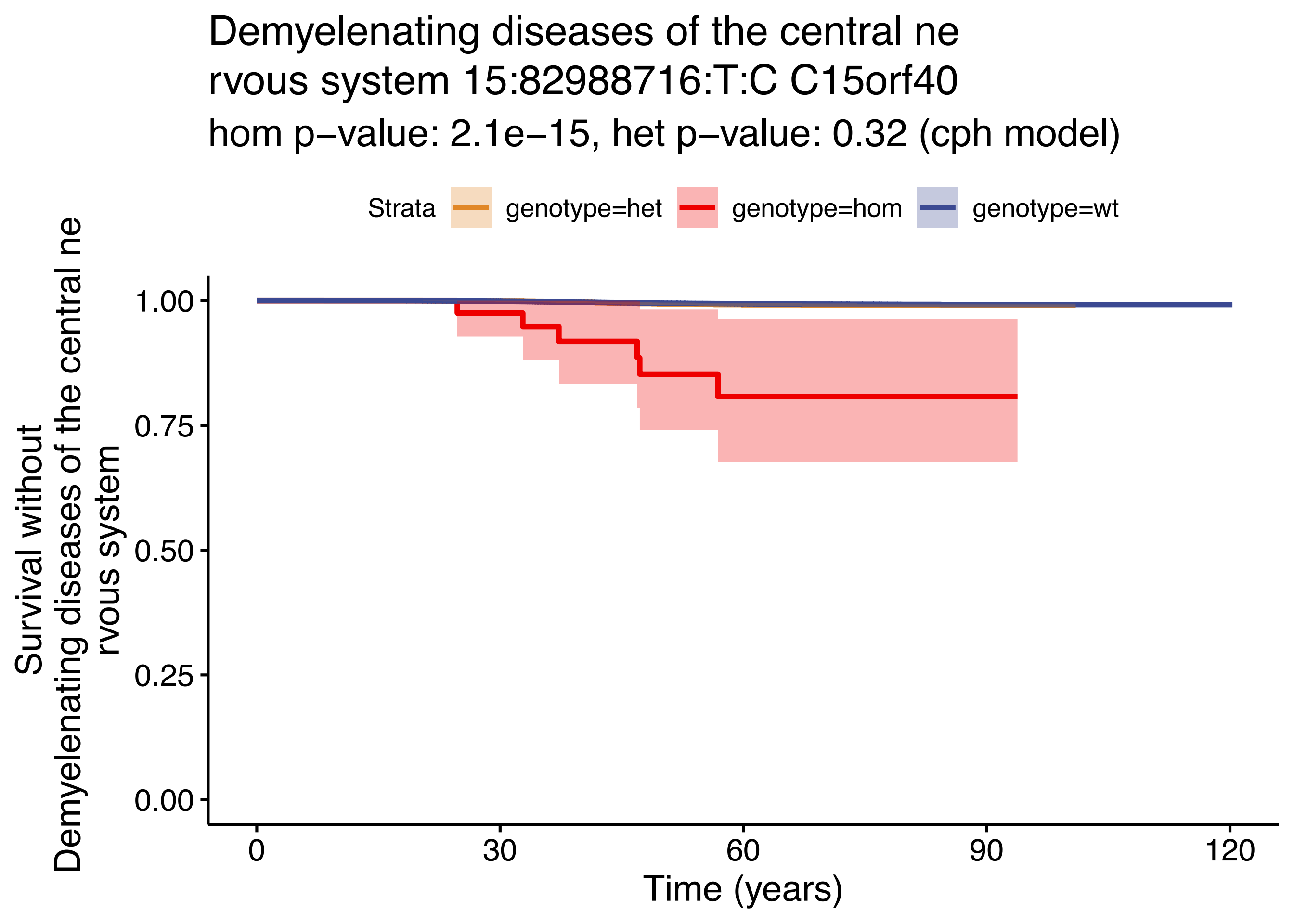

Demyelenating diseases of the central ne
rvous system 15:82988716:T:C C15orf40
hom p−value: 2.1e−15, het p−value: 0.32 (cph model)
Strata
genotype=het
genotype=hom
genotype=wt
Survival without
Demyelenating diseases of the central ne
rvous system
1.00
0.75
0.50
0.25
0.00
0
30
60
90
120
Time (years)

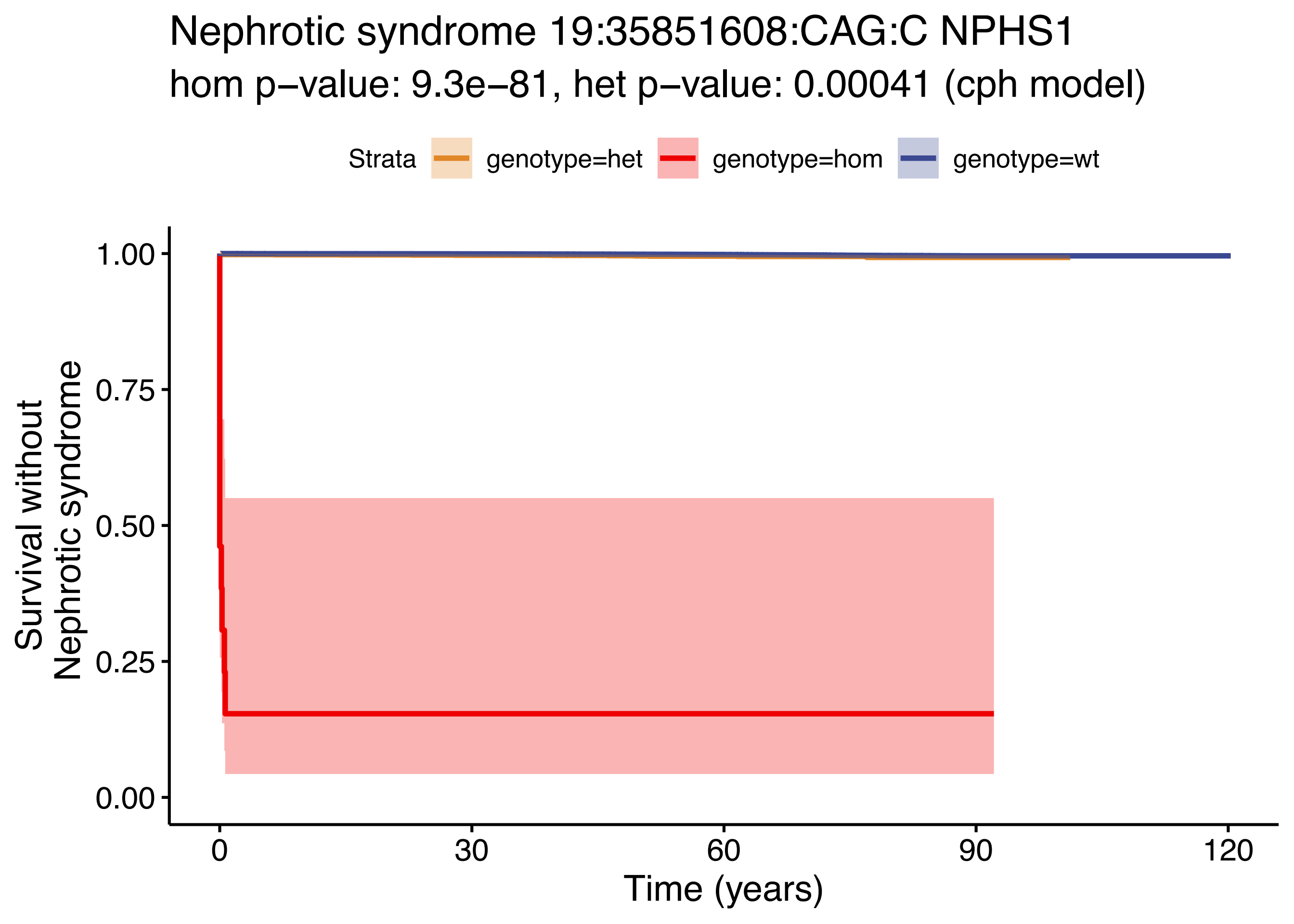

Nephrotic syndrome 19:35851608:CAG:C NPHS1
hom p−value: 9.3e−81, het p−value: 0.00041 (cph model)
Strata
genotype=het
genotype=hom
genotype=wt
Survival without
Nephrotic syndrome
1.00
0.75
0.50
0.25
0.00
0
30
60
90
120
Time (years)

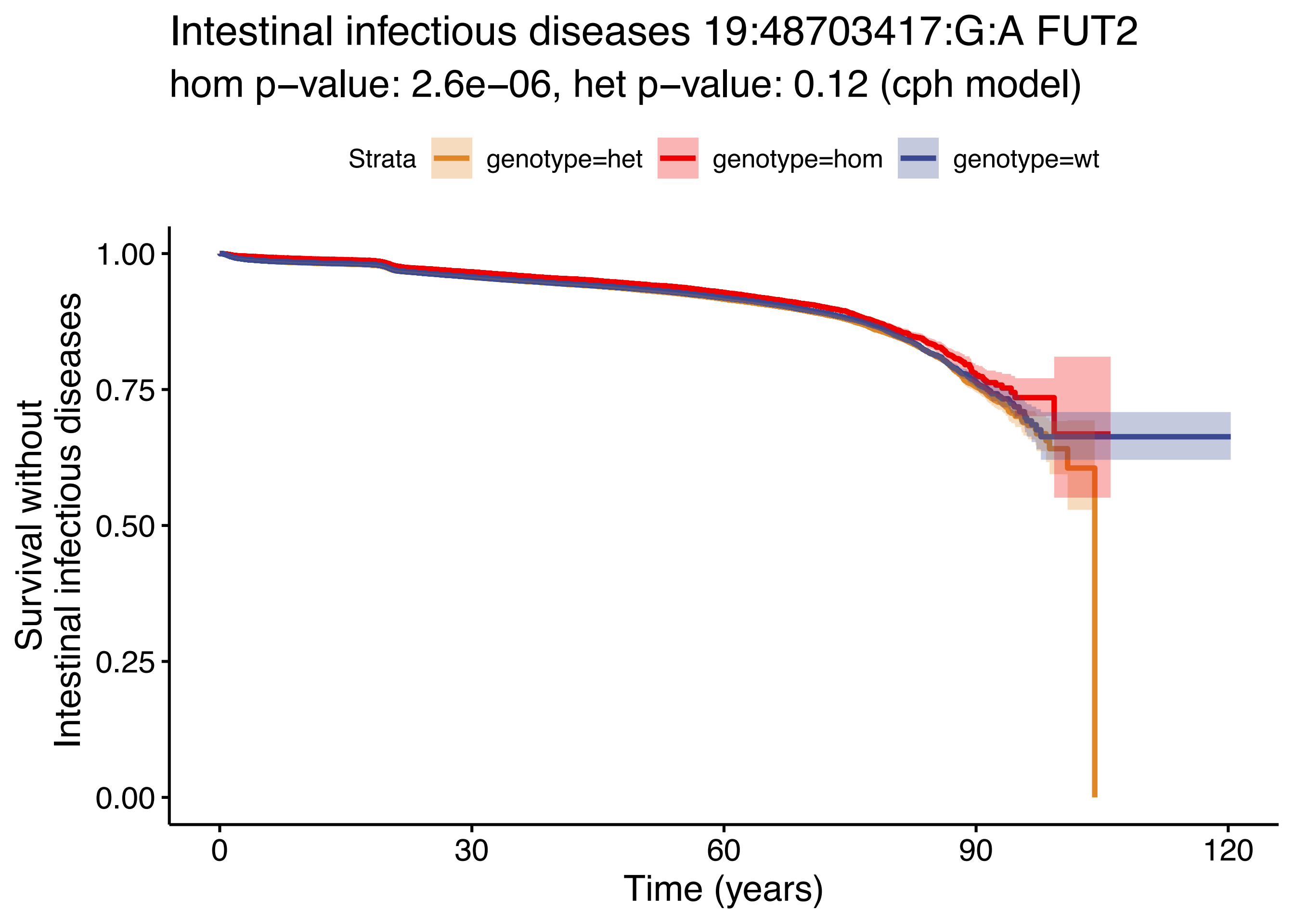
Intestinal infectious diseases 19:48703417:G:A FUT2
hom p−value: 2.6e−06, het p−value: 0.12 (cph model)
Strata
genotype=het
genotype=hom
genotype=wt
Survival without
Intestinal infectious diseases
1.00
0.75
0.50
0.25
0.00
0
30
60
90
120
Time (years)

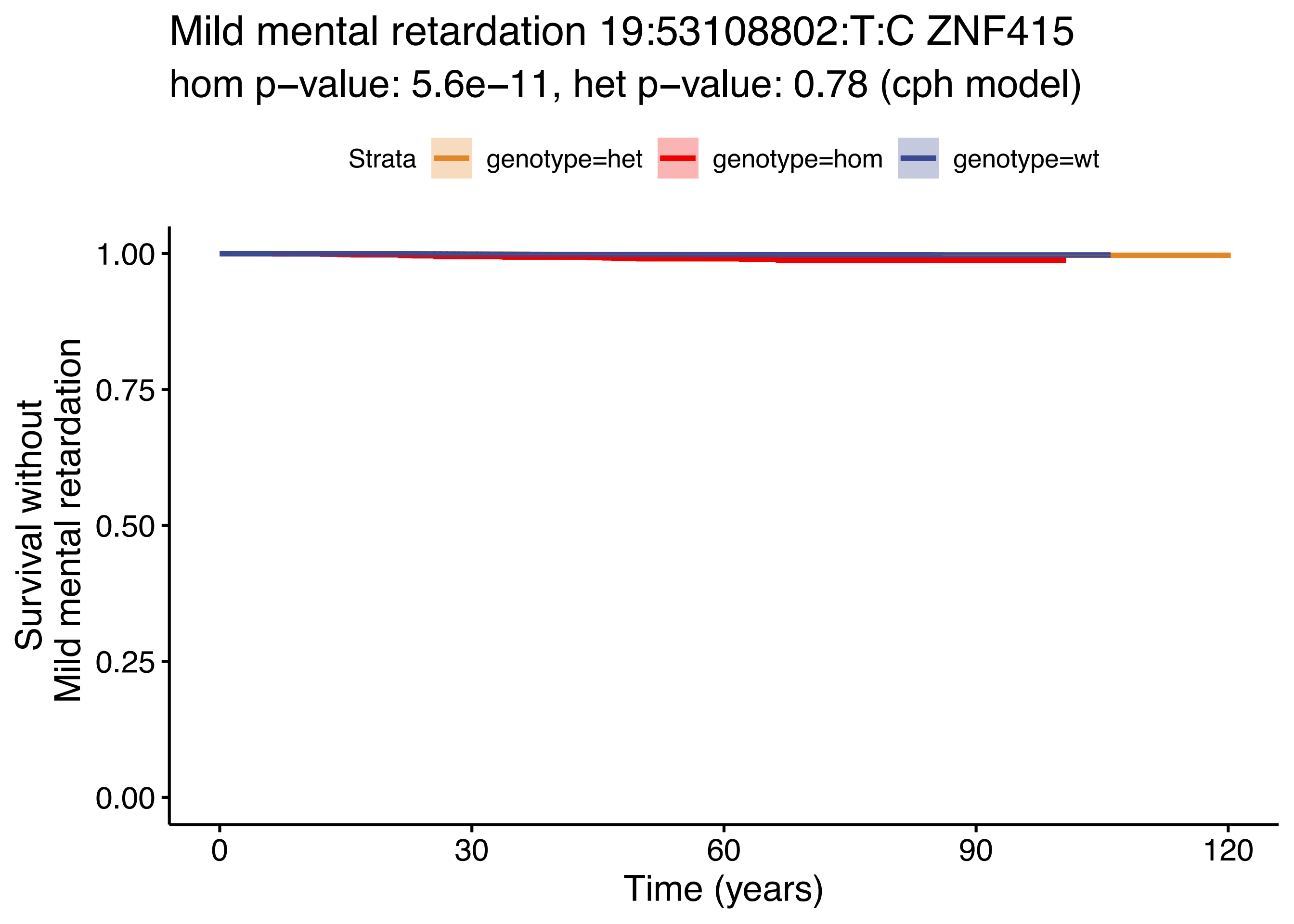
Mild mental retardation 19:53108802:T:C ZNF415
hom p−value: 5.6e−11, het p−value: 0.78 (cph model)
Strata
genotype=het
genotype=hom
genotype=wt
Survival without
Mild mental retardation
1.00
0.75
0.50
0.25
0.00
0
30
60
90
120
Time (years)

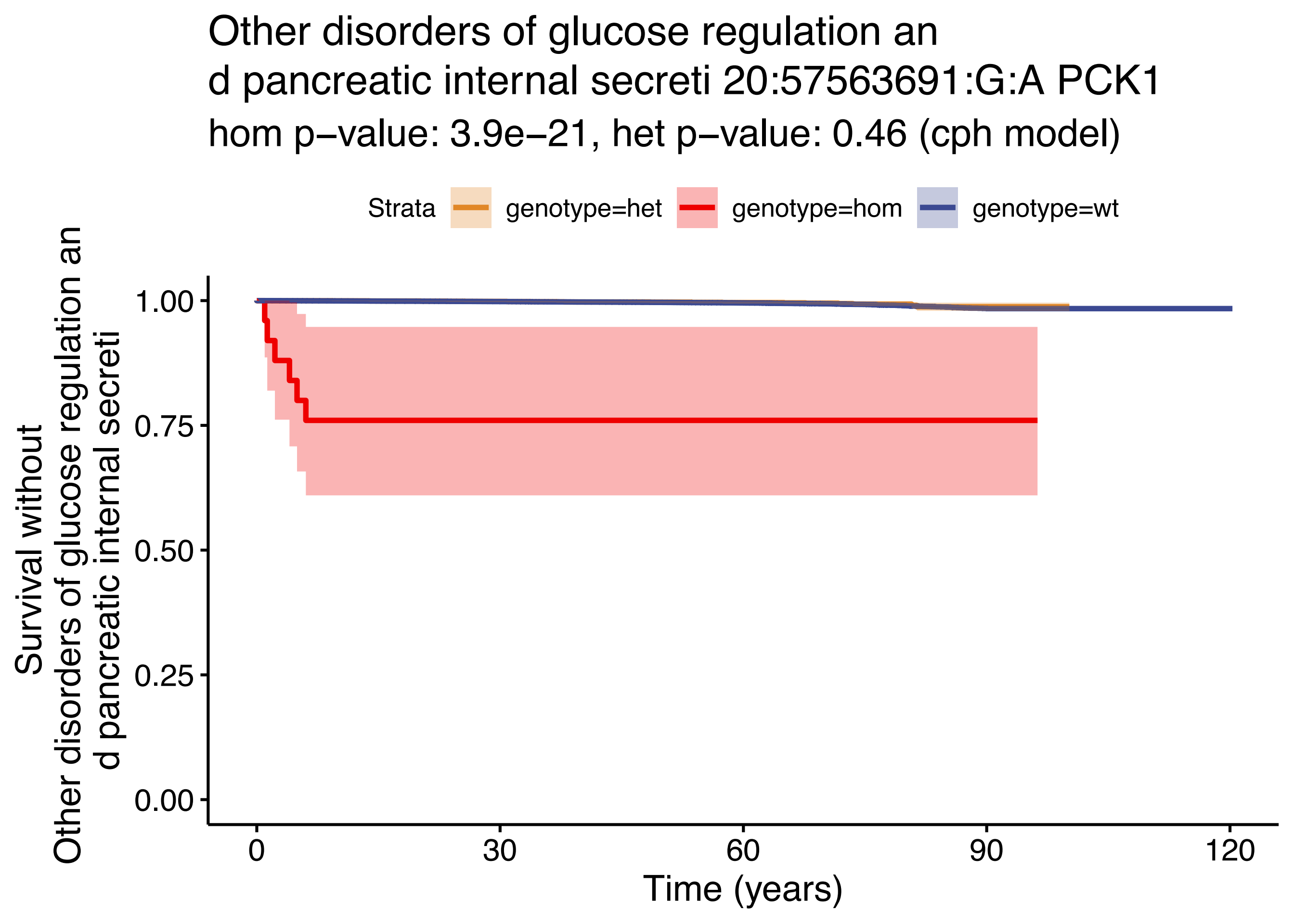

Other disorders of glucose regulation an
d pancreatic internal secreti 20:57563691:G:A PCK1
hom p−value: 3.9e−21, het p−value: 0.46 (cph model)
Strata
genotype=het
genotype=hom
genotype=wt
Survival without
Other disorders of glucose regulation an
d pancreatic internal secreti
1.00
0.75
0.50
0.25
0.00
0
30
60
90
120
Time (years)

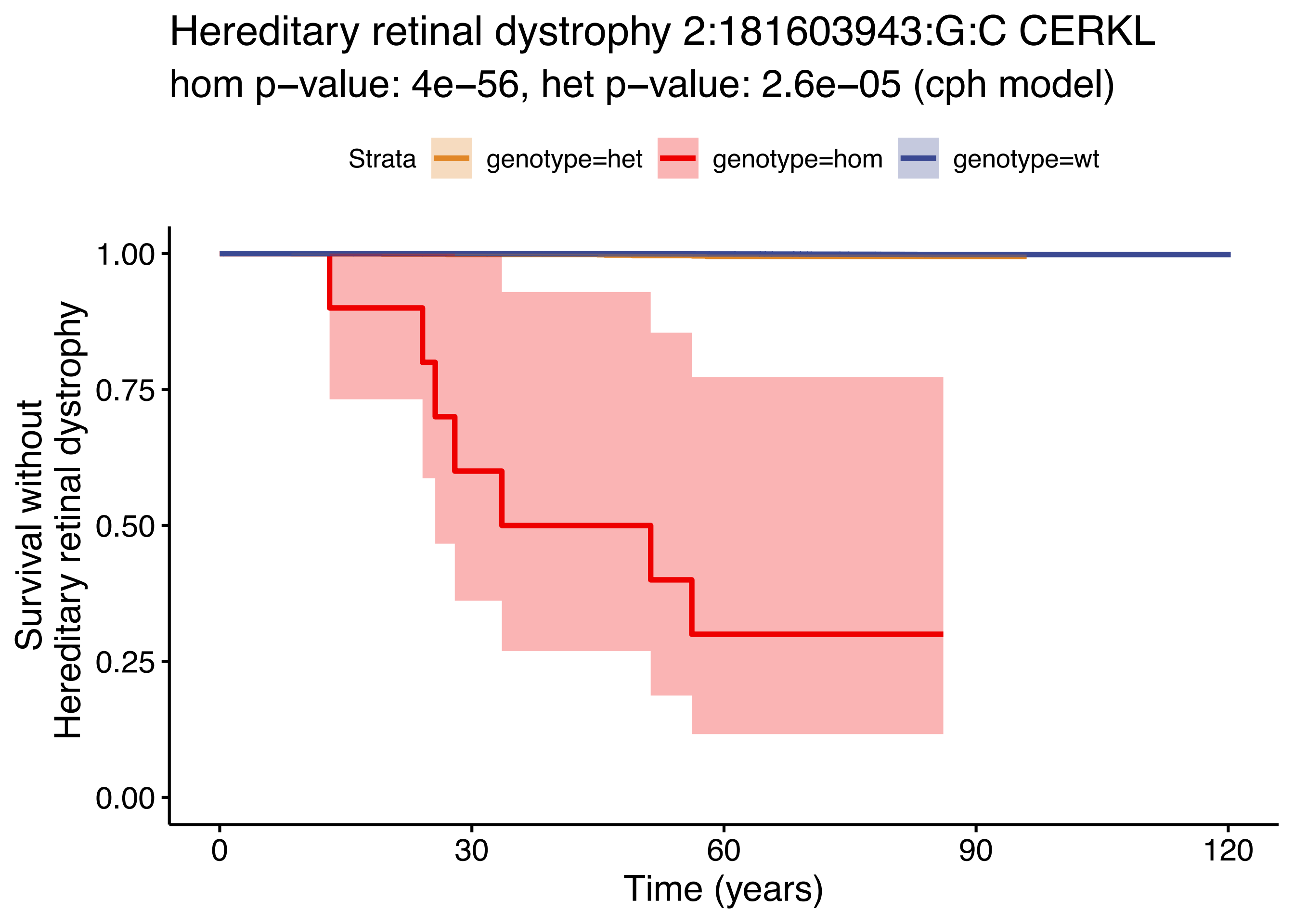

Hereditary retinal dystrophy 2:181603943:G:C CERKL
hom p−value: 4e−56, het p−value: 2.6e−05 (cph model)
Strata
genotype=het
genotype=hom
genotype=wt
Survival without
Hereditary retinal dystrophy
1.00
0.75
0.50
0.25
0.00
0
30
60
90
120
Time (years)

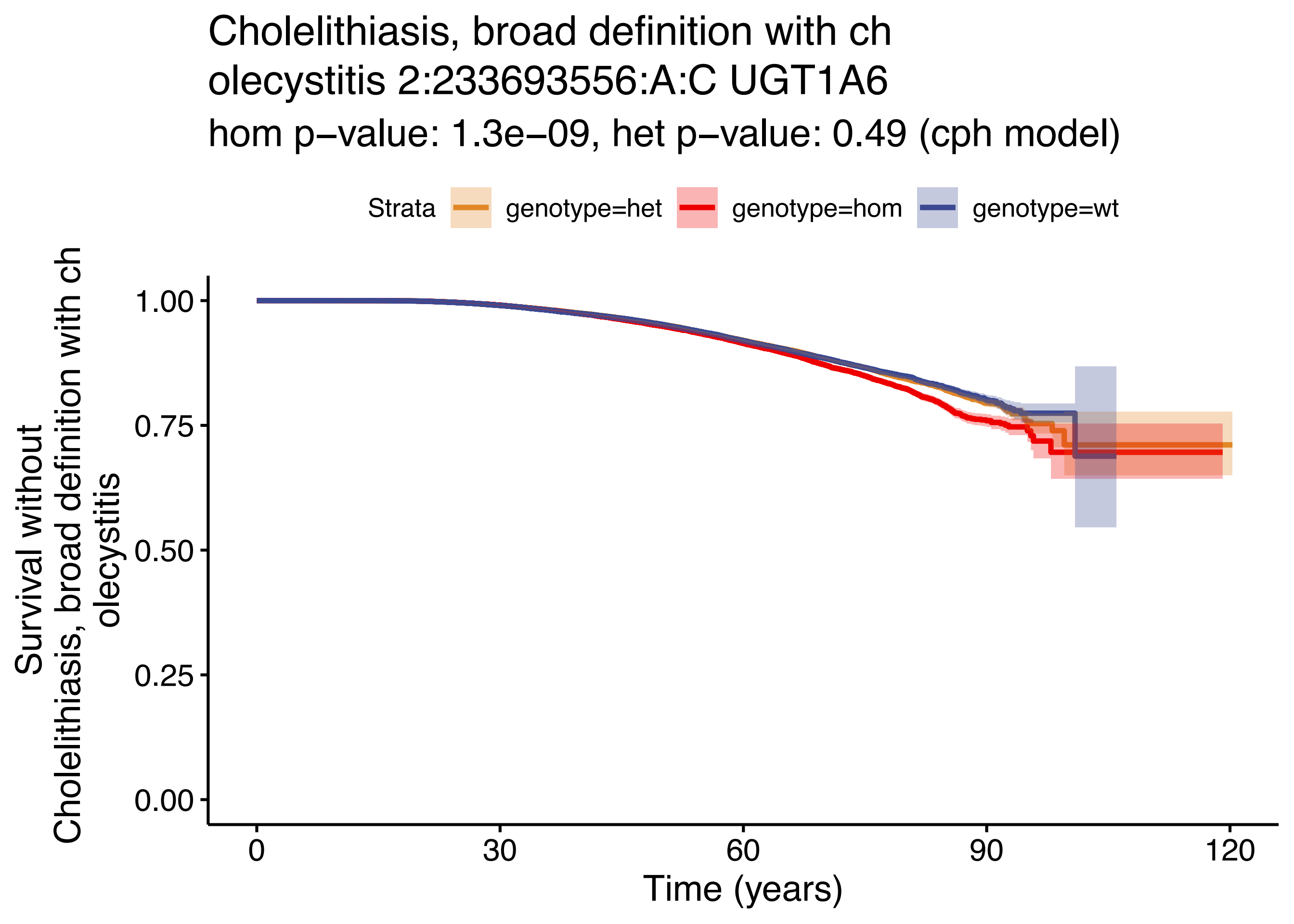

Cholelithiasis, broad definition with ch
olecystitis 2:233693556:A:C UGT1A6
hom p−value: 1.3e−09, het p−value: 0.49 (cph model)
Strata
genotype=het
genotype=hom
genotype=wt
Survival without
Cholelithiasis, broad definition with ch
olecystitis
1.00
0.75
0.50
0.25
0.00
0
30
60
90
120
Time (years)

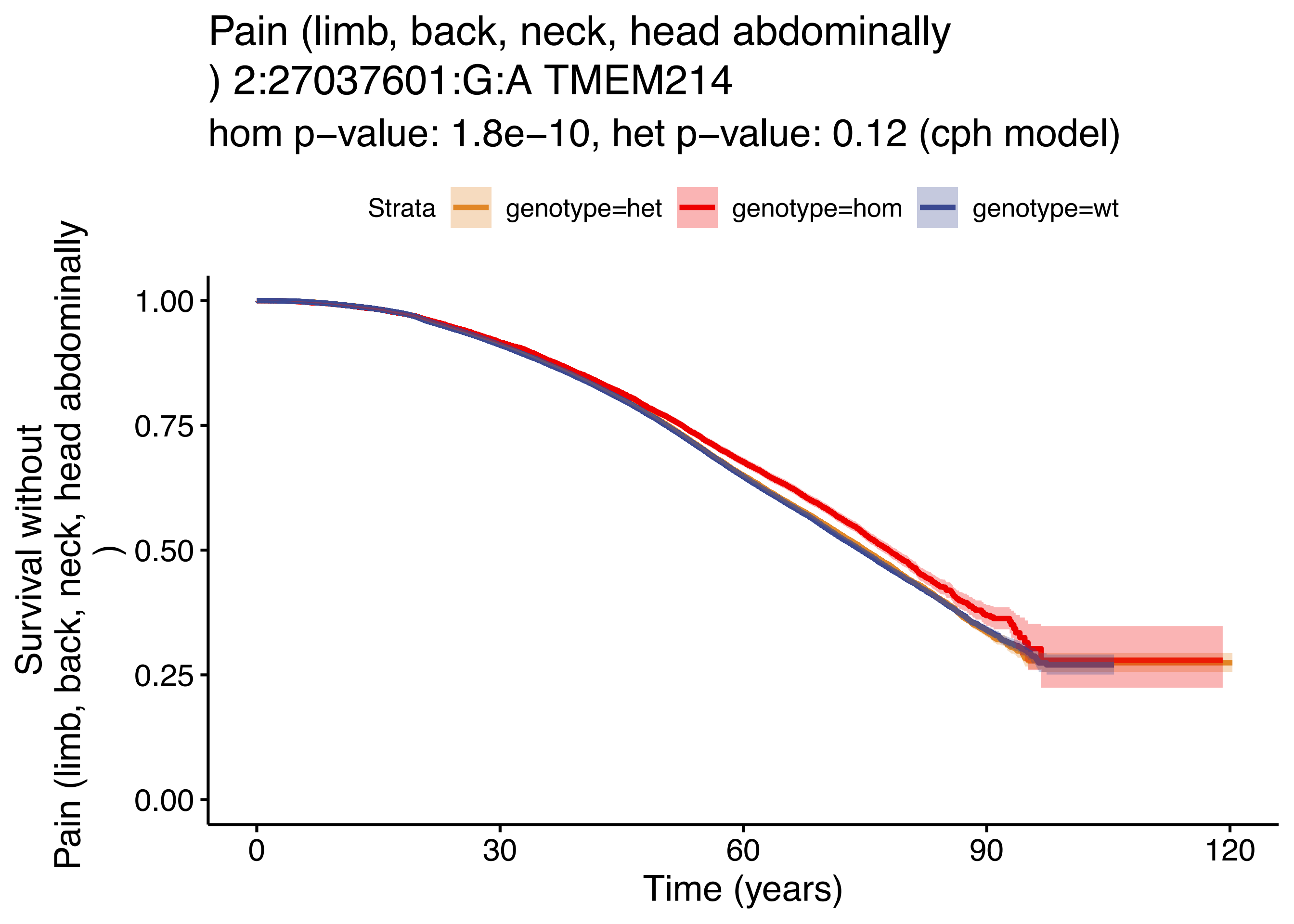

Pain (limb, back, neck, head abdominally
) 2:27037601:G:A TMEM214
hom p−value: 1.8e−10, het p−value: 0.12 (cph model)
Strata
genotype=het
genotype=hom
genotype=wt
Survival without
Pain (limb, back, neck, head abdominally
)
1.00
0.75
0.50
0.25
0.00
0
30
60
90
120
Time (years)

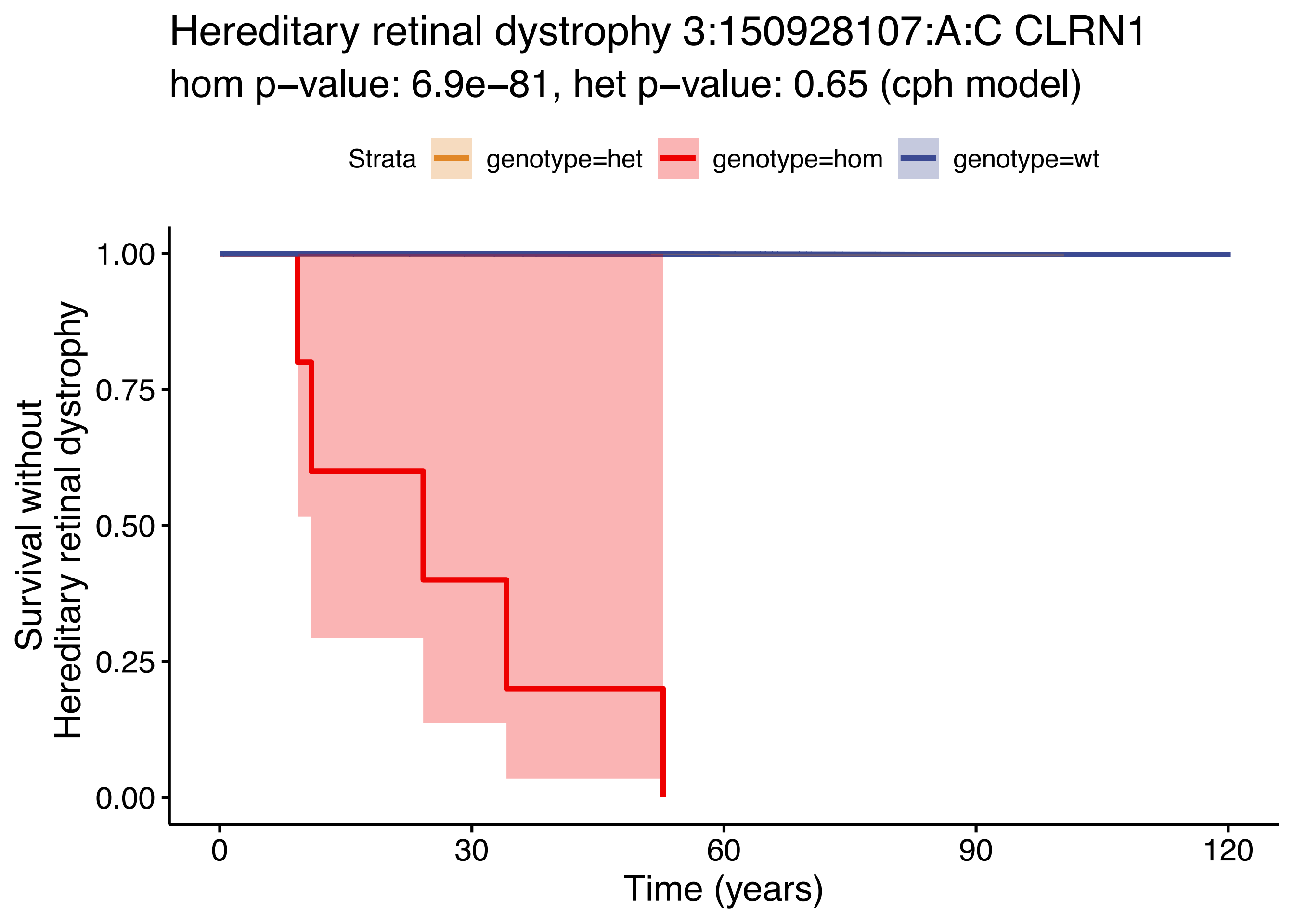

Hereditary retinal dystrophy 3:150928107:A:C CLRN1
hom p−value: 6.9e−81, het p−value: 0.65 (cph model)
Strata
genotype=het
genotype=hom
genotype=wt
Survival without
Hereditary retinal dystrophy
1.00
0.75
0.50
0.25
0.00
0
30
60
90
120
Time (years)

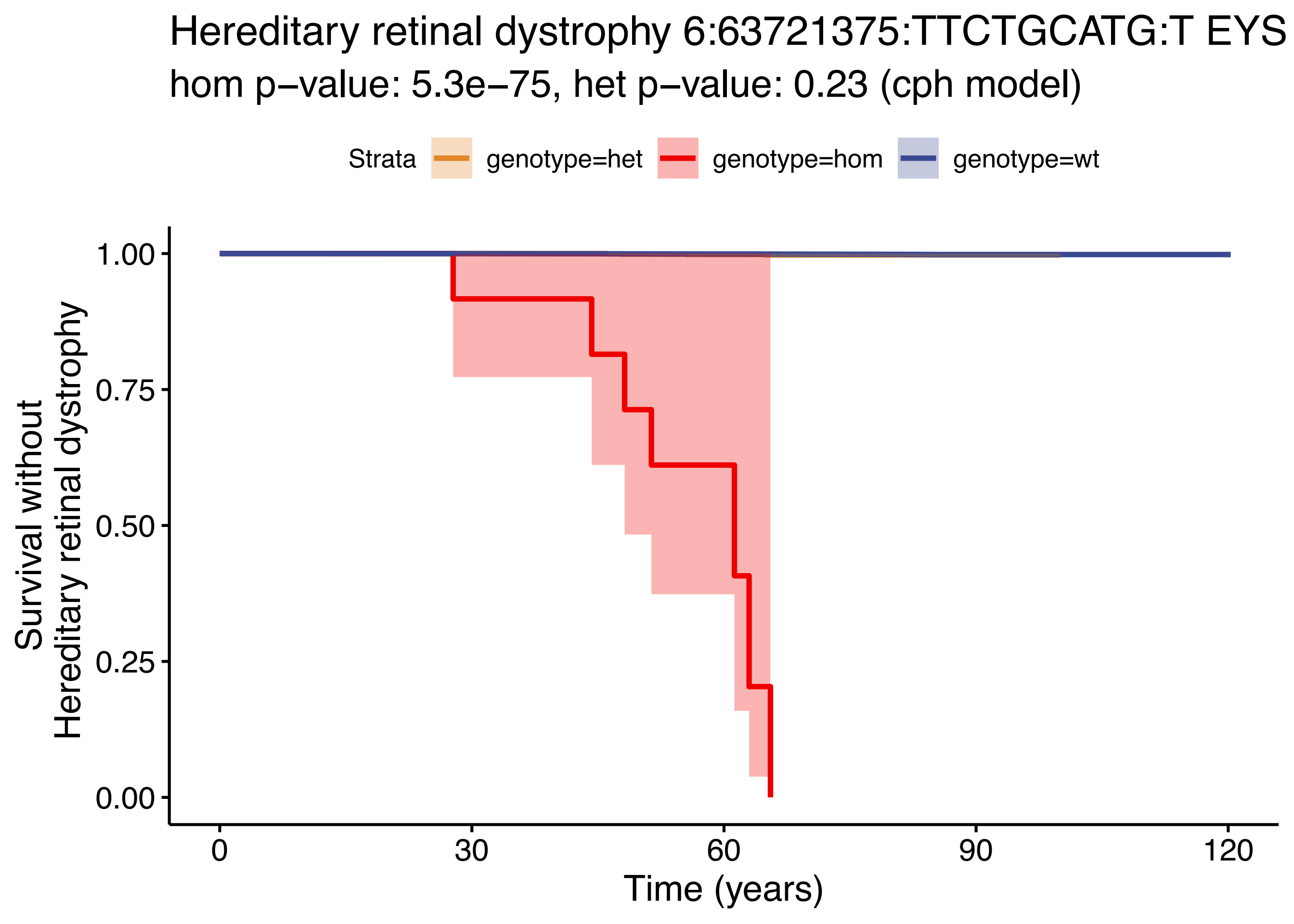
Hereditary retinal dystrophy 6:63721375:TTCTGCATG:T EYS
hom p−value: 5.3e−75, het p−value: 0.23 (cph model)
Strata
genotype=het
genotype=hom
genotype=wt
Survival without
Hereditary retinal dystrophy
1.00
0.75
0.50
0.25
0.00
0
30
60
90
120
Time (years)

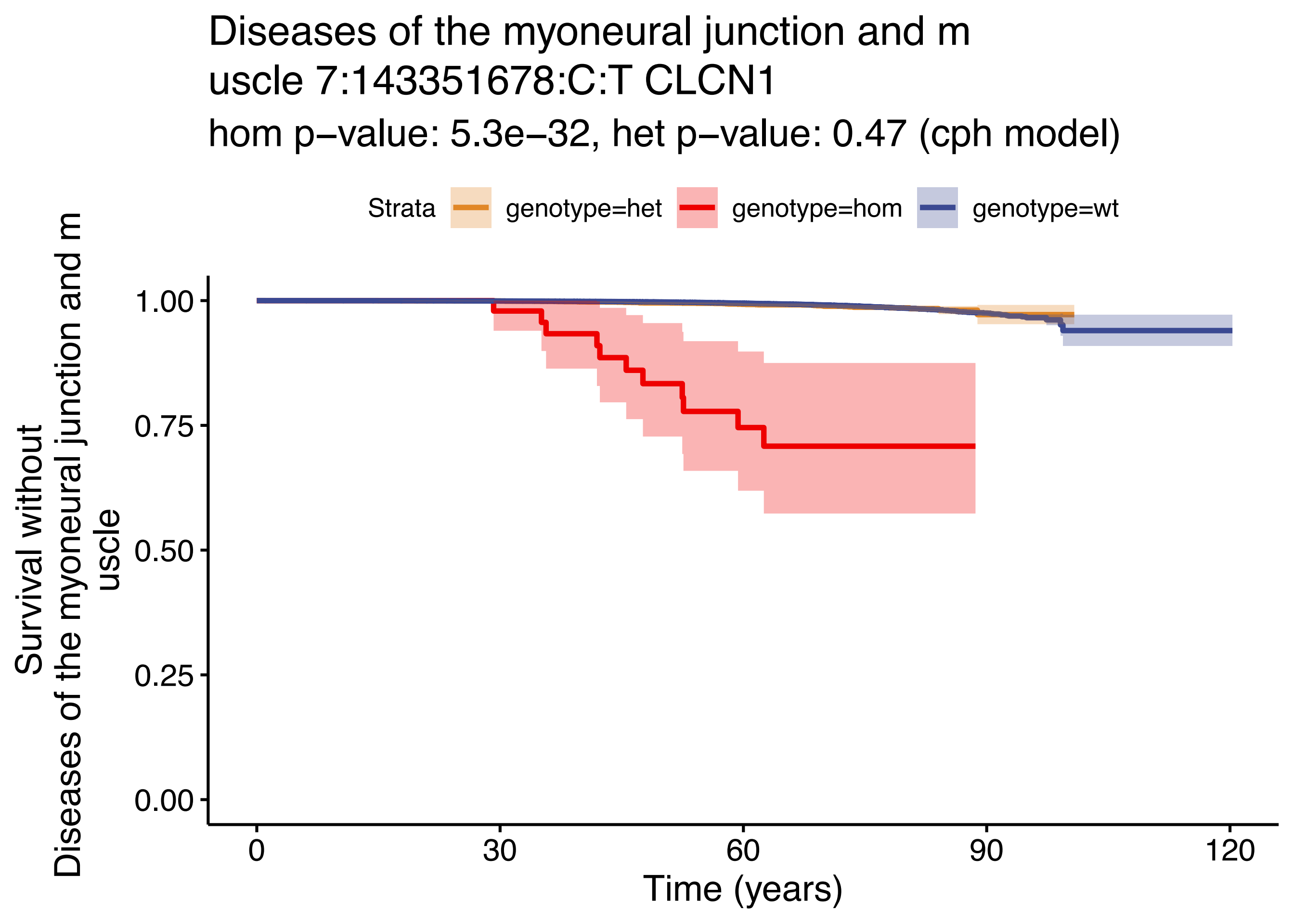

Diseases of the myoneural junction and m
uscle 7:143351678:C:T CLCN1
hom p−value: 5.3e−32, het p−value: 0.47 (cph model)
Strata
genotype=het
genotype=hom
genotype=wt
Survival without
Diseases of the myoneural junction and m
uscle
1.00
0.75
0.50
0.25
0.00
0
30
60
90
120
Time (years)

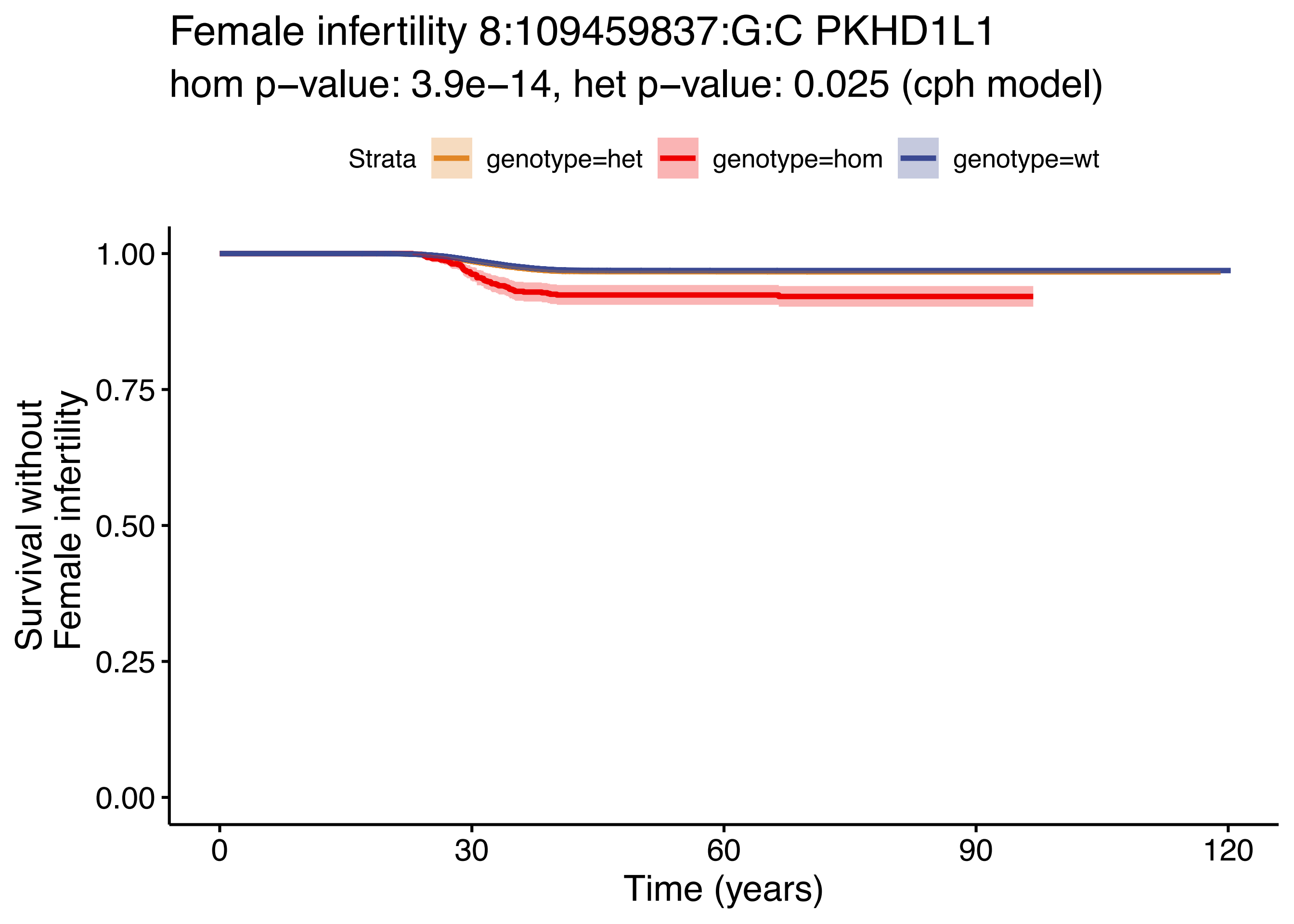

Female infertility 8:109459837:G:C PKHD1L1
hom p−value: 3.9e−14, het p−value: 0.025 (cph model)
Strata
genotype=het
genotype=hom
genotype=wt
Survival without Female infertility
1.00
0.75
0.50
0.25
0.00
0
30
60
90
120
Time (years)

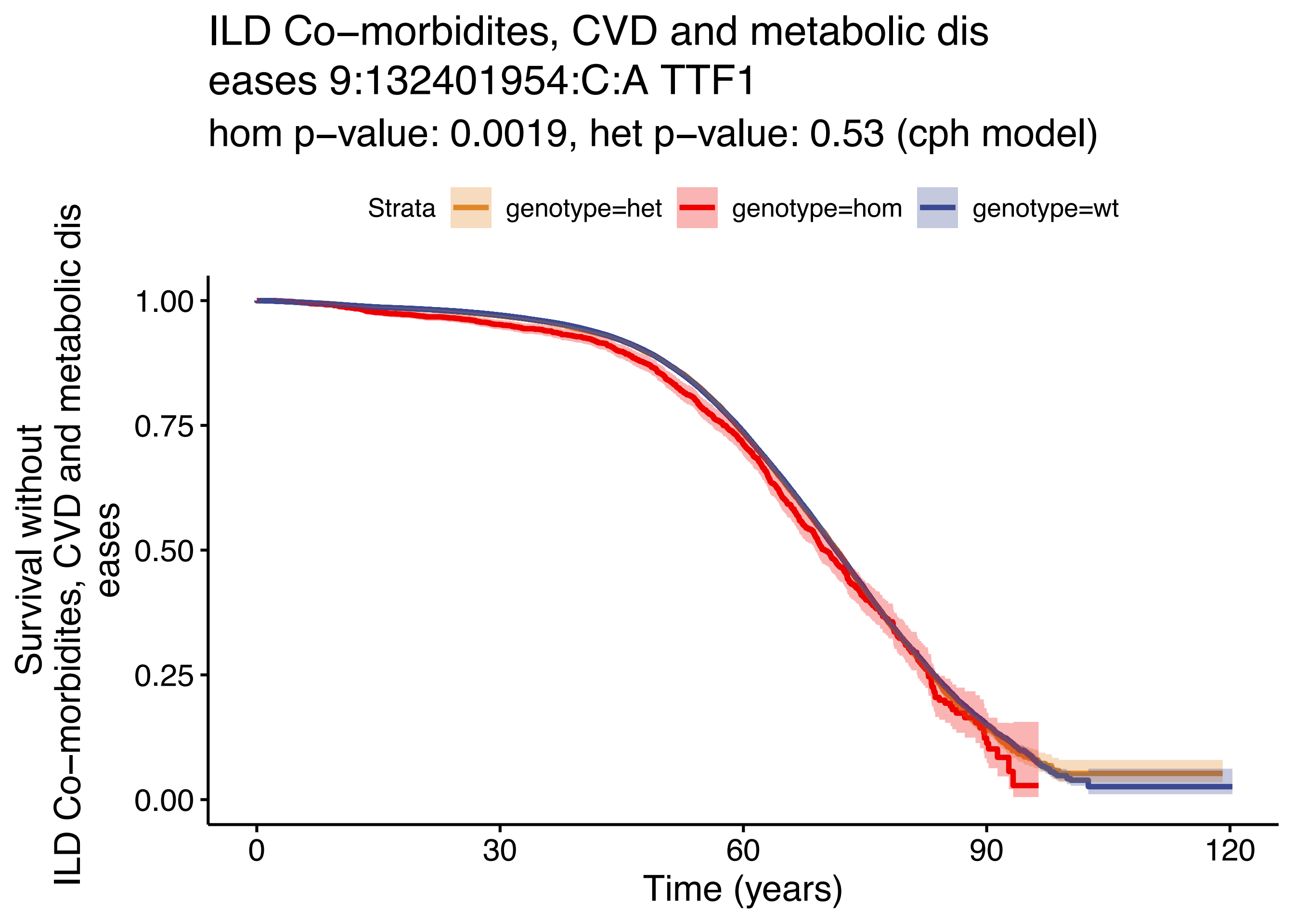

ILD Co−morbidites, CVD and metabolic dis
eases 9:132401954:C:A TTF1
hom p−value: 0.0019, het p−value: 0.53 (cph model)
Strata
genotype=het
genotype=hom
genotype=wt
Survival without
ILD Co−morbidites, CVD and metabolic dis
eases
1.00
0.75
0.50
0.25
0.00
0
30
60
90
120
Time (years)
