## Supplementary Figure S2A-AG for "Mono- and bi-allelic effects of coding variants on disease in 176,899 Finns"

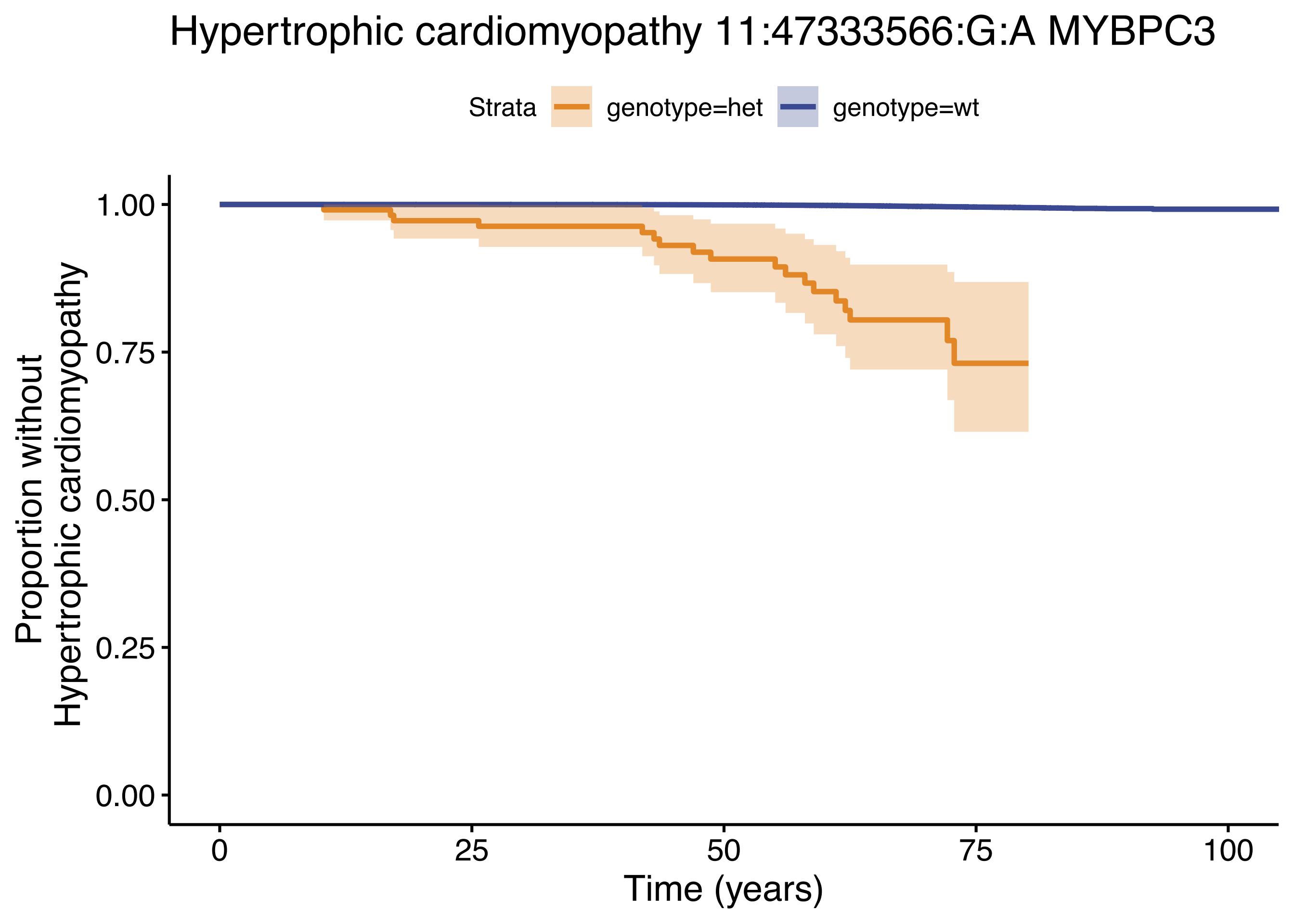

Hypertrophic cardiomyopathy 11:47333566:G:A MYBPC3
Strata
genotype=het
genotype=wt
Proportion without
Hypertrophic cardiomyopathy
1.00
0.75
0.50
0.25
0.00
0
25
50
75
100
Time (years)

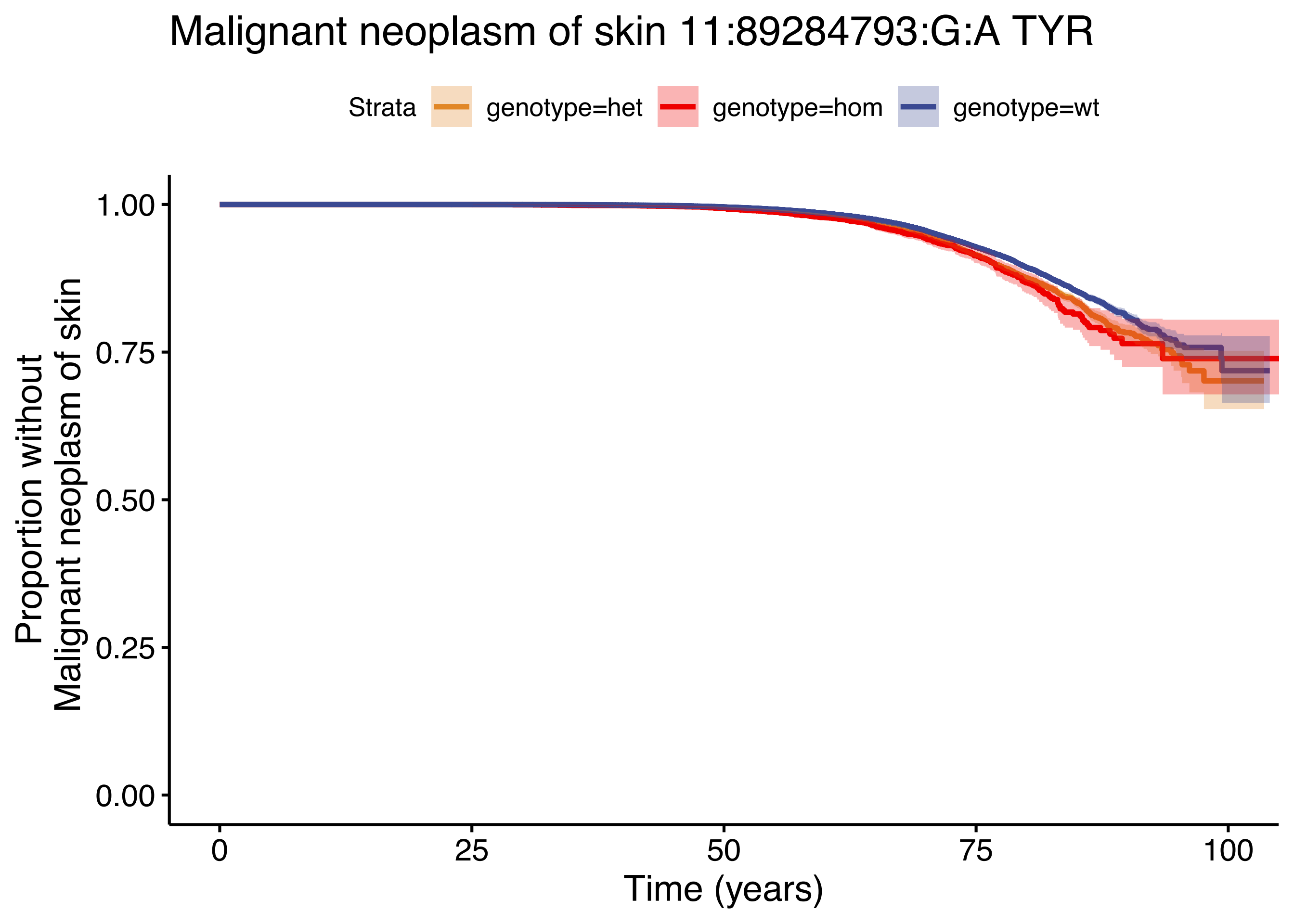
Malignant neoplasm of skin 11:89284793:G:A TYR
Strata
genotype=het
genotype=hom
genotype=wt
Proportion without
Malignant neoplasm of skin
1.00
0.75
0.50
0.25
0.00
0
25
50
75
100
Time (years)

### Cholelithiasis, broad definition wi
### th cholecystitis 14:94378610:C:T SERPINA1

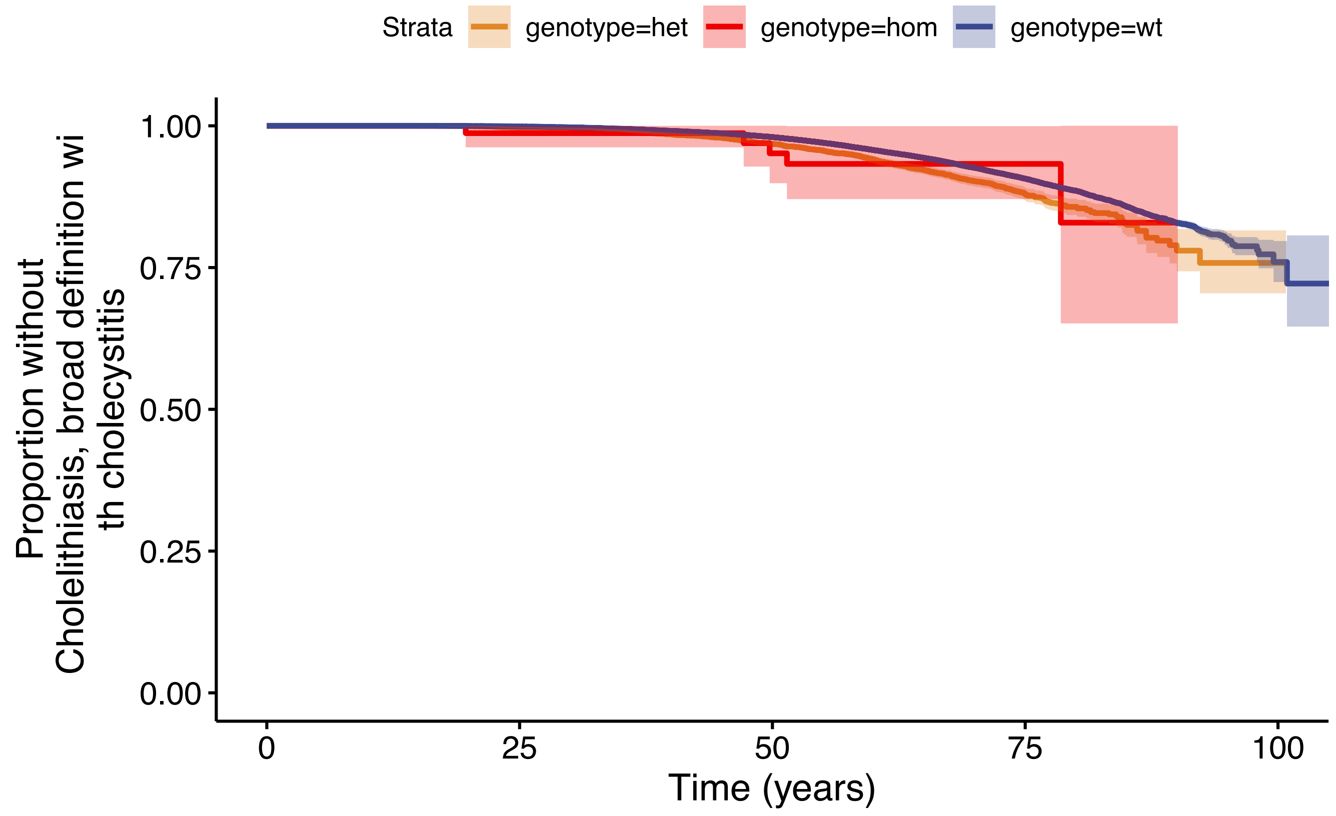

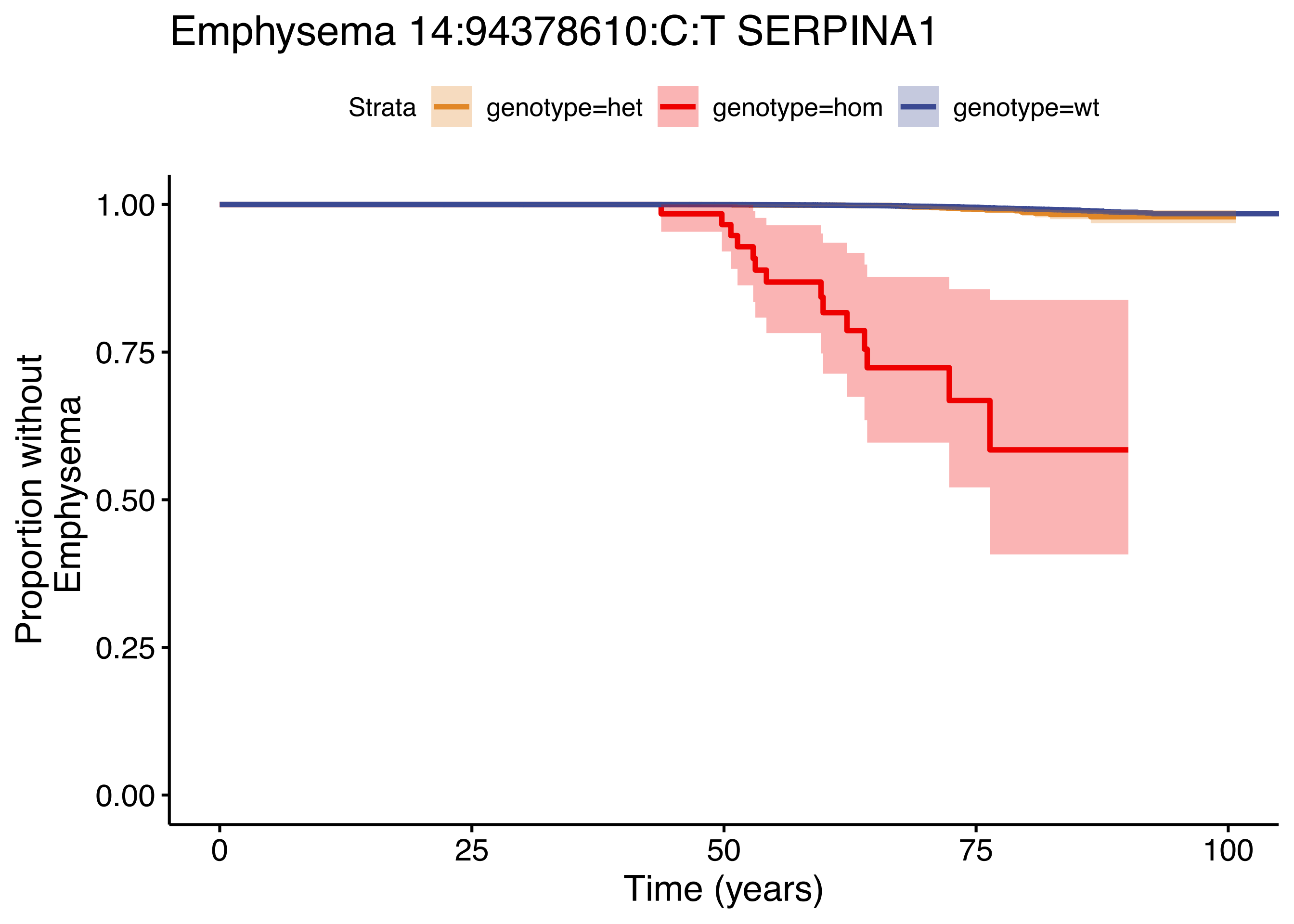

Emphysema 14:94378610:C:T SERPINA1
Strata
genotype=het
genotype=hom
genotype=wt
Proportion without
Emphysema
1.00
0.75
0.50
0.25
0.00
0
25
50
75
100
Time (years)

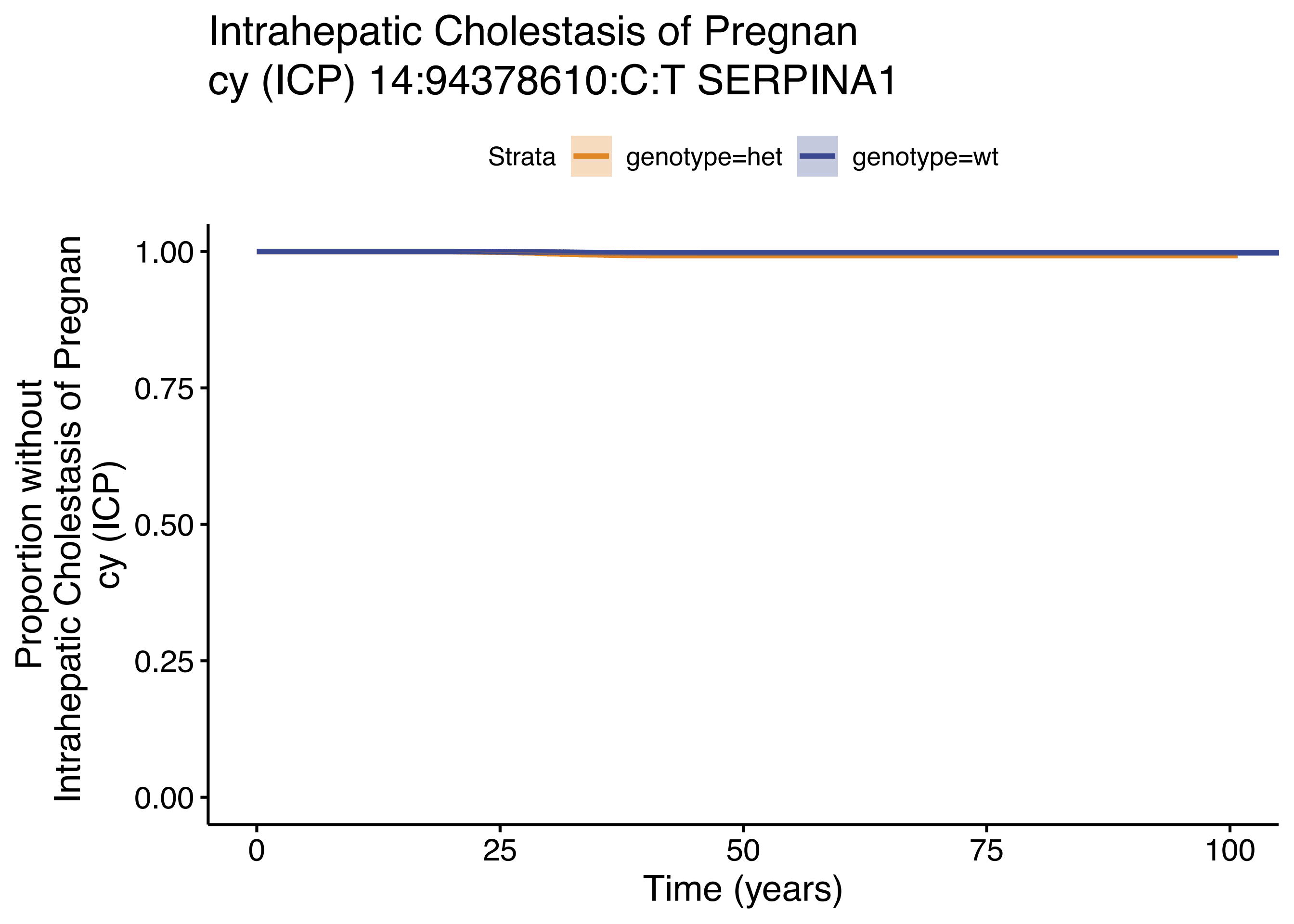
Intrahepatic Cholestasis of Pregnan
cy (ICP) 14:94378610:C:T SERPINA1
Strata
genotype=het
genotype=wt
Proportion without
Intrahepatic Cholestasis of Pregnan
cy (ICP)
1.00
0.75
0.50
0.25
0.00
0
25
50
75
100
Time (years)

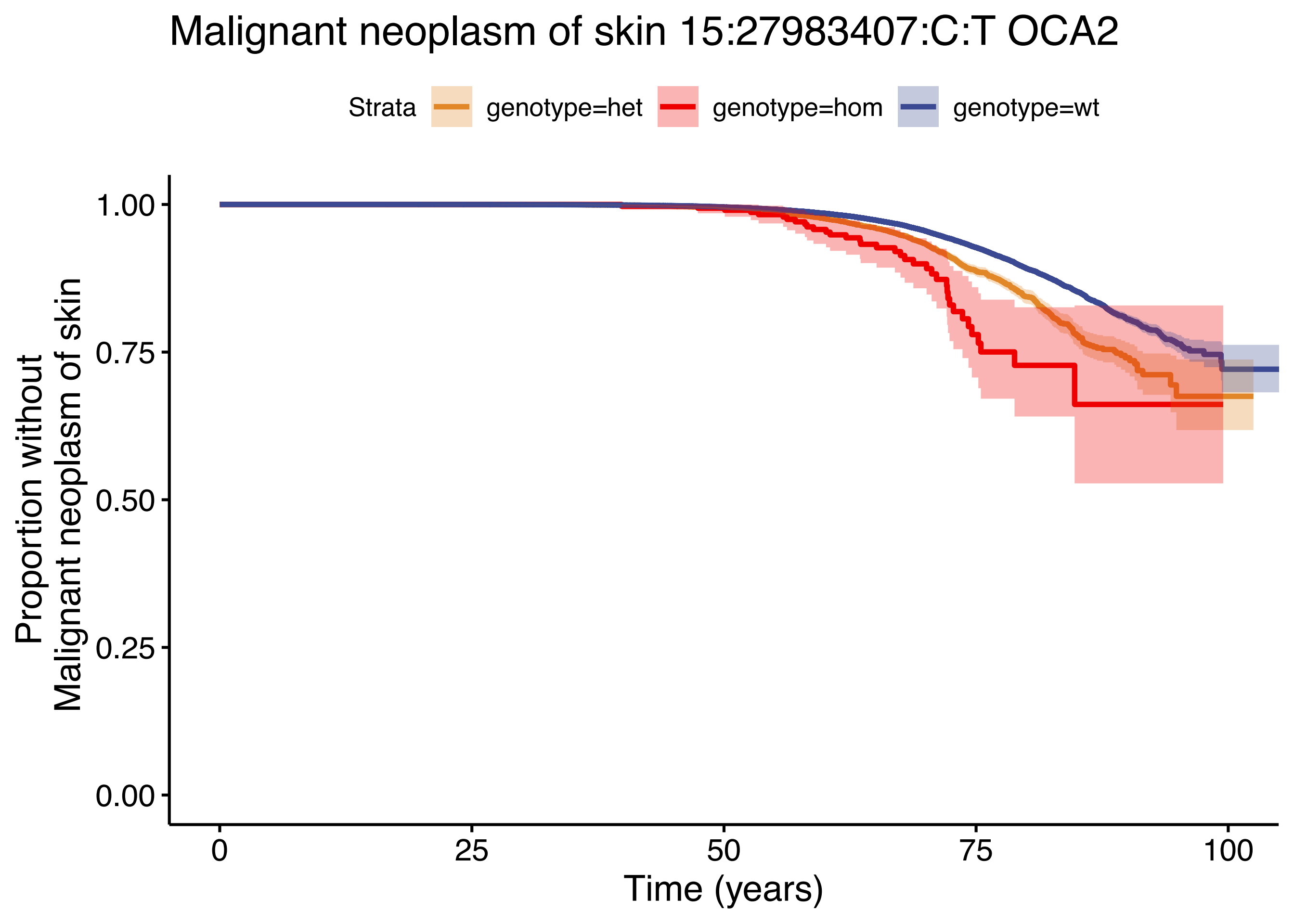

Malignant neoplasm of skin 15:27983407:C:T OCA2
Strata
genotype=het
genotype=hom
genotype=wt
Proportion without
Malignant neoplasm of skin
1.00
0.75
0.50
0.25
0.00
0
25
50
75
100
Time (years)

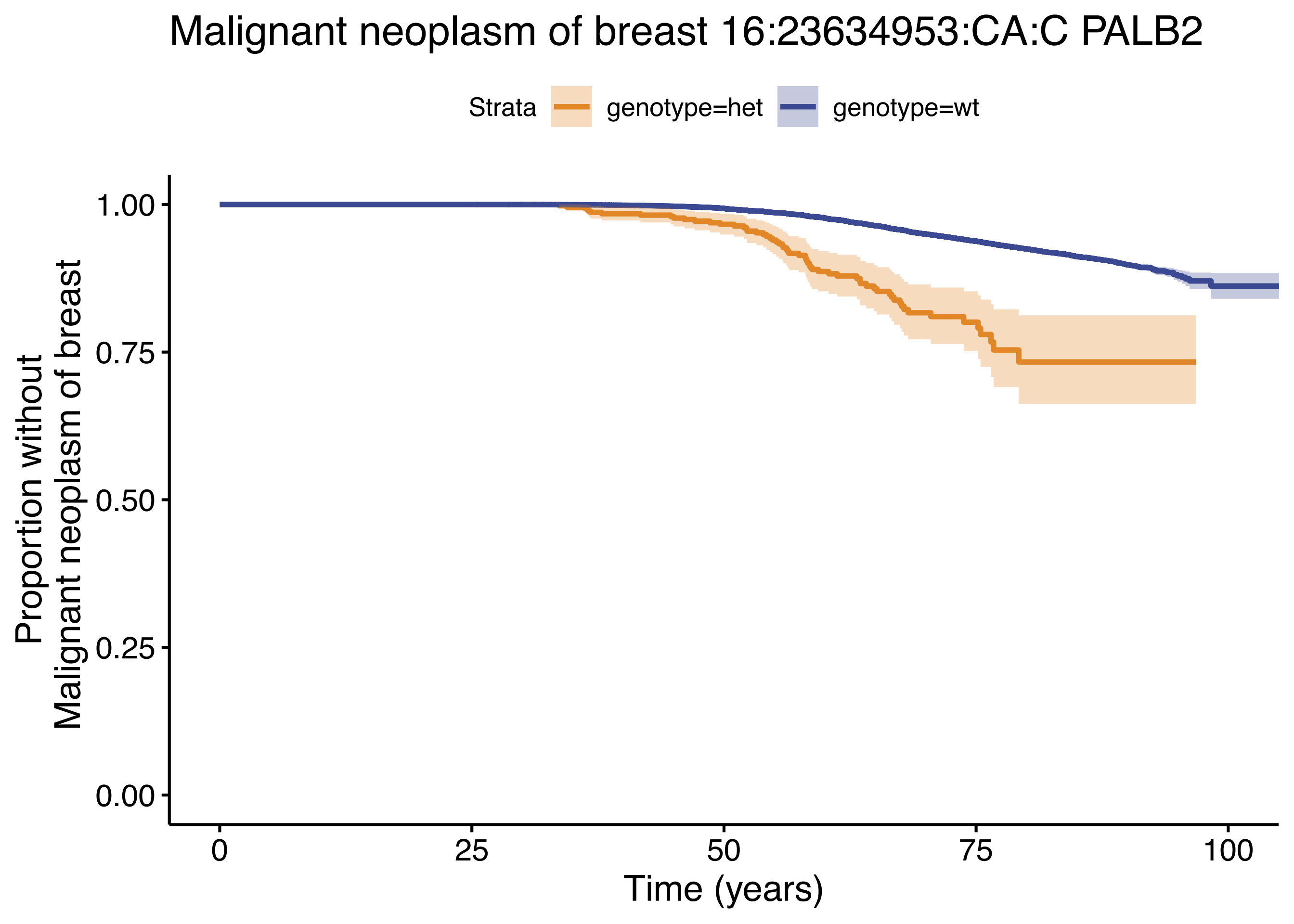

Malignant neoplasm of breast 16:23634953:CA:C PALB2
Strata
genotype=het
genotype=wt
Proportion without
Malignant neoplasm of breast
1.00
0.75
0.50
0.25
0.00
0
25
50
75
100
Time (years)

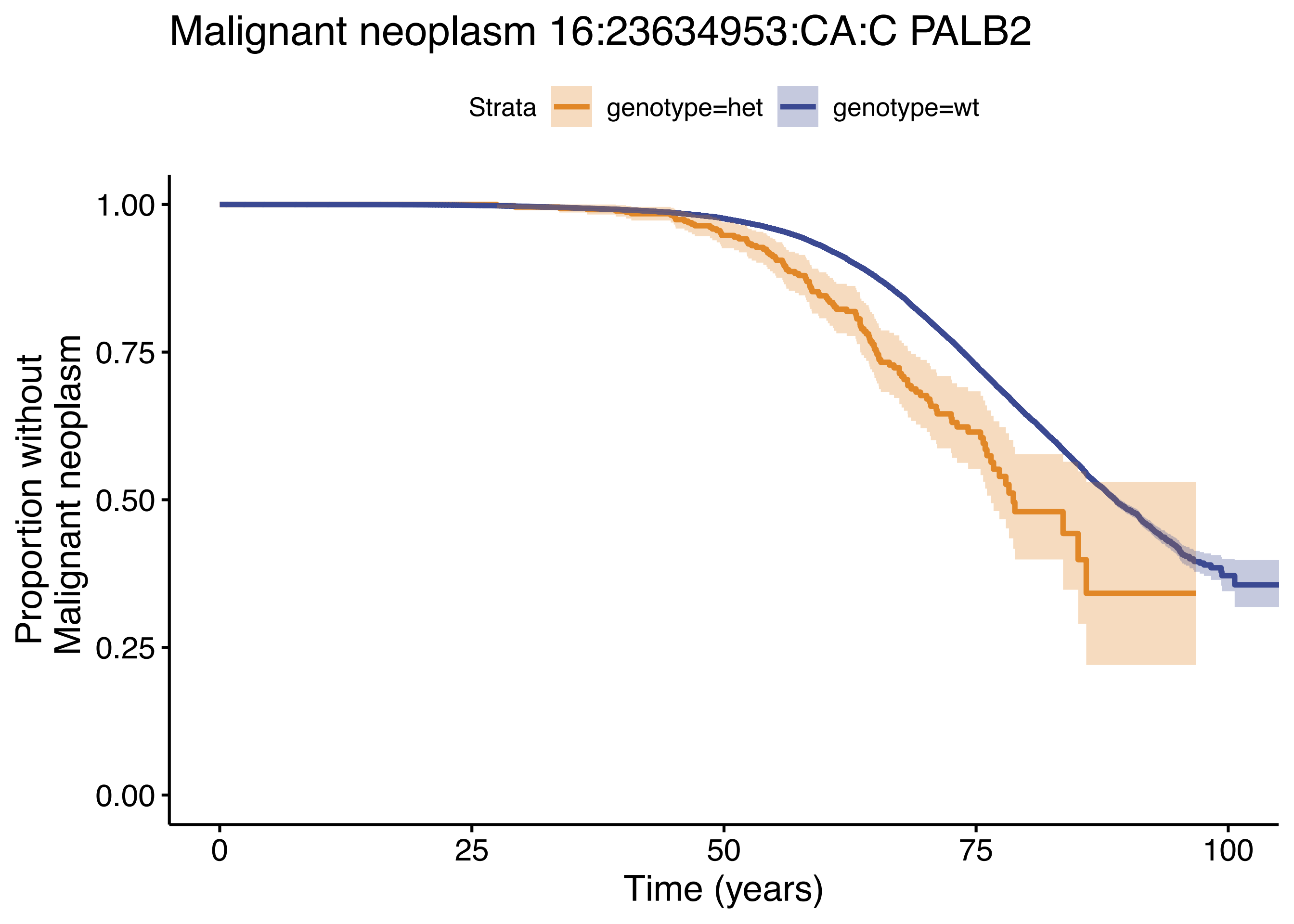

Malignant neoplasm 16:23634953:CA:C PALB2
Strata
genotype=het
genotype=wt
Proportion without Malignant neoplasm
1.00
0.75
0.50
0.25
0.00
0
25
50
75
100
Time (years)

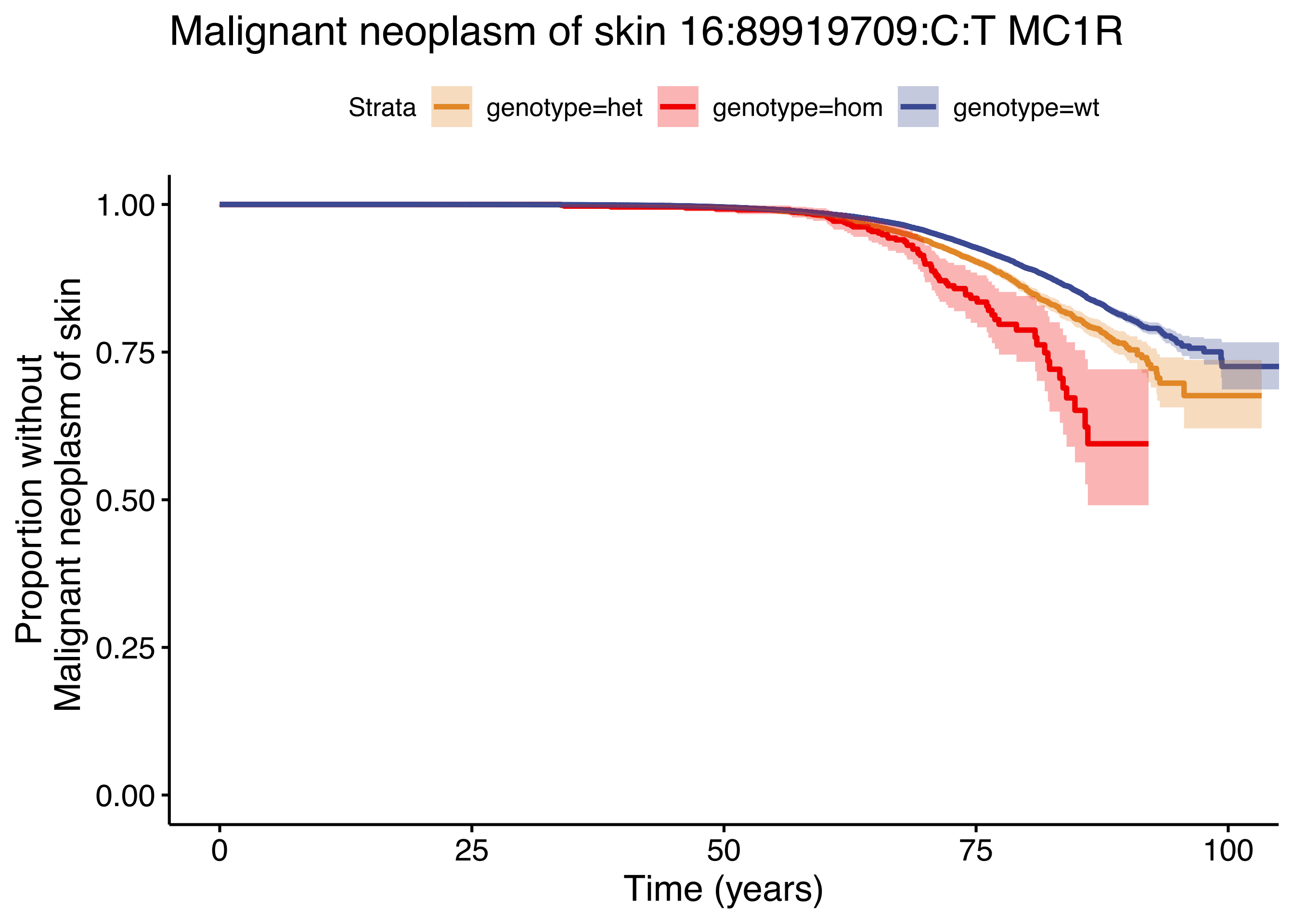

Malignant neoplasm of skin 16:89919709:C:T MC1R
Strata
genotype=het
genotype=hom
genotype=wt
Proportion without
Malignant neoplasm of skin
1.00
0.75
0.50
0.25
0.00
0
25
50
75
100
Time (years)

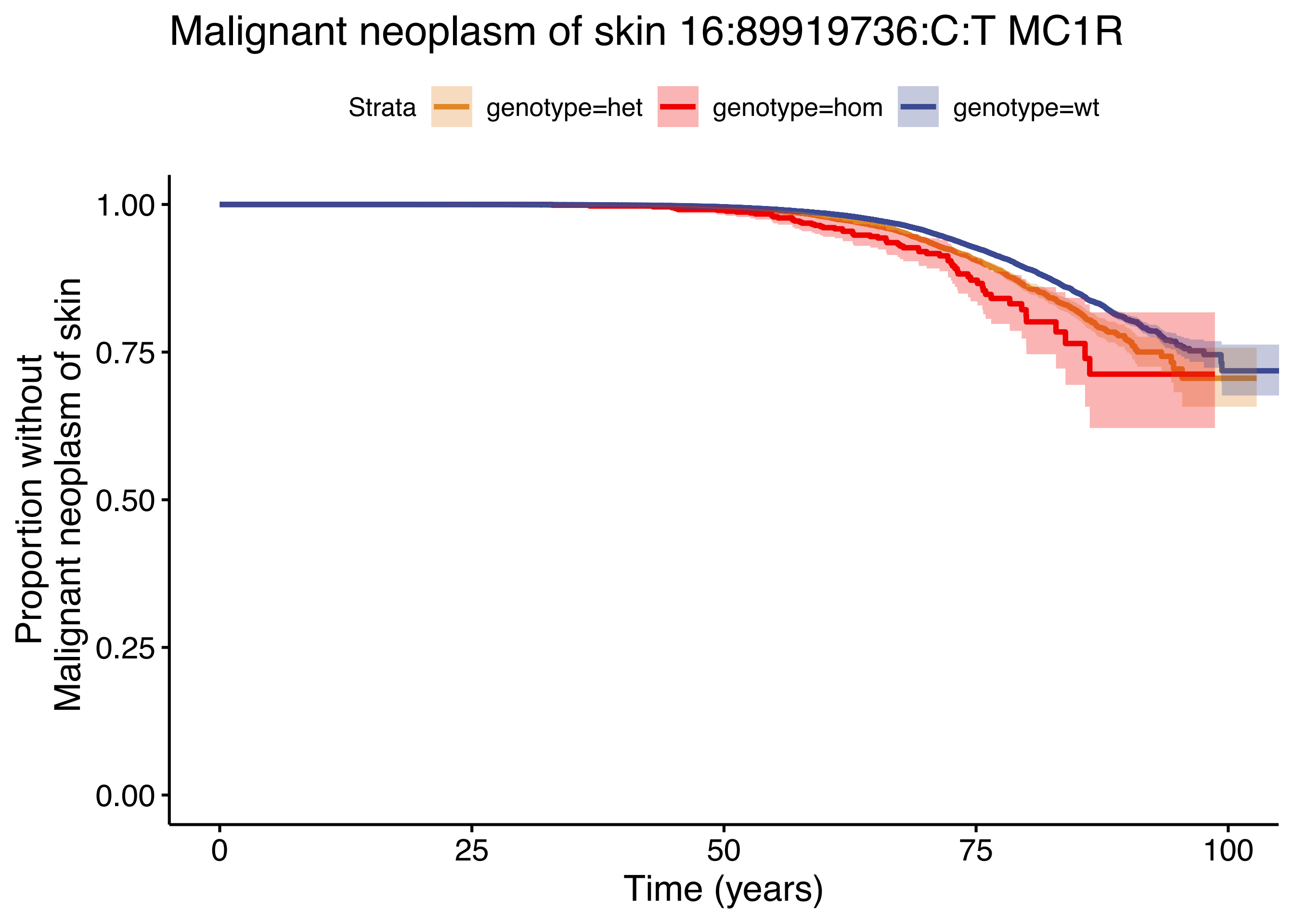
Malignant neoplasm of skin 16:89919736:C:T MC1R
Strata
genotype=het
genotype=hom
genotype=wt
Proportion without
Malignant neoplasm of skin
1.00
0.75
0.50
0.25
0.00
0
25
50
75
100
Time (years)

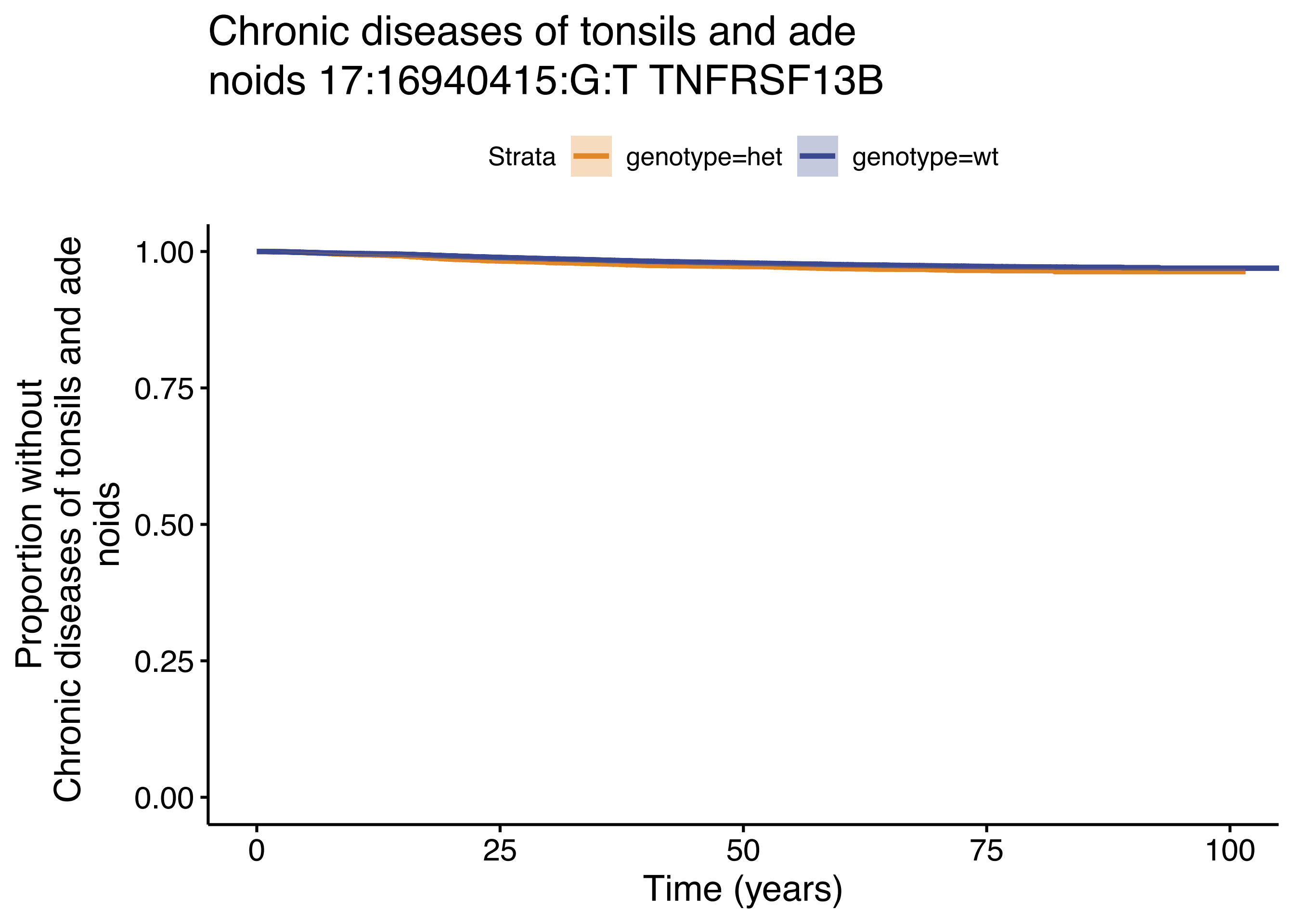
Chronic diseases of tonsils and ade
noids 17:16940415:G:T TNFRSF13B
Strata
genotype=het
genotype=wt
Proportion without
Chronic diseases of tonsils and ade
noids
1.00
0.75
0.50
0.25
0.00
0
25
50
75
100
Time (years)

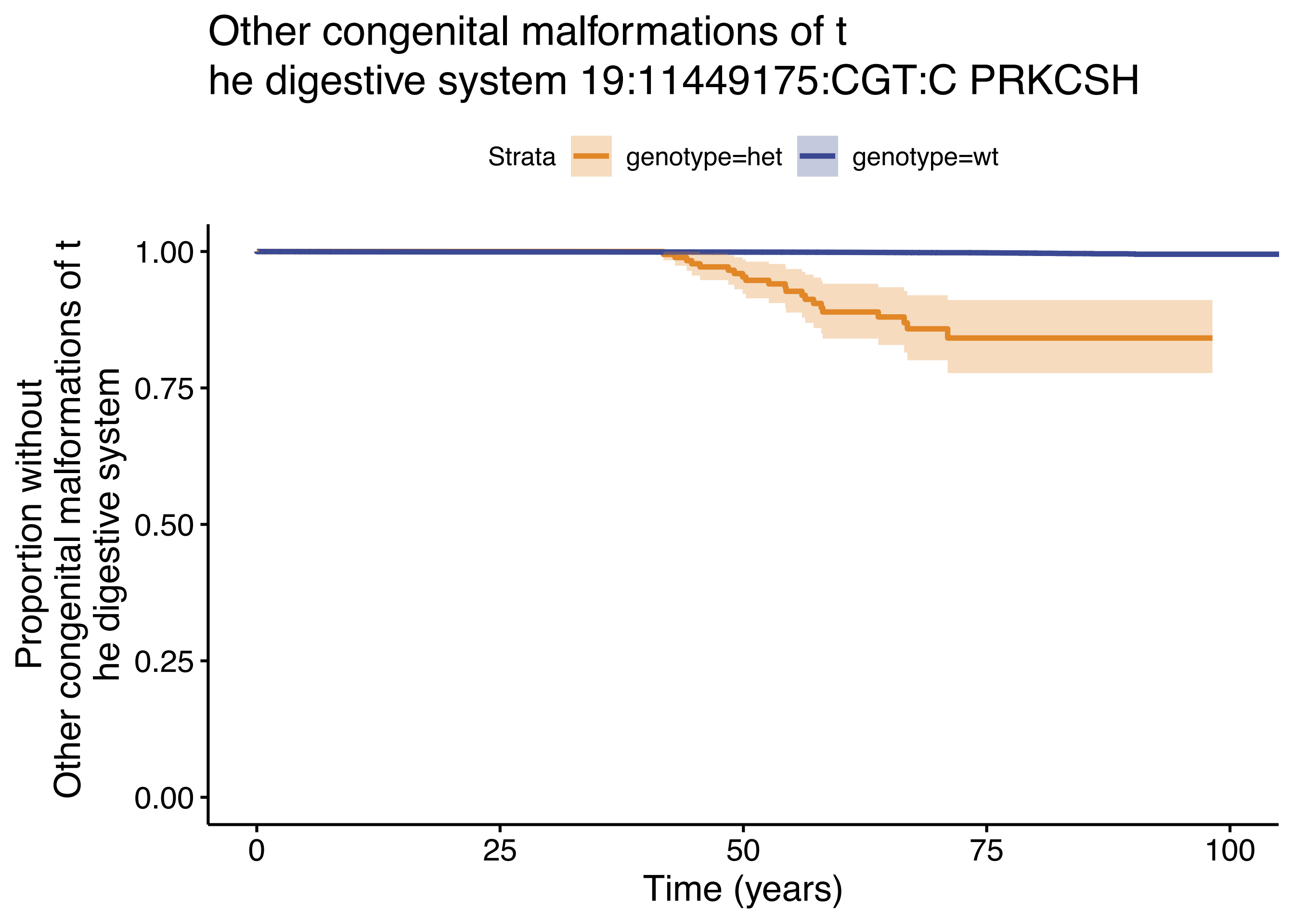

Other congenital malformations of t
he digestive system 19:11449175:CGT:C PRKCSH
Strata
genotype=het
genotype=wt
Proportion without
Other congenital malformations of t
he digestive system
1.00
0.75
0.50
0.25
0.00
0
25
50
75
100
Time (years)

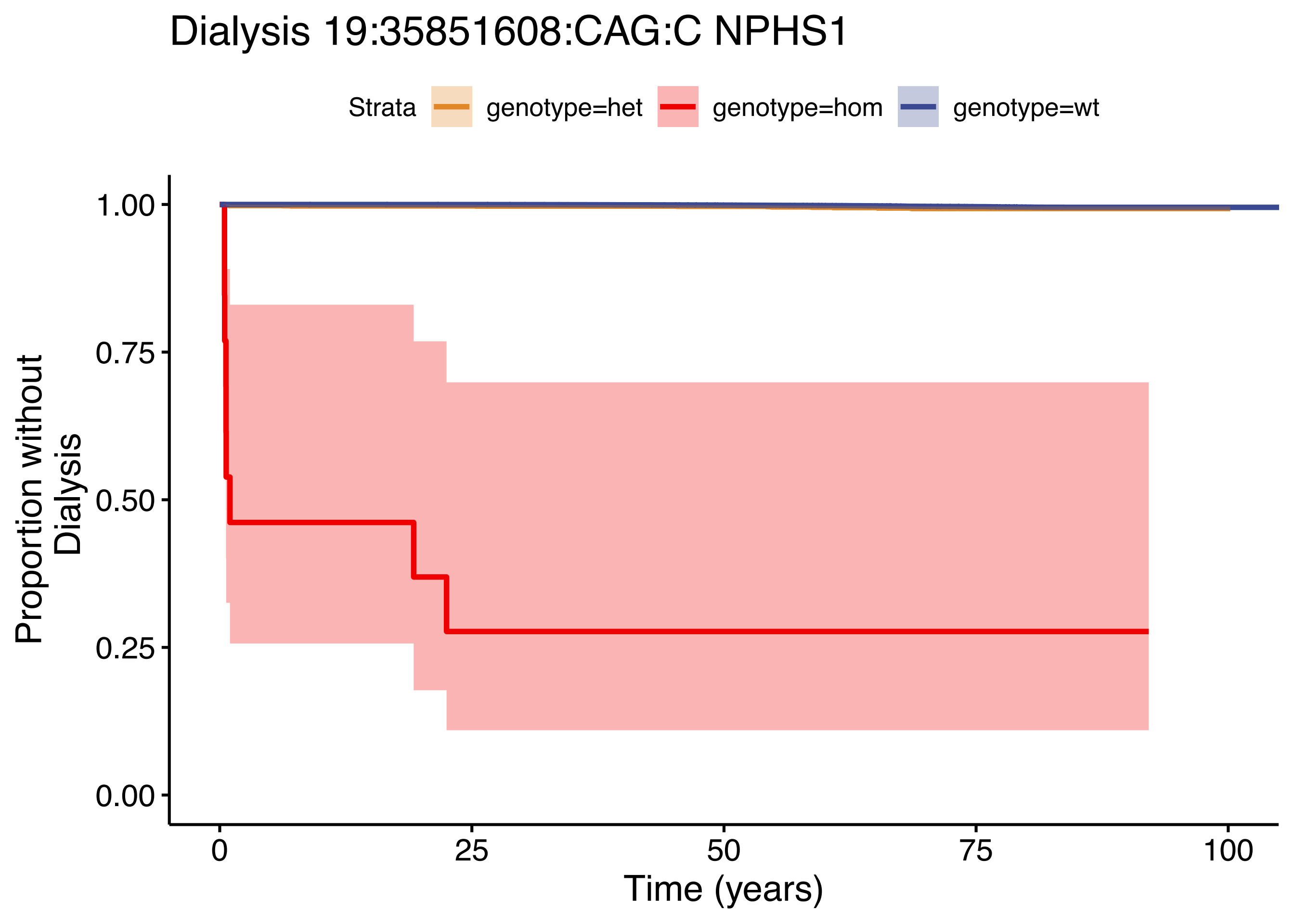

Dialysis 19:35851608:CAG:C NPHS1
Strata
genotype=het
genotype=hom
genotype=wt
Proportion without Dialysis
1.00
0.75
0.50
0.25
0.00
0
25
50
75
100
Time (years)

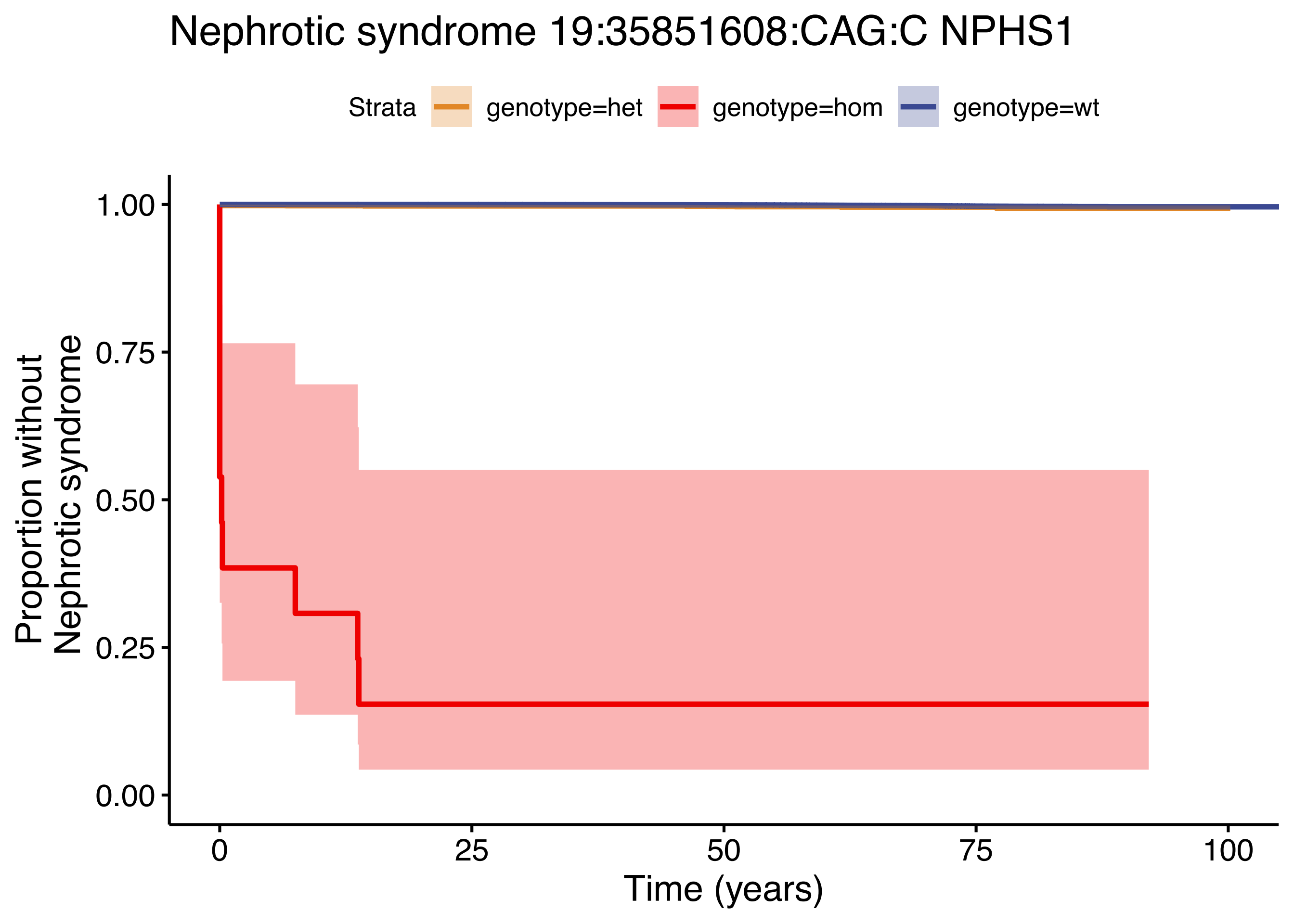

Nephrotic syndrome 19:35851608:CAG:C NPHS1
Strata
genotype=het
genotype=hom
genotype=wt
Proportion without
Nephrotic syndrome
1.00
0.75
0.50
0.25
0.00
0
25
50
75
100
Time (years)

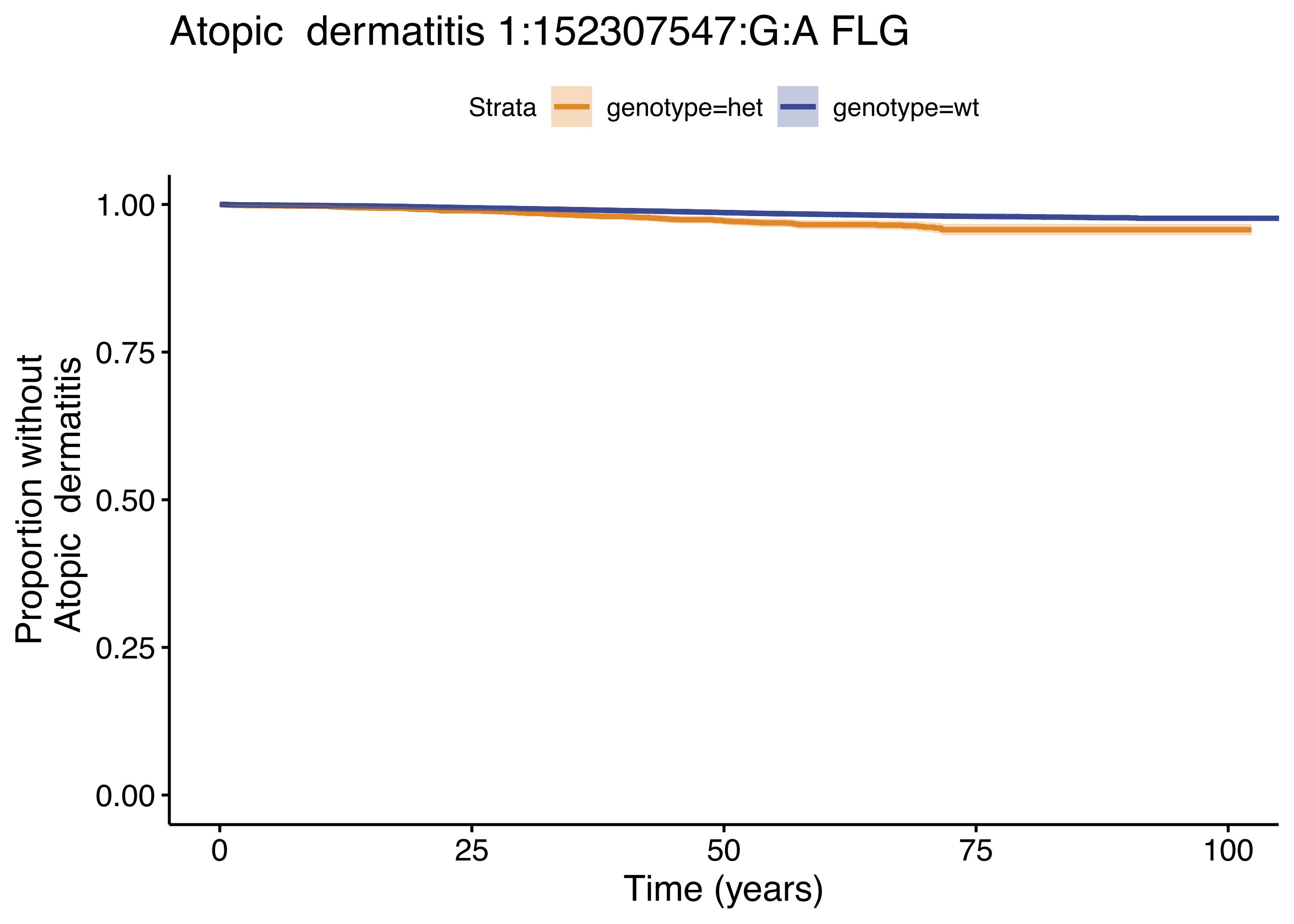

Atopic dermatitis 1:152307547:G:A FLG
Strata
genotype=het
genotype=wt
1.00
0.75
0.50
0.25
0.00
Proportion without
Atopic dermatitis
0
25
50
75
100
Time (years)

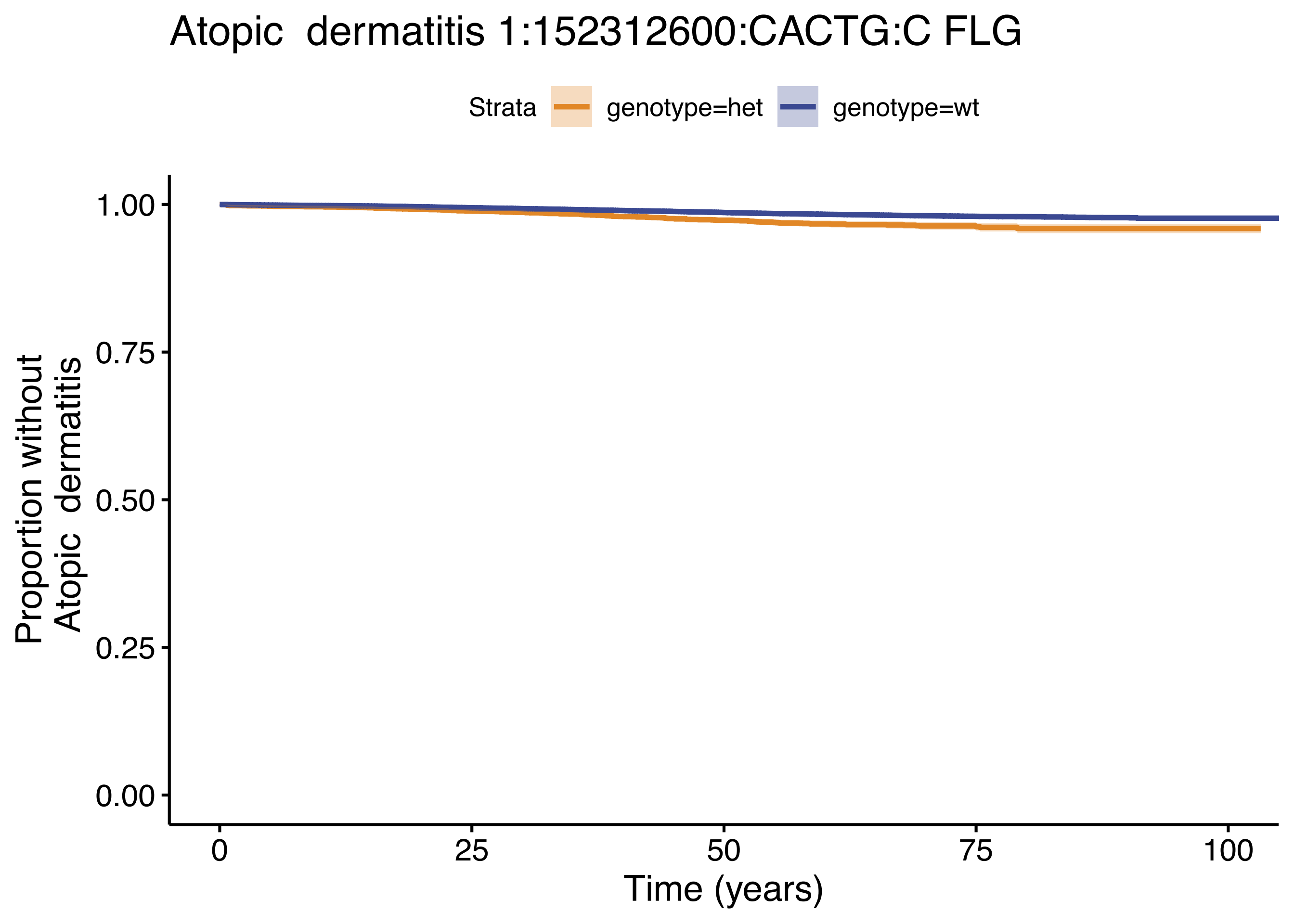

Atopic dermatitis 1:152312600:CACTG:C FLG
Strata
genotype=het
genotype=wt
Proportion without
Atopic dermatitis
1.00
0.75
0.50
0.25
0.00
0
25
50
75
100
Time (years)

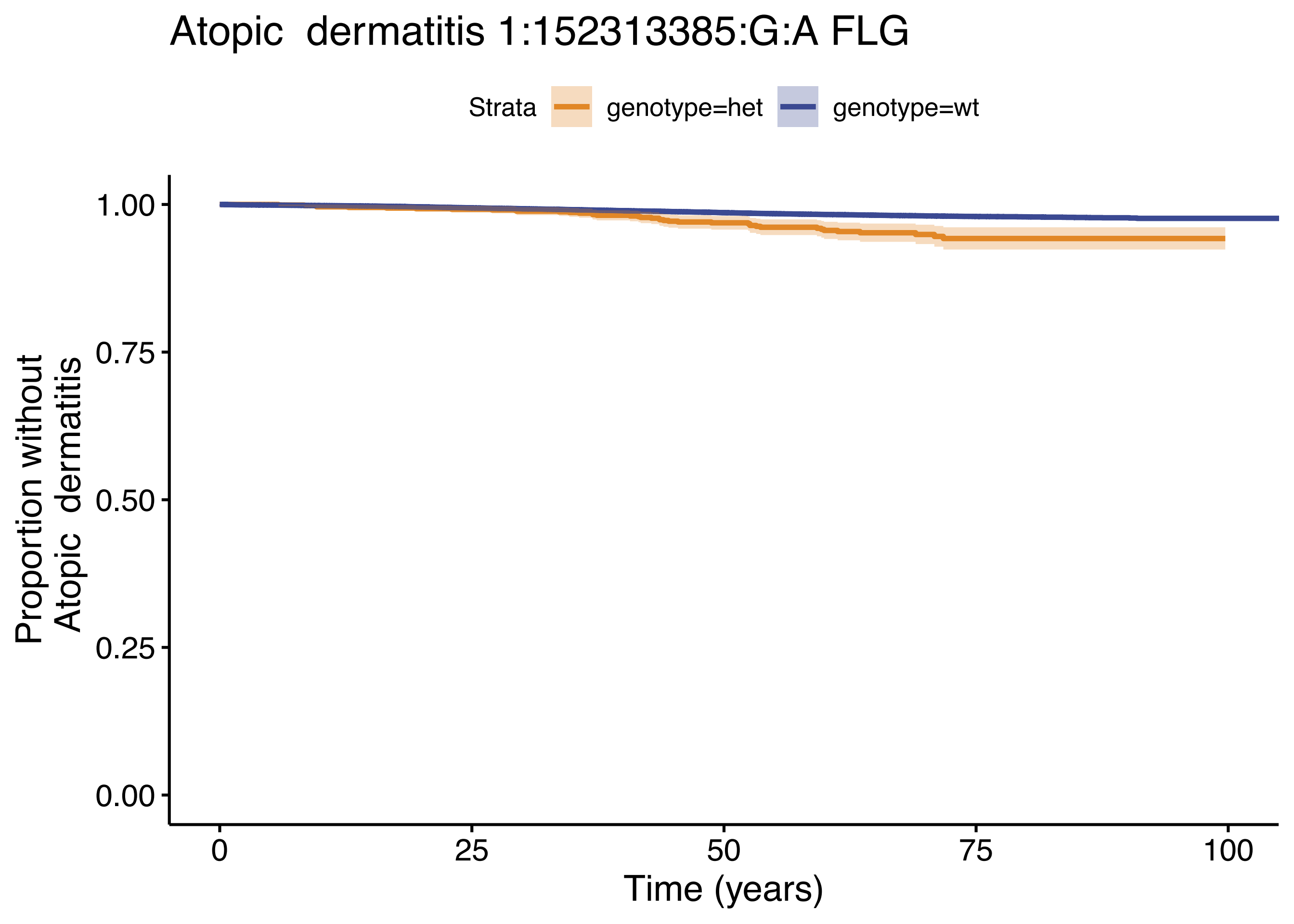

Atopic dermatitis 1:152313385:G:A FLG
Strata
genotype=het
genotype=wt
Proportion without
Atopic dermatitis
1.00
0.75
0.50
0.25
0.00
0
25
50
75
100
Time (years)

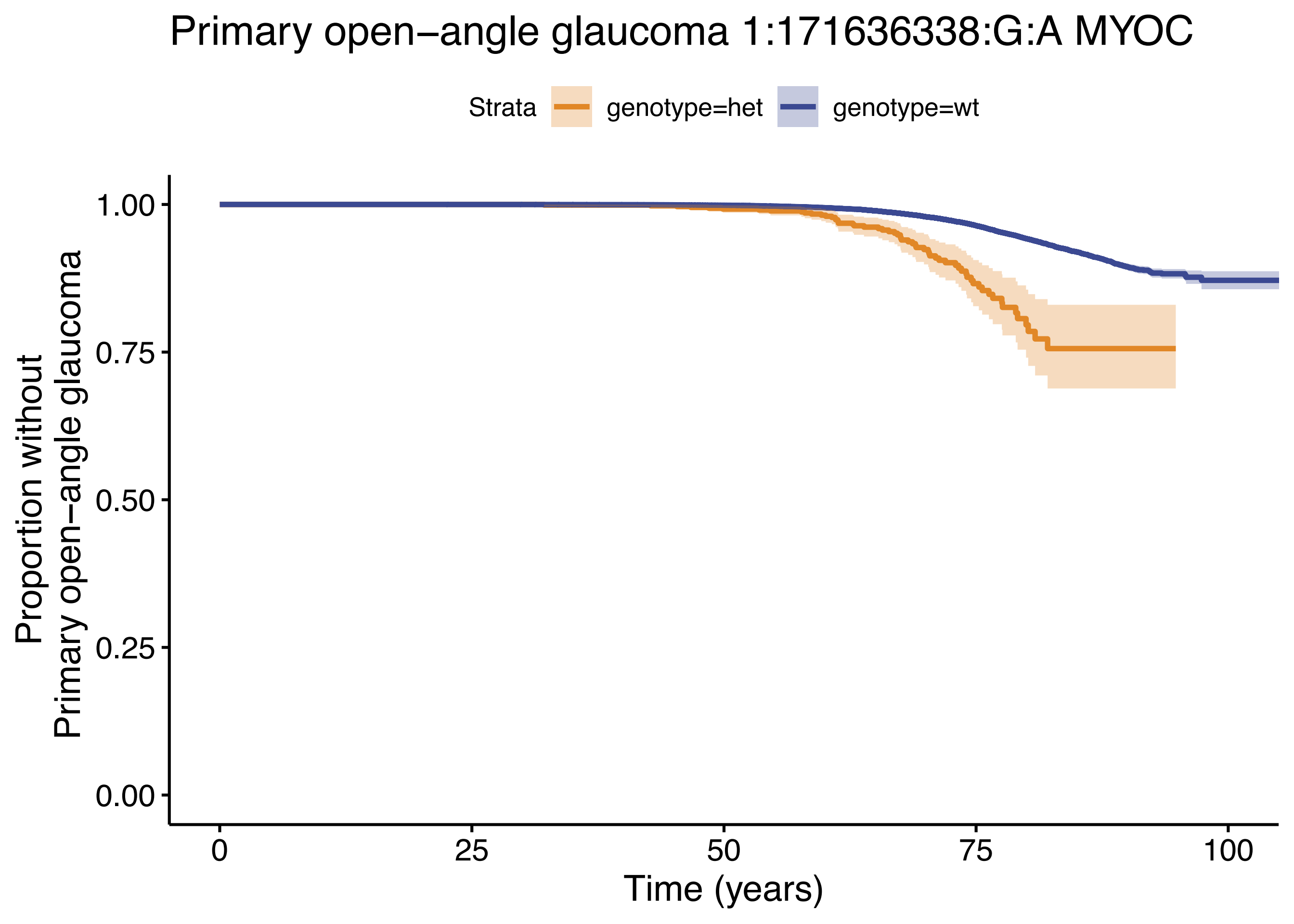

Primary open−angle glaucoma 1:171636338:G:A MYOC
Strata
genotype=het
genotype=wt
Proportion without
Primary open−angle glaucoma
1.00
0.75
0.50
0.25
0.00
0
25
50
75
100
Time (years)

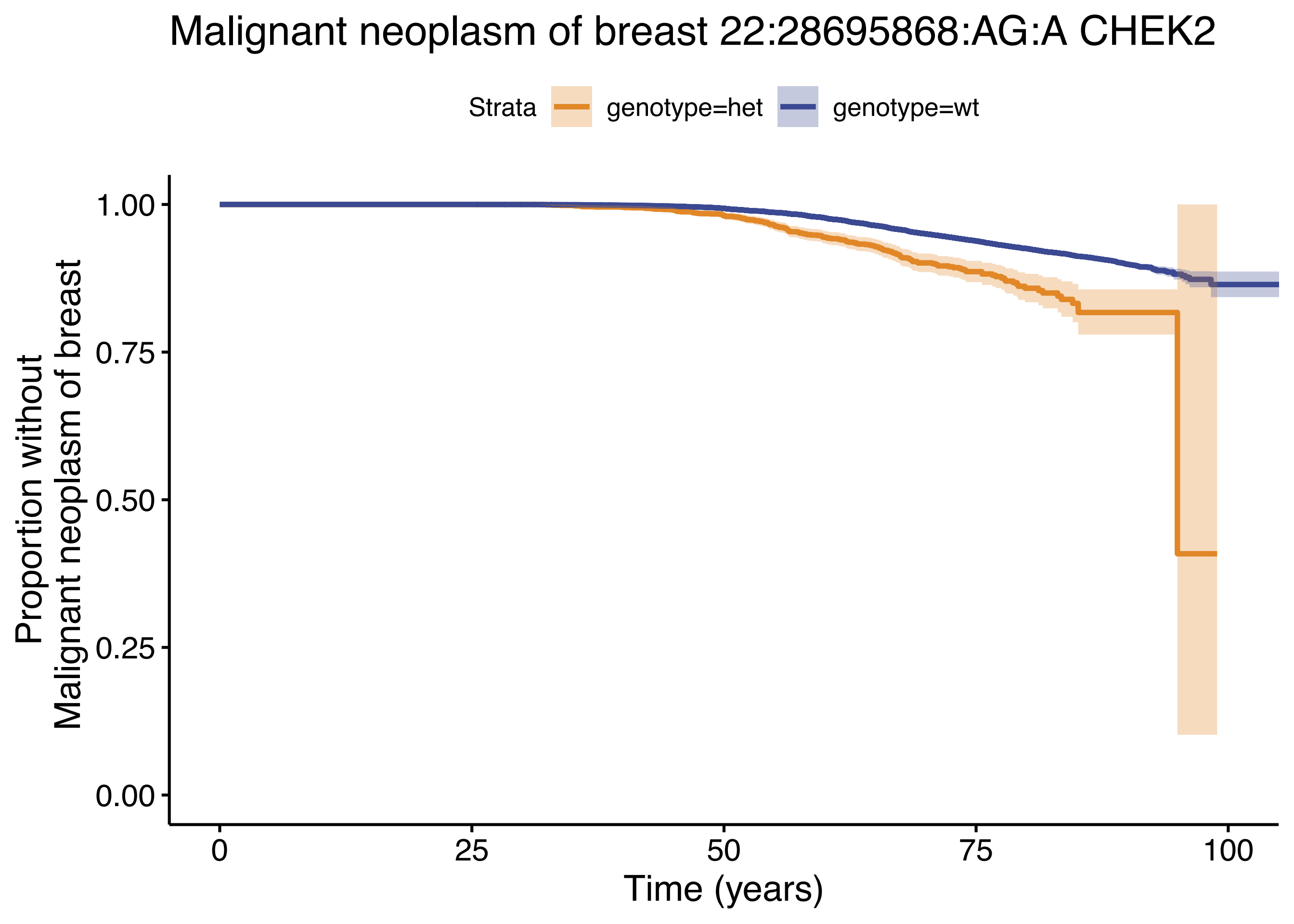
Malignant neoplasm of breast 22:28695868:AG:A CHEK2
Strata
genotype=het
genotype=wt
Proportion without
Malignant neoplasm of breast
1.00
0.75
0.50
0.25
0.00
0
25
50
75
100
Time (years)

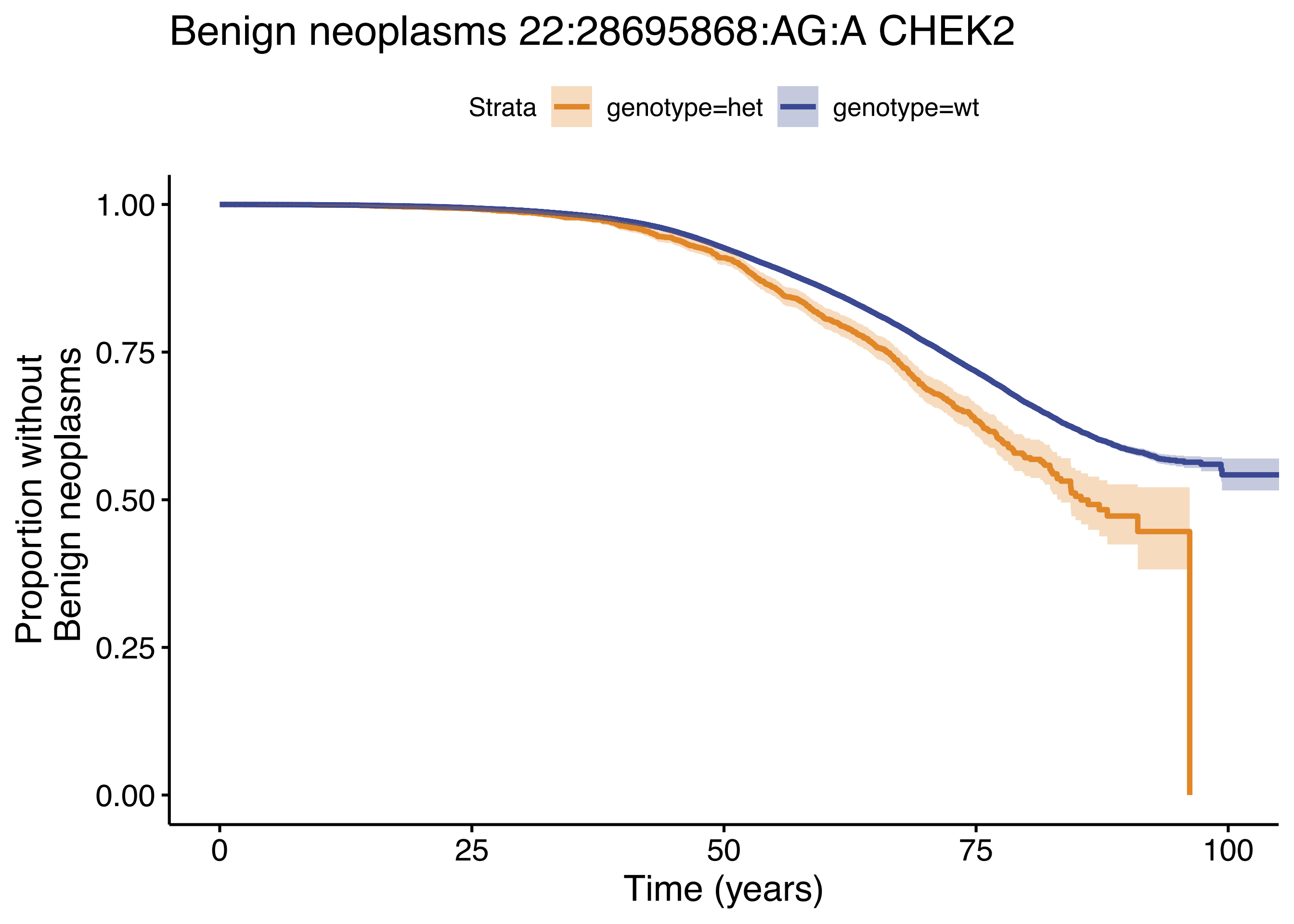
Benign neoplasms 22:28695868:AG:A CHEK2
Strata
genotype=het
genotype=wt
Proportion without
Benign neoplasms
1.00
0.75
0.50
0.25
0.00
0
25
50
75
100
Time (years)

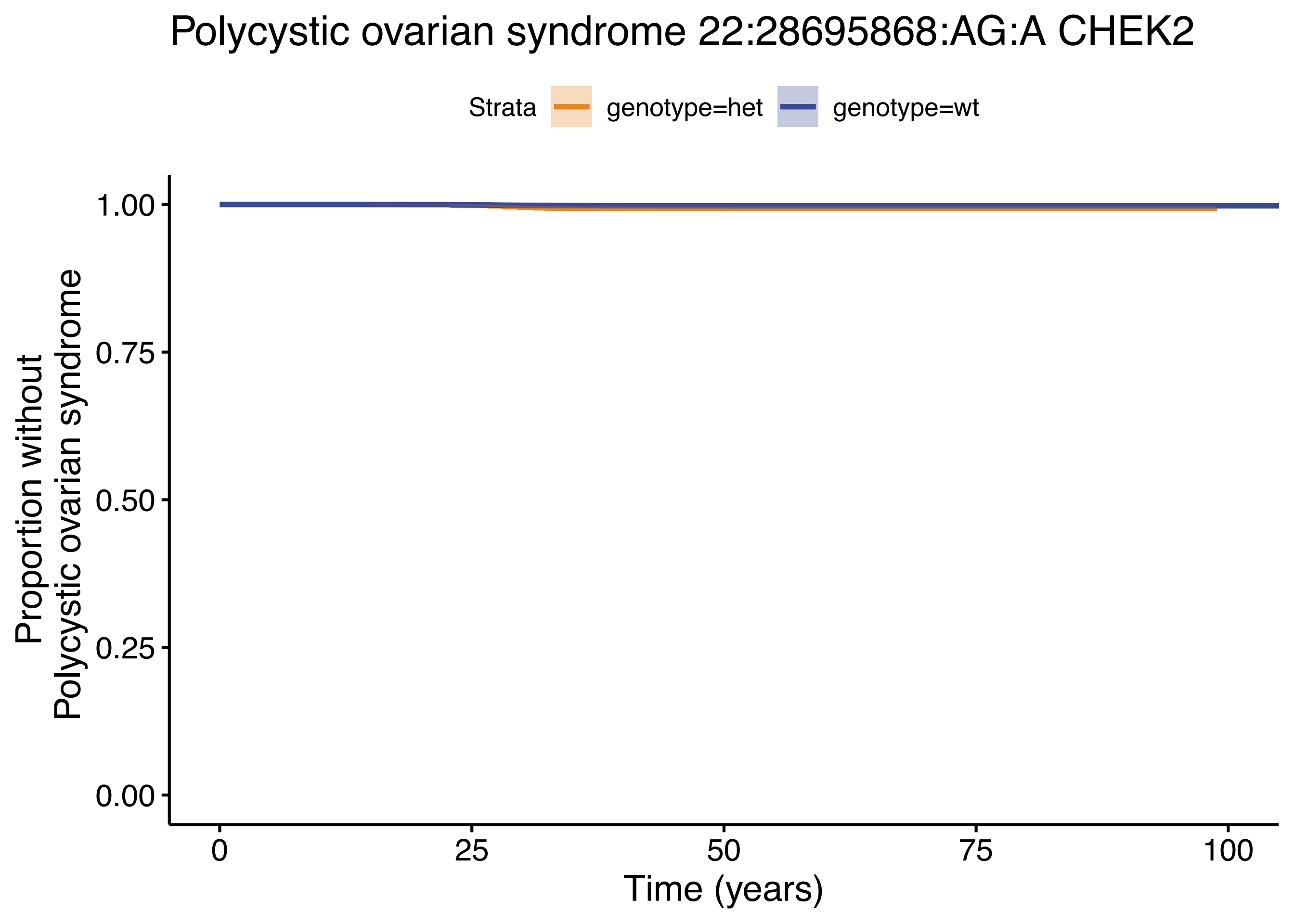

Polycystic ovarian syndrome 22:28695868:AG:A CHEK2
Strata
genotype=het
genotype=wt
1.00
0.75
0.50
0.25
0.00
Proportion without
Polycystic ovarian syndrome
0
25
50
75
100
Time (years)

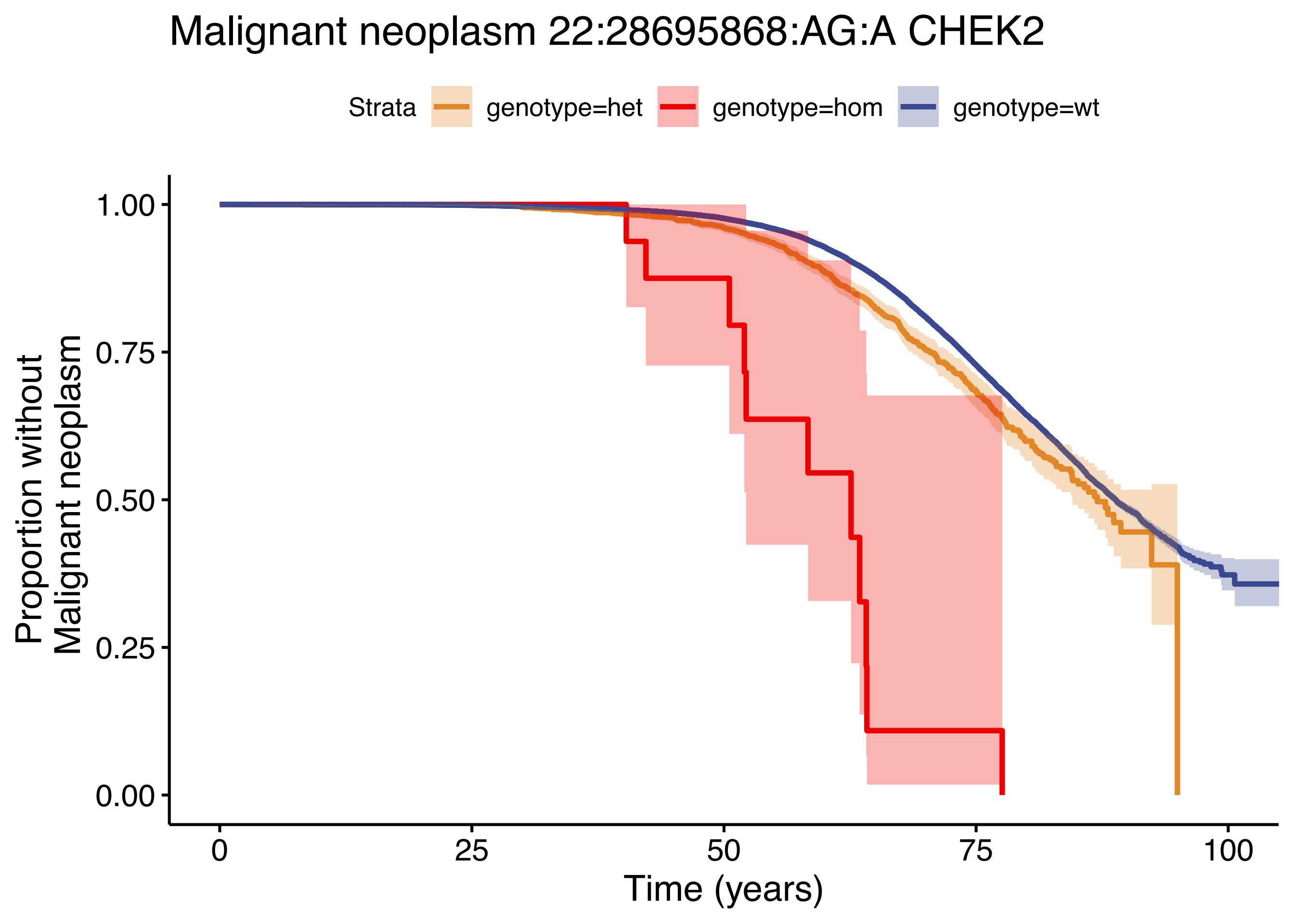

Malignant neoplasm 22:28695868:AG:A CHEK2
Strata
genotype=het
genotype=hom
genotype=wt
Proportion without
Malignant neoplasm
1.00
0.75
0.50
0.25
0.00
0
25
50
75
100
Time (years)

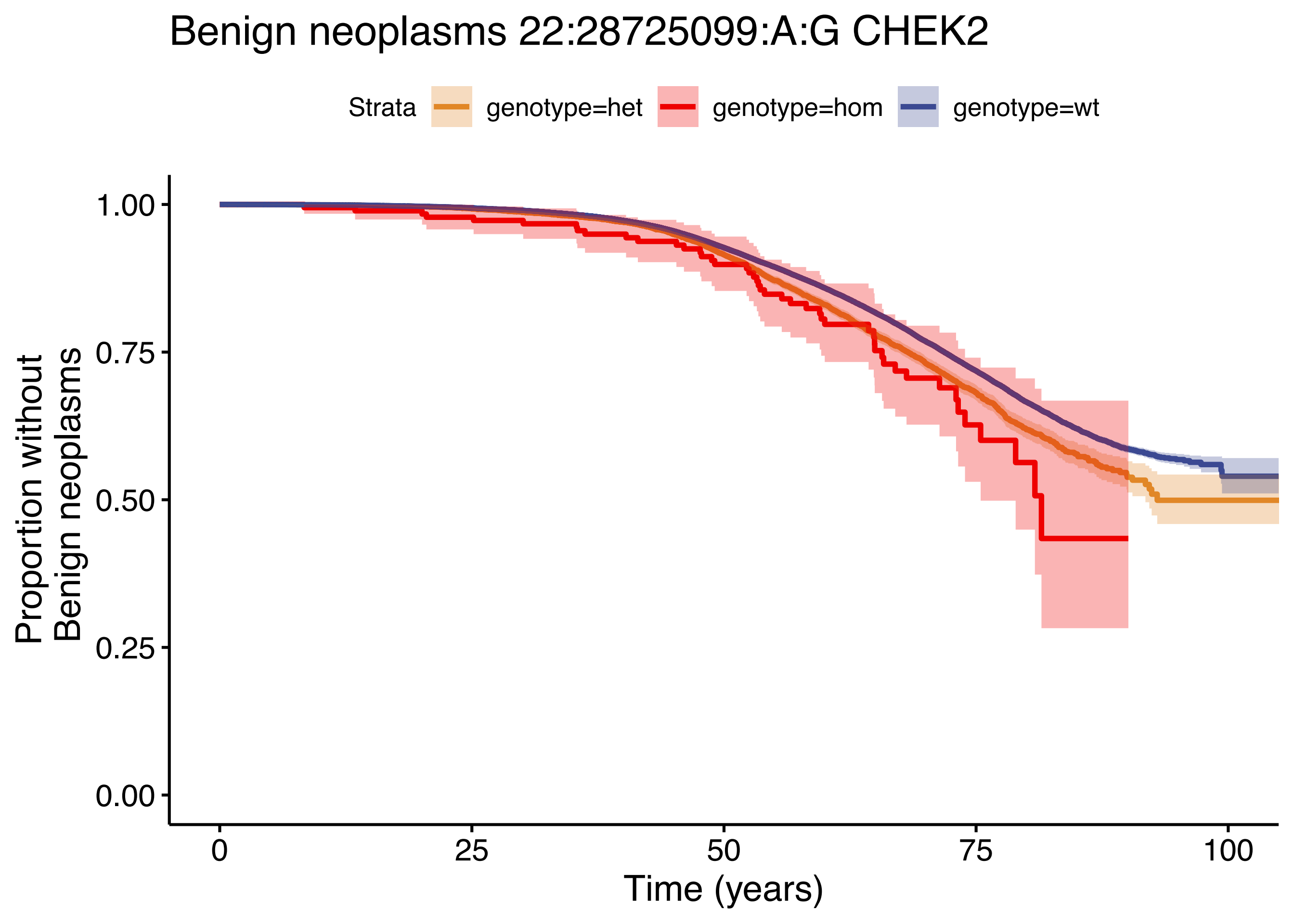

Benign neoplasms 22:28725099:A:G CHEK2
Strata
genotype=het
genotype=hom
genotype=wt
Proportion without Benign neoplasms
1.00
0.75
0.50
0.25
0.00
0
25
50
75
100
Time (years)

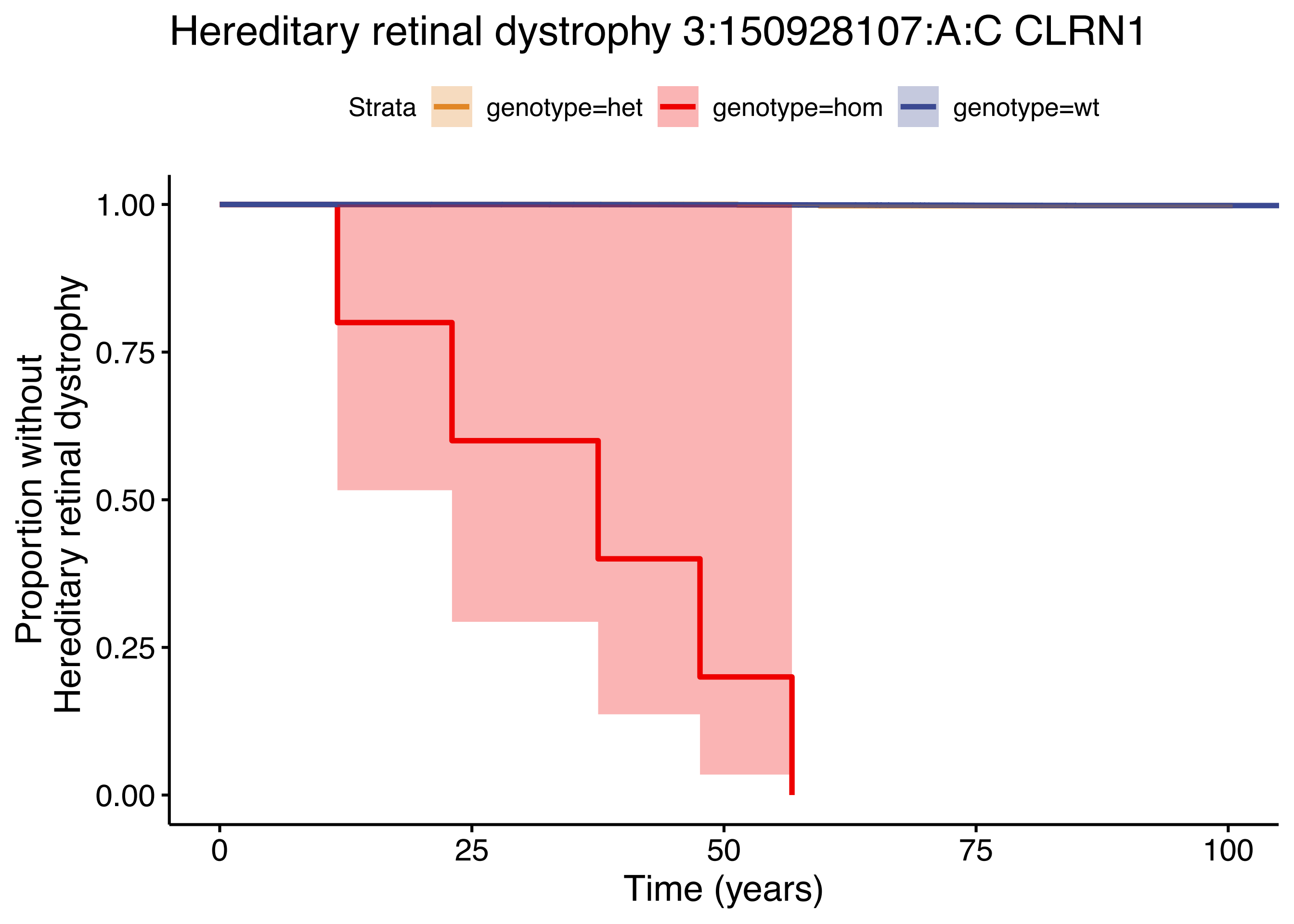

Hereditary retinal dystrophy 3:150928107:A:C CLRN1
Strata
genotype=het
genotype=hom
genotype=wt
Proportion without
Hereditary retinal dystrophy
1.00
0.75
0.50
0.25
0.00
0
25
50
75
100
Time (years)

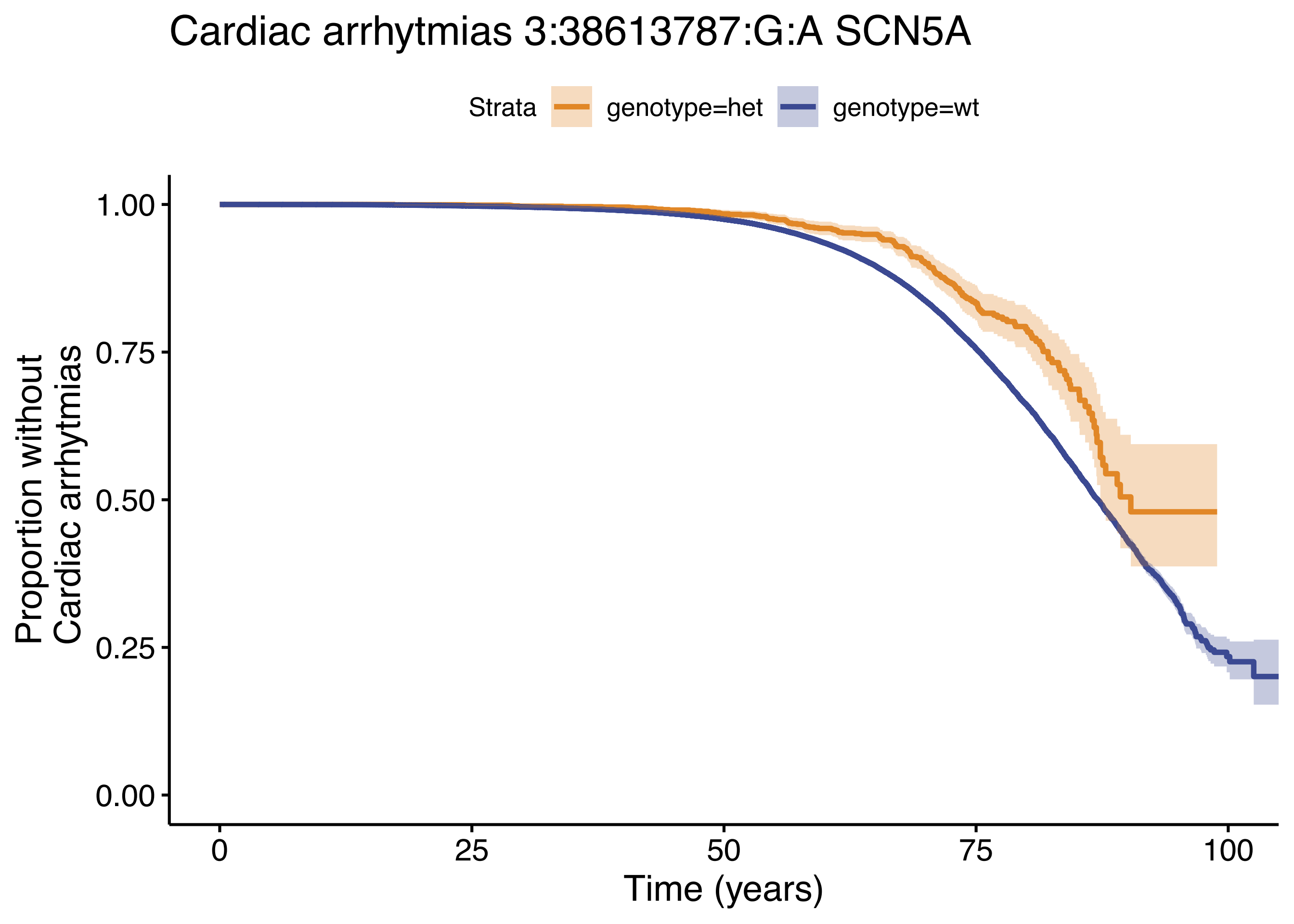

Cardiac arrhytmias 3:38613787:G:A SCN5A
Strata
genotype=het
genotype=wt
Proportion without
Cardiac arrhytmias
1.00
0.75
0.50
0.25
0.00
0
25
50
75
100
Time (years)

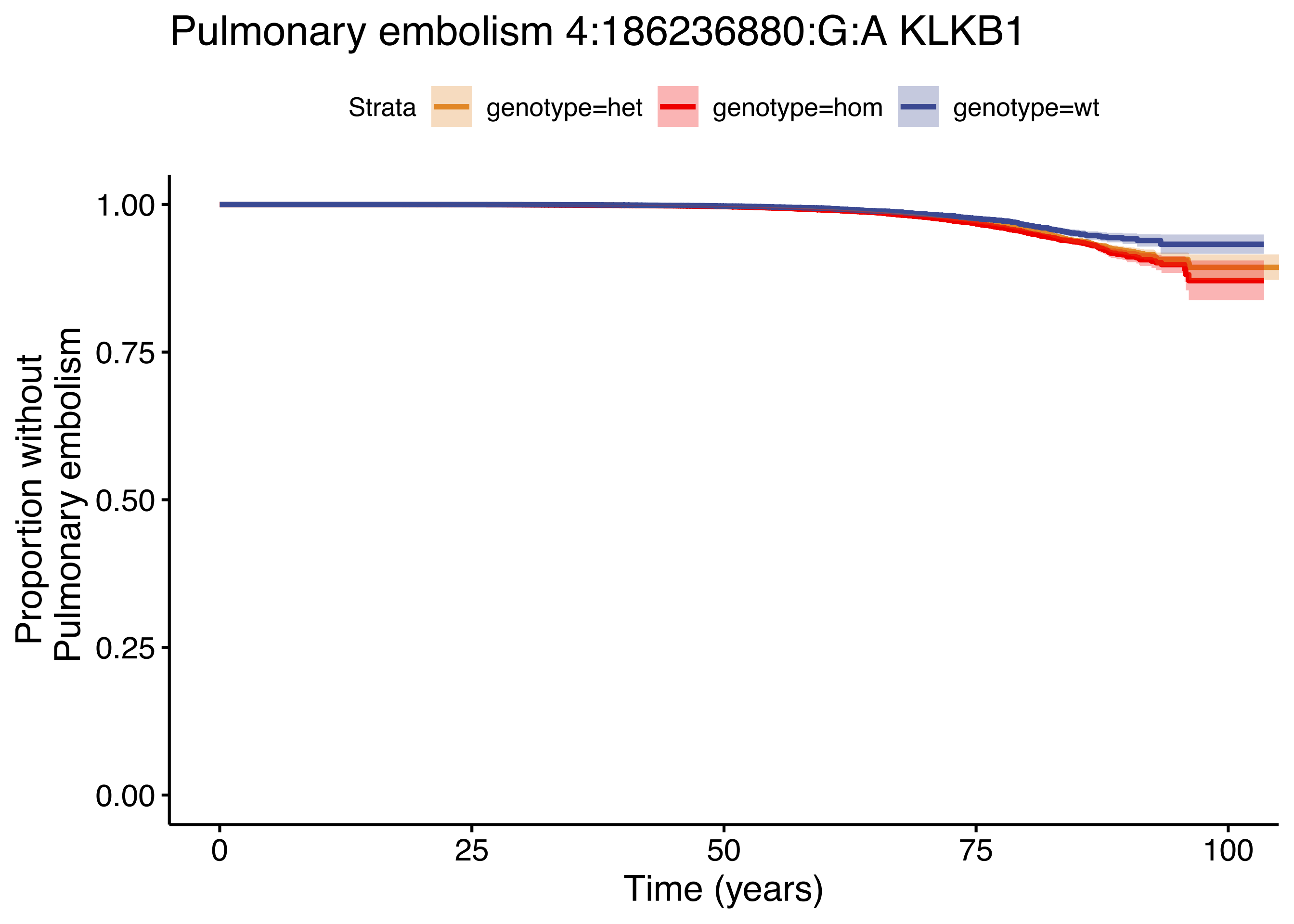

Pulmonary embolism 4:186236880:G:A KLKB1
Strata
genotype=het
genotype=hom
genotype=wt
Proportion without Pulmonary embolism
1.00
0.75
0.50
0.25
0.00
0
25
50
75
100
Time (years)

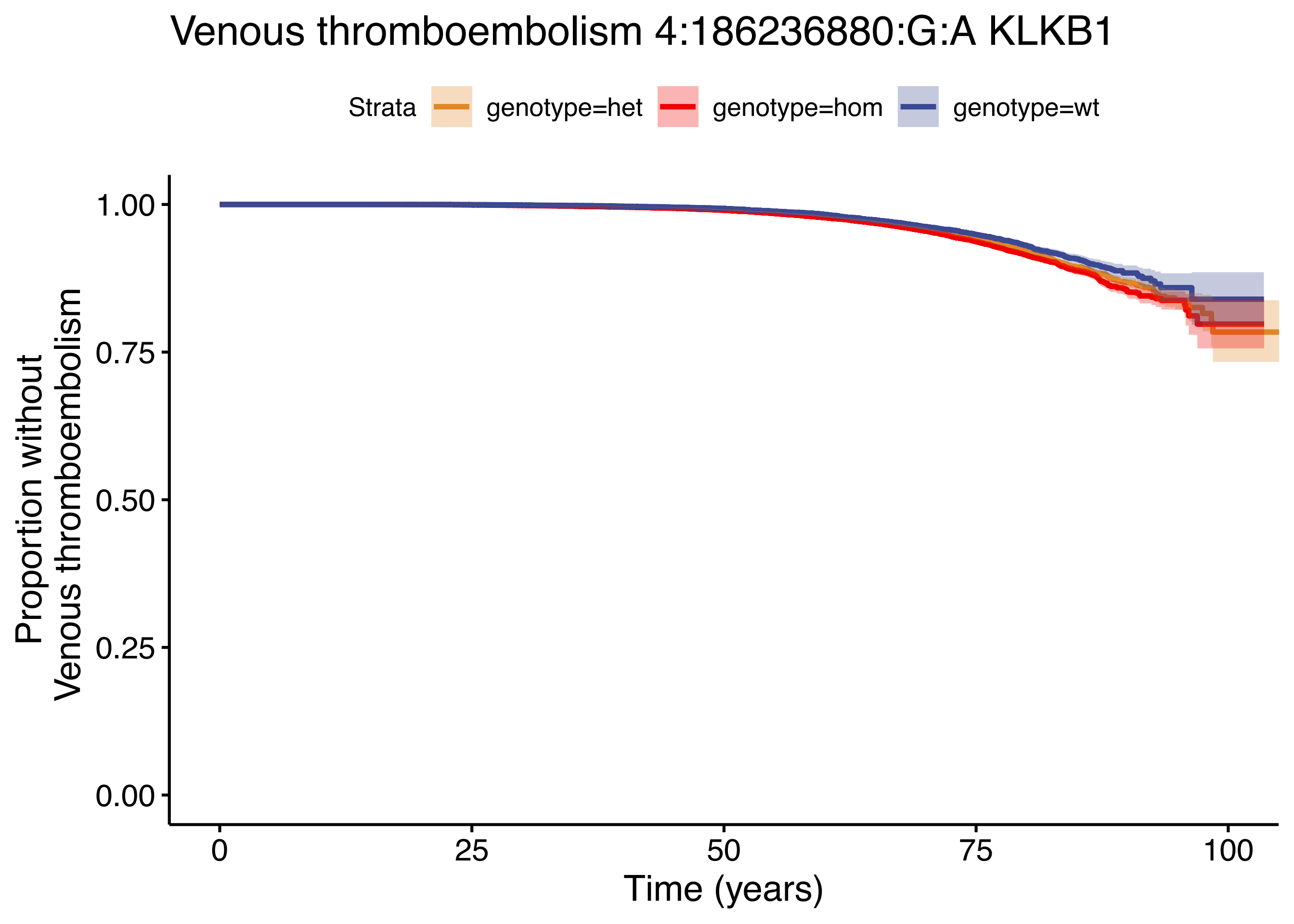
Venous thromboembolism 4:186236880:G:A KLKB1
Strata
genotype=het
genotype=hom
genotype=wt
Proportion without
Venous thromboembolism
1.00
0.75
0.50
0.25
0.00
0
25
50
75
100
Time (years)

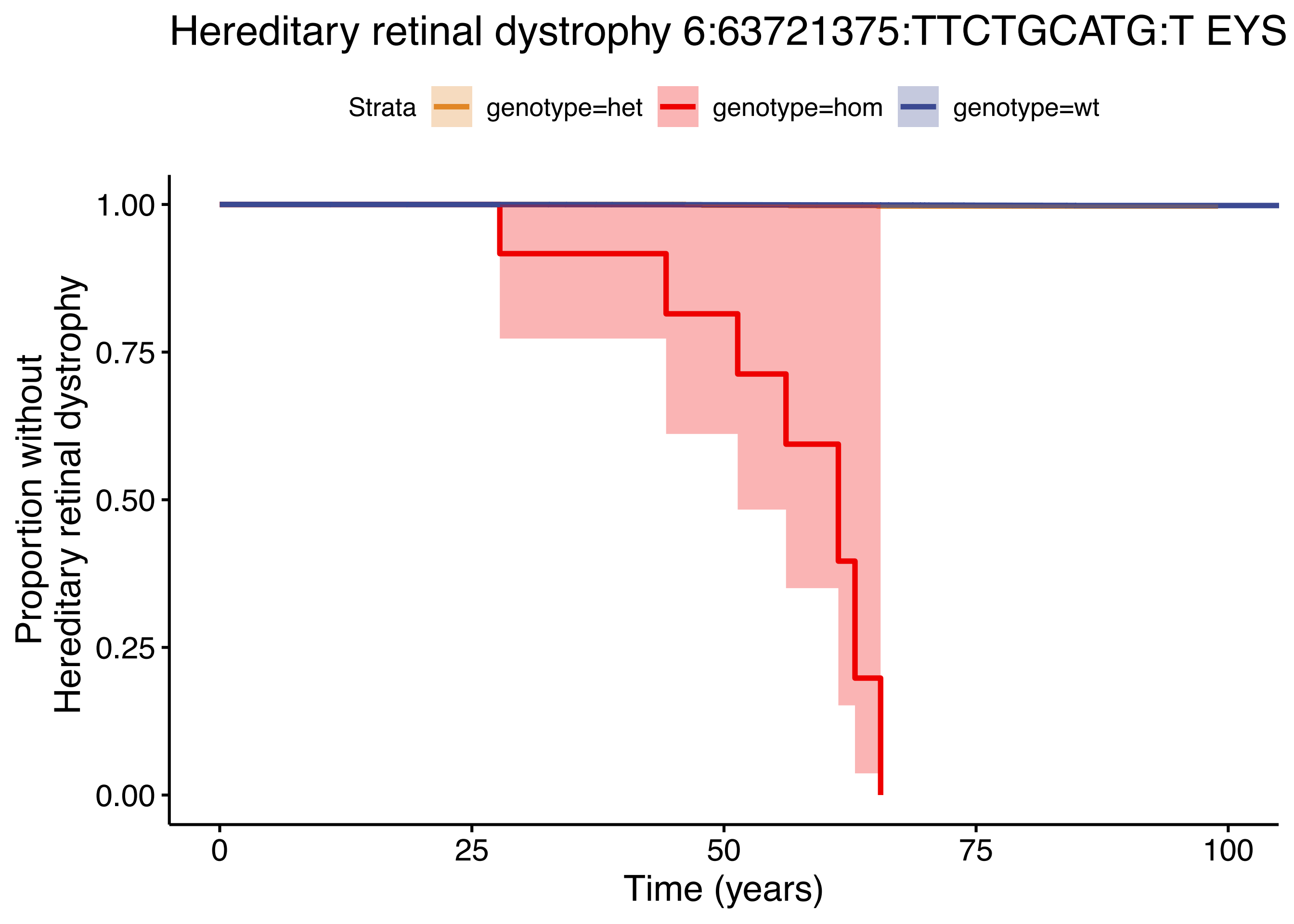

Hereditary retinal dystrophy 6:63721375:TTCTGCATG:T EYS
Strata
genotype=het
genotype=hom
genotype=wt
1.00
0.75
0.50
0.25
0.00
Proportion without
Hereditary retinal dystrophy
0
25
50
75
100
Time (years)

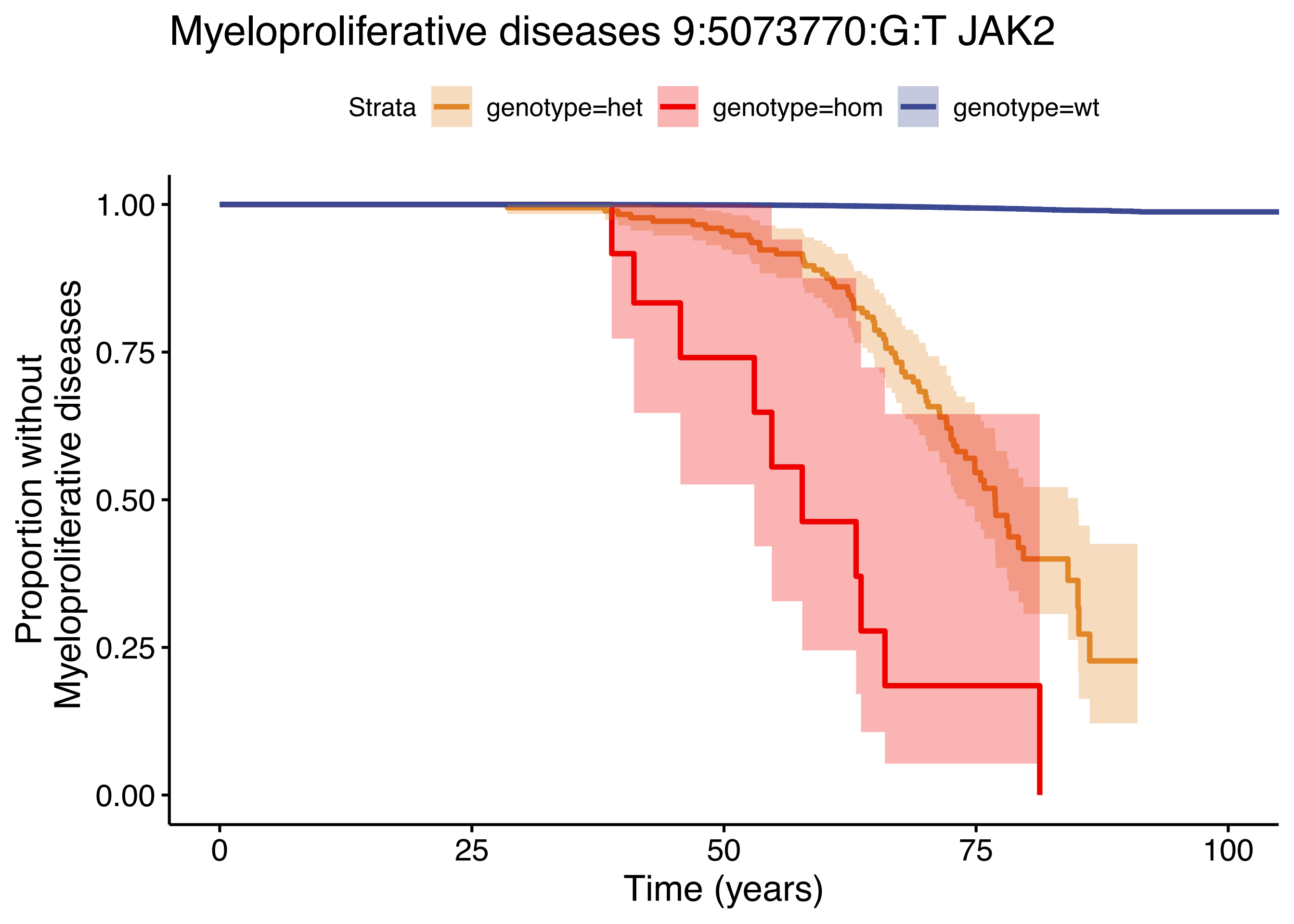

Myeloproliferative diseases 9:5073770:G:T JAK2
Strata
genotype=het
genotype=hom
genotype=wt
Proportion without Myeloproliferative diseases
1.00
0.75
0.50
0.25
0.00
0
25
50
75
100
Time (years)

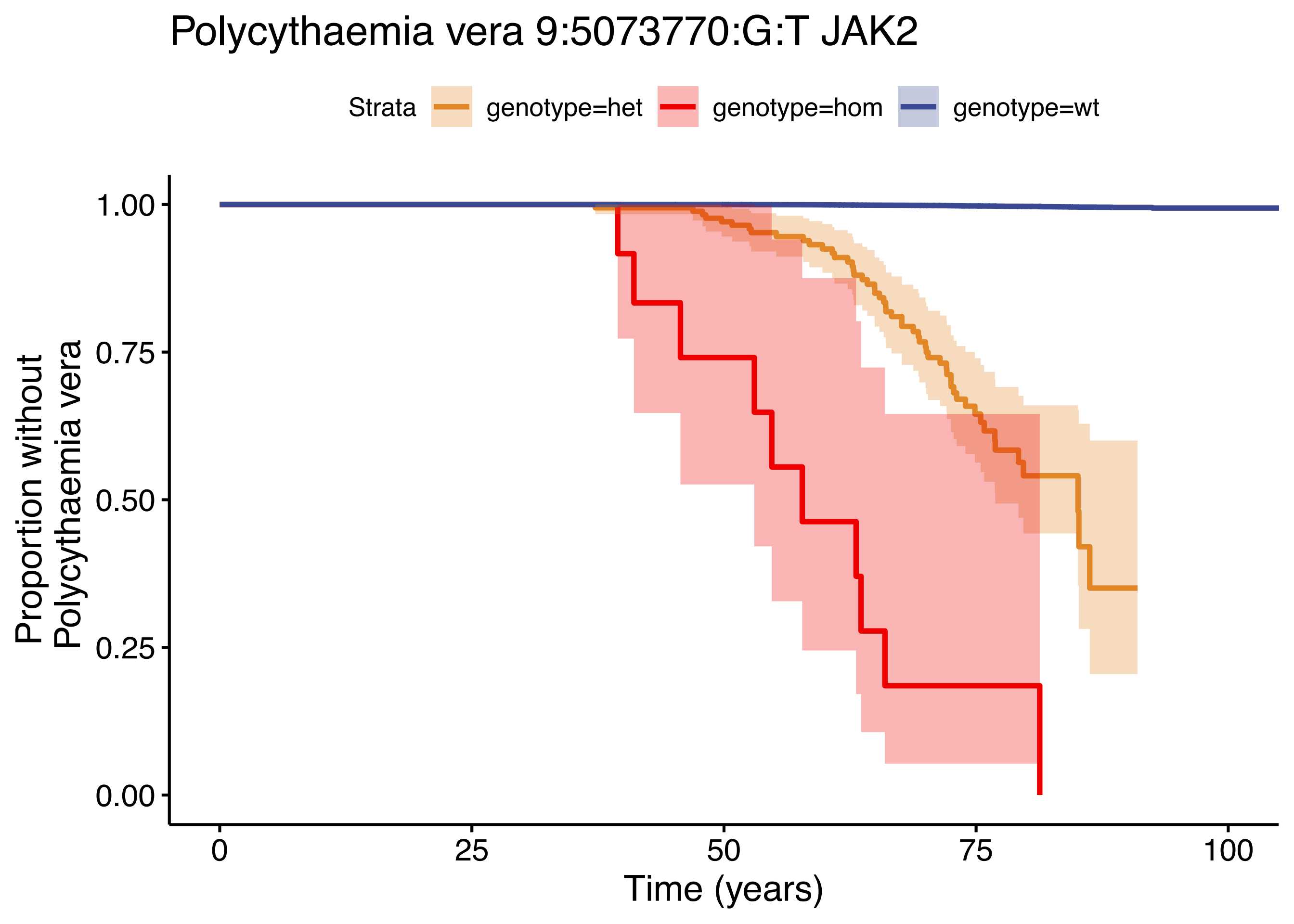

Polycythaemia vera 9:5073770:G:T JAK2
Strata
genotype=het
genotype=hom
genotype=wt
Proportion without Polycythaemia vera
1.00
0.75
0.50
0.25
0.00
0
25
50
75
100
Time (years)

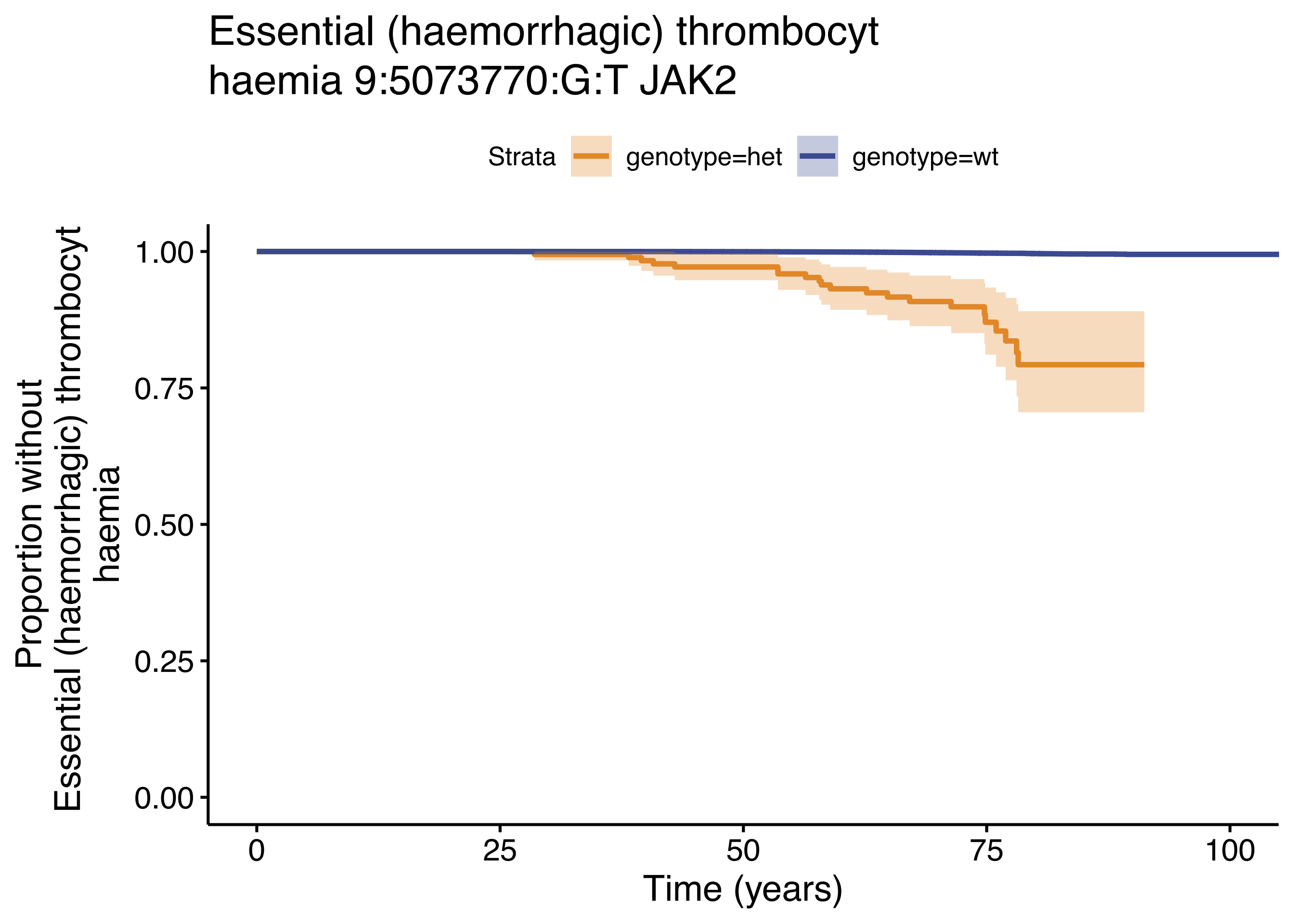
Essential (haemorrhagic) thrombocyt
haemia 9:5073770:G:T JAK2
Strata
genotype=het
genotype=wt
Proportion without
Essential (haemorrhagic) thrombocyt
haemia
1.00
0.75
0.50
0.25
0.00
0
25
50
75
100
Time (years)

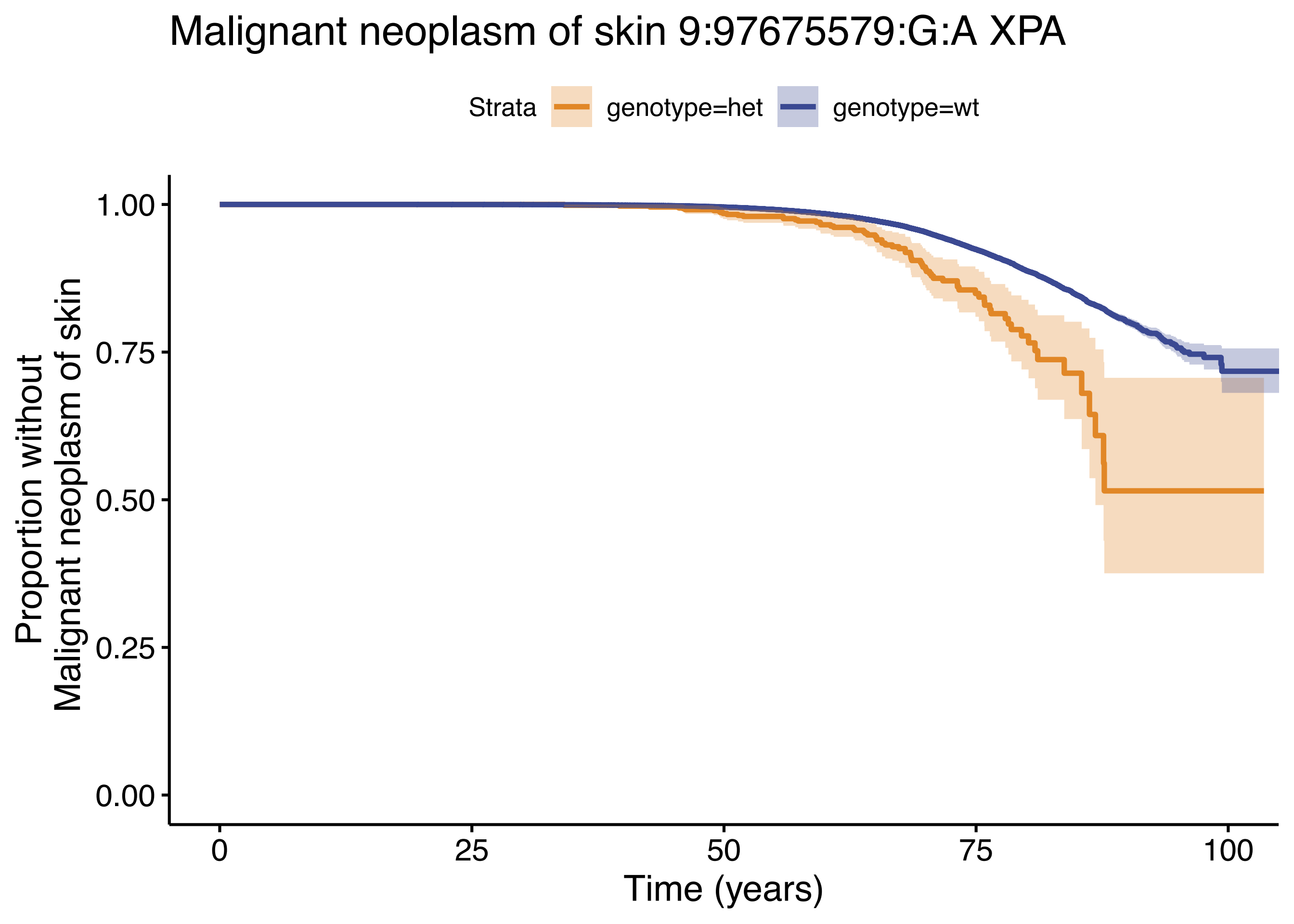
Malignant neoplasm of skin 9:97675579:G:A XPA
Strata
genotype=het
genotype=wt
Proportion without
Malignant neoplasm of skin
1.00
0.75
0.50
0.25
0.00
0
25
50
75
100
Time (years)
